# Long-term safety and efficacy of treating symptomatic, partial-thickness rotator cuff tears with fresh, uncultured, unmodified, autologous, adipose-derived regenerative cells isolated at the point of care: 41 months follow-up of a prospective, randomized, controlled, first-in-human clinical trial

**DOI:** 10.1101/2022.12.14.22283447

**Authors:** Mark Lundeen, Jason L. Hurd, Matthew Hayes, Meredith Hayes, Tiffany R. Facile, John P. Furia, Nicola Maffulli, Christopher Alt, Eckhard U. Alt, Christoph Schmitz, David A. Pearce

**Affiliations:** Sanford Orthopedics & Sports Medicine Fargo, Fargo, ND, USA; Sanford Orthopedics & Sports Medicine Sioux Falls, Sioux Falls, SD, USA; Sanford Radiology Clinic, Sioux Falls, SD, USA; Sanford Health, Sioux Falls, SD, USA; SUN Orthopedics of Evangelical Community Hospital, Lewisburg, PA, USA; Department of Musculoskeletal Disorders, Faculty of Medicine and Surgery, University of Salerno, Salerno, Italy; Centre for Sports and Exercise Medicine, Barts and The London School of Medicine and Dentistry, Mile End Hospital, Queen Mary University of London, London, UK; School of Pharmacy and Bioengineering, Guy Hilton Research Centre, Keele University School of Medicine, Stoke on Trent, UK; InGeneron, Inc., Houston, TX, USA; Institute of Anatomy, Faculty of Medicine, LMU Munich, Munich, Germany; Isar Klinikum, Munich, Germany; Sanford School of Medicine, University of South Dakota, Sioux Falls, SD, USA; Heart and Vascular Institute, Department of Medicine, Tulane University Health Science Center, New Orleans, LA, USA; Sanford Research, Sioux Falls, SD, USA

**Keywords:** UA-ADRCs, Safety, Shoulder disease, Stem cells, Stromal vascular fraction

## Abstract

**Background:** Symptomatic, partial-thickness rotator cuff tears (sPTRCT) are problematic. Management of sPTRCT with fresh, uncultured, unmodified, autologous, adipose-derived regenerative cells (UA-ADRCs) isolated from lipoaspirate at the point of care is safe and leads to improved shoulder function without adverse effects. This study tested the hypothesis that management of sPTRCT with injection of UA-ADRCs is safe and more effective than injection of corticosteroid even in the long run.

**Methods:** Subjects who had completed a former randomized controlled trial were enrolled in the present study. At baseline these subjects had not responded to physical therapy treatments for at least six weeks, and were randomly assigned to receive either a single injection of an average 11.4 × 106 UA-ADRCs (n = 11) or a single injection of 80 mg of methylprednisolone (n = 5). Safety was assessed by rigorously documenting and evaluating treatment emergent adverse events. As per protocol efficacy was assessed using the ASES Total score, RAND Short Form-36 Health Survey (SF-36) Total score and VAS pain score at 24 weeks (W24) and W52 post-treatment as well as at 33.2 ± 1.0 (mean ± standard deviation) months (M33) and 40.6 ± 1.9 months (M41) post-treatment. Magnetic resonance imaging (MRI) of the index shoulder was performed at baseline, W24, W52, M33 and M41 post-treatment.

**Results:** There were no greater risks connected with injection of UA-ADRCs than those connected with injection of corticosteroid. Injection of UA-ADRCs resulted in significantly higher mean ASES Total scores at W24, W52 and M41, a significantly higher mean SF-36 Total score at W24, and significantly higher mean VAS Pain scores at W24 and W52 post-treatment than injection of corticosteroid (p<0.05). Treatment outcome could not be assessed using measurements of tear volume on MRI scans. On the other hand, MRI scans at W24 post-treatment allowed to “watch the UA-ADRCs at work”. There was no relationship between treatment outcome and baseline data, including those data characterizing UA-ADRCs that can be collected with a clinical test.

**Conclusions:** The present study further supports management of sPTRCT with injection of UA-ADRCs.

**Trial registration:** Clinicaltrials.gov NCT04077190 (September 4, 2019).

## INTRODUCTION

Symptomatic, partial-thickness rotator cuff tear (sPTRCT) is a common cause of shoulder pain, loss of function and occupational disability [1–3]. Cadaveric and magnetic resonance imaging (MRI) studies reported the incidence of partial-thickness rotator cuff tears between 13% and 25%, with an increasing incidence with age [4–6]. The majority of sPTRCT cases are associated with aging, repetitive overhead use of the arm, sudden and forceful trauma, or a combination of these factors [1–3].

Current non-surgical and surgical treatment options to address sPTRCT do not offer the potential to naturally replace damaged tendon tissue and often do not improve clinical results. Subacromial injection of corticosteroids, among the most widely used nonoperative treatment options for sPTRCT [7], can provide short-term pain relief but may not modify the course of the condition [7]. Even worse, subacromial injection of corticosteroid carries the risk that a partial-thickness rotator cuff tear develops into a full-thickness rotator cuff tear [8]. A recent meta-analysis and a recent double-blinded, randomized controlled clinical trial (RCT) concluded that injections of platelet rich plasma might also not be beneficial in non-operative treatment of rotator cuff disease [9,10]. Surgical treatment of sPTRCT is generally successful among patients who, for a period of 3 to 6 months, unsuccessfully underwent conservative treatment modalities [2]. However, surgical intervention presents potential complications and a more lengthy recovery, and some authors have argued that results from these procedures may not exceed those obtained with conservative management [11].

A recent, first-in-human RCT [12] (hereafter: the former study) indicated that treatment of sPTRCT with fresh, uncultured, unmodified, autologous, adipose-derived regenerative cells (UA-ADRCs) isolated from lipoaspirate at the point of care is safe and leads to improved shoulder function without adverse effects. This study also showed that the risks associated with treating sPTRCT with UA-ADRCs were as low as those associated with injection of corticosteroid over 12 months post-treatment, with no serious adverse events observed for either treatment [12].

Unlike most other cell preparations currently under investigation for use in regenerative medicine (including cultured adipose derived stem cells (ADSCs), induced pluripotent stem cells, etc.) UA-ADRCs are not expanded in culture, and are therefore not exposed to potential, culture-related mechanic and oxidative stress that could affect their safety as a medicinal product [13]. Furthermore, UA-ADRCs do not share the risk of potentially developing tumors (reported for induced pluripotent stem cells) and immunological defensive reactions (reported for allogeneic adult stem cells) [14,15]. Only 0.001–0.1% of the total population of bone marrow nucleated cells represent mesenchymal stromal cells, whereas these cells can represent up to 12% of the total population of UA-ADRCs [16,17]. Additionally, harvesting adipose tissue is typically much less invasive than harvesting bone marrow [18].

In the former study [12] subjects were not followed up beyond 12 months post-treatment. The present study tested the hypothesis that treatment of sPTRCT with injection of UA-ADRCs is safe and more effective than injection of corticosteroid even in the long run, with a minimum follow-up of 36 months.

## METHODS

### Study design

The present study was a long term follow-up study of a first-in-human, two center, prospective, open-label, randomized controlled trial [12]. Both the present and the former studies [12] were conducted at Sanford Orthopedics and Sports Medicine – Fargo (Fargo, ND, USA) (PI: M.L.) and Sanford Orthopedics and Sports Medicine – Sioux Falls (Sioux Falls, SD, USA) (principal investigator (PI): J.H.).

Fig. 1 shows the flow of subjects in the present and the former studies [12] according to the CONSORT statement [19].

**FIGURE 1.**
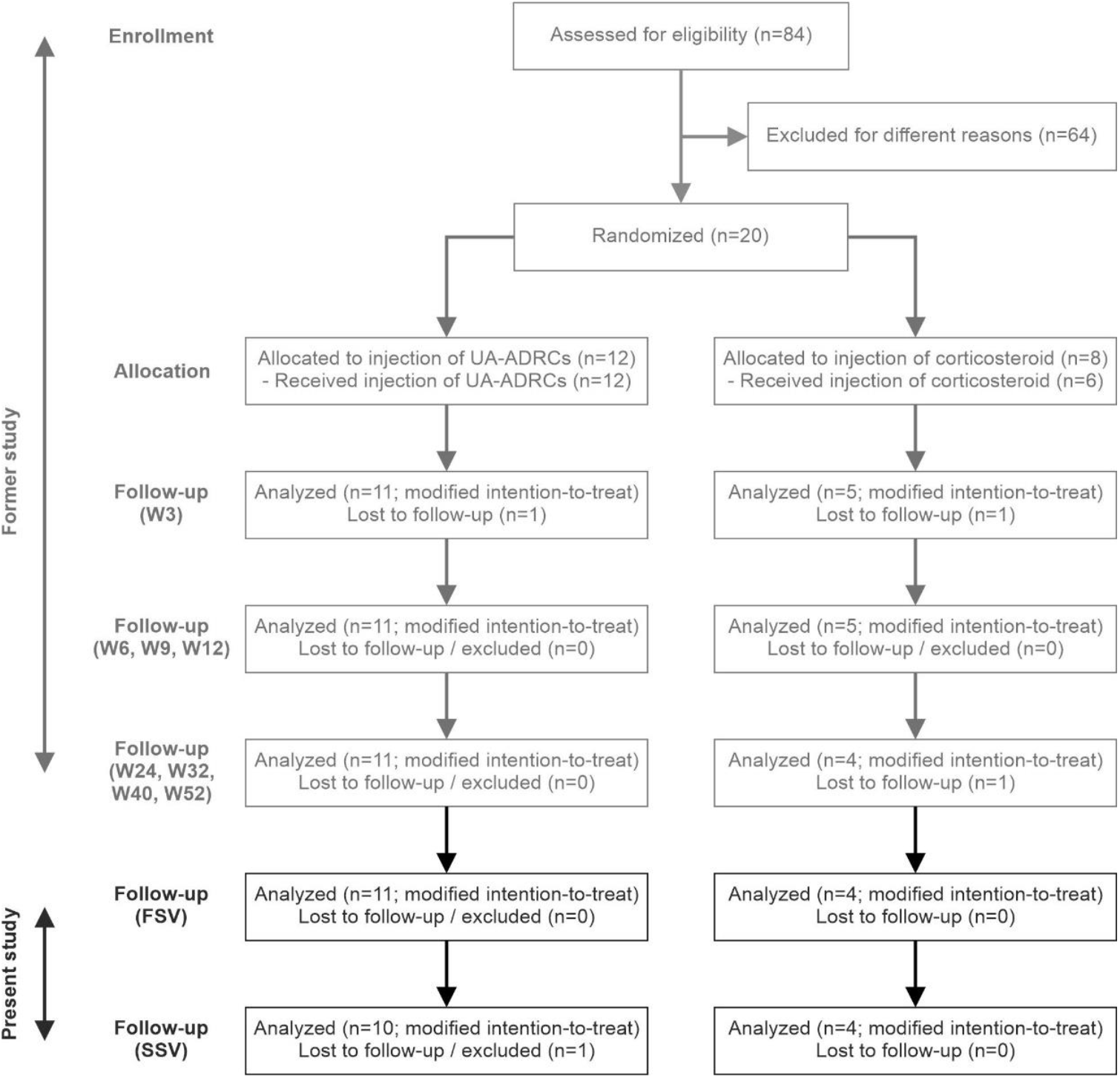
Flow of subjects in the present and the former studies [12] according to CONSORT [19]. Abbreviations: W3 / W6 / W9 / W12 / W24 / W32 / W40 / W52, study visits scheduled in the former study [12] at 3 / 6 / 9 / 12 / 24 / 32 / 40 / 52 weeks post-treatment; FSV, first study visit of the present study at 33.2 ± 1.0 (mean ± standard deviation) months post-treatment; SSV, second study visit of the present study at 40.6 ± 1.9 months post-treatment.

### Ethics

The former study [12] received Investigational Device Exemption (IDE) from the U.S. Food and Drug Administration (FDA) on September 23, 2016 (no. 16956), was registered at Clinicaltrials.gov on September 28, 2016 (ID NCT02918136), and received Institutional Review Board (IRB) approval of Sanford Health (Sioux Falls, SD, USA) on November 4, 2016 (Sanford IRB #3 registration number 00007985) in accordance with the Declaration of Helsinki. The first subject was enrolled in the former study [12] on January 04, 2017, and the last subject on April 21, 2017. The study was closed on November 7, 2019. After having received additional IRB approval from Sanford Health on November 07, 2019 (Sanford IRB #3 registration number STUDY00001869) to re-examine MRI scans, the former study [12] was re-opened on September 14, 2020.

The present study received IDE from the FDA on May 13, 2019, received IRB approval from WIRB Copernicus Group, Inc. (Olympia, WA, USA) on July 23, 2019 for study site Sanford USD Medical Center, Sioux Falls, SD, USA and on September 29, 29 for study site Sanford Orthopedic Sports Medicine Clinic, Fargo, ND, USA (IRB Tracking Number: 20191931), and was registered at Clinicaltrials.gov on September 4, 2019 (NCT04077190). The first subject was enrolled in the present study on November 21, 2019, and the last subject on March 20, 2020. The present study was closed on May 09, 2022.

### Participants, randomization and interventions

In brief, all the subjects enrolled in the former study [12] suffered from a sPTRCT of the supraspinatus tendon at baseline, had not responded to physical therapy treatments for at least six weeks, and were randomly assigned to receive either a single injection of an average 11.4 × 10^6^ UA-ADRCs (in 5 mL liquid; mean cell viability: 88%) (n=11; modified intention-to-treat (mITT) population) (hereafter: UA-ADRCs group) or a single injection of 80 mg of methylprednisolone (40 mg/mL; 2 mL) plus 3 mL of 0.25% bupivacaine (n=5 in the former study [12]; n=4 in the present study; mITT population) (hereafter: corticosteroid group). The UA-ADRCs were isolated from lipoaspirate using the Transpose RT system (InGeneron, Houston, TX, USA) [15,20]. One subject in the corticosteroid group experienced progression of sPTRCT into a symptomatic, full-thickness rotator cuff tear during the former study [12] and was therefore not enrolled in the present study. For this reason, the baseline data of the subjects in the corticosteroid group enrolled in the present study (summarized in Table 1) slightly differ from the baseline data of those in the corticosteroid group provided in the former study [12].

**TABLE 1.**
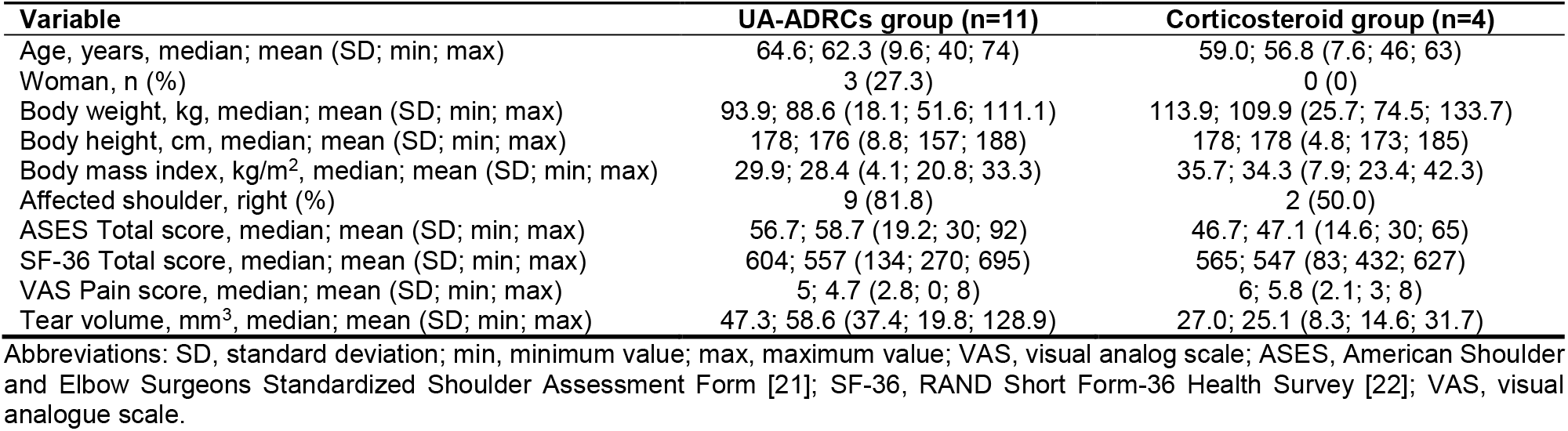
Characteristics of the subjects enrolled in the present study at baseline (modified intention-to-treat population).

### Outcome measurements and assessments

According to the study protocol the primary endpoints of the present study were long-term safety as indicated through the rate of treatment emergent adverse events (TEAEs), and long-term efficacy of pain and function through American Shoulder and Elbow Surgeons Standardized Shoulder Assessment Form [21] (ASES Total score) and RAND Short Form-36 [22] (SF-36) health questionnaires between the UA-ADRCs group and the corticosteroid group. The secondary endpoint of the present study was long-term efficacy evaluated through VAS Pain score and MRI pre- and post-injection for the therapeutic intent to treat sPTRCT between the UA-ADRCs group and the corticosteroid group.

Adverse events in the present and the former studies [12] were defined as any untoward or unfavorable medical occurrence in a subject, including any abnormal sign, symptom or disease temporally associated with the subject’s participation in these studies, whether or not considered related to the subject’s participation in these studies.

In the former study [12], safety was assessed immediately after treatment and three weeks (W3), W6, W9, W12, W24, W32, W40 and W52 post-treatment; ASES Total score, SF-36 Total score and VAS Pain score were assessed at baseline (BL) and at W3, W6, W9, W12, W24, W32, W40 and W52 post-treatment; MRI was performed at BL and at W24 and W52 post-treatment.

In the present study, the primary and secondary endpoints were assessed at 33.2 ± 1.0 (mean ± standard deviation) months post-treatment (range, 30.7 – 34.7) (first study visit; FSV) and at 40.6 ± 1.9 months post-treatment (range, 36.5 – 44.7) (second study visit; SSV).

### Analysis of MRI scans

Next to the determination of the partial-thickness tear size (calculated as ellipsoid volume), the proton density weighted, fat saturated, T2-weighted (PD FS T2) coronal MRI scans of all subjects who were enrolled in the present study were transferred in digital and fully anonymized form (compliant with the HIPAA regulation) [23] to C.S. who was only aware of the Subject IDs. Then, C.S. mounted these MRI scans as shown in Figs S1–S15 (**Figs S1–S21 and Tables S1–S21 are in Appendix 1**), evaluated them and indicated hyperintense structures at the position of the supraspinatus tendon that were present at W24 post-treatment but not at baseline (arrows in Figs S1–S7 and S9-S11). Afterwards, the files with the MRI scan montages and the indicated hyperintense structures were transferred in fully anonymized form (even without the Subject IDs) to M.H. and M.H., who performed an independent, blinded re-analysis of the hyperintense structures at the position of the supraspinatus tendon indicated by C.S.

### Estimand strategies for handling intercurrent events

In line with the new *The International Council for Harmonisation of Technical Requirements for Pharmaceuticals for Human Use (ICH) E9 (R1) Addendum* on the use of estimands in clinical trials (i.e., a precise description of the treatment effect to be estimated from a trial (the question) [24–26]) a comprehensive estimand was constructed for the present study. The four components of this estimand (Population (i.e., the target population for the research question), Variables (i.e., the endpoints that were obtained from all subjects), Intercurrent Events (i.e., all events that occurred after treatment initiation and either precluded the observation of a variable, or affected its interpretation) and Population-Level Summary (i.e., the variables on which the comparison between treatments was based) are outlined in **Appendix 2**.

In short, in case of treatment failures (comprising all intercurrent events that required additional injections of corticosteroid into the index shoulder or surgery of the index shoulder that were definitely, probably or possibly related to the study treatments, including development of a full-thickness rotator cuff tear) subjects’ data were handled using a combination of the While-on-Treatment Strategy [24–26] and the Composite Strategy [24–26]. Specifically, response to study treatment before the occurrence of the intercurrent event was handled using the While-on-Treatment Strategy (i.e., subjects’ data were used as collected), whereas response to study treatment after the occurrence of the intercurrent event was imputed according to the Composite Variable Strategy as minimum ASES Total score (0), minimum SF-36 Total score (0), maximum VAS pain score (10) and maximum tear volume measured on MRIs (150 mm^3^, which was greater than all data measured during the present and the former studies [12]).

In contrast, intercurrent events that required additional injections of corticosteroid into the index shoulder or surgery of the index shoulder that were unlikely related or unrelated to the study treatments (e.g., accidents that affected the index shoulder) were handled using a combination of the While-on-Treatment Strategy and a Hyopthetical Strategy [24–26]. Specifically, response to the study treatment before the occurrence of the intercurrent event was handled according to the While-on-Treatment Strategy, whereas response to the study treatment after the occurrence of the intercurrent event was imputed according to a Hypothetical Strategy in which the intercurrent event would not occur. Imputation of subjects’ data after occurrence of the intercurrent event was performed using the Last Observation Carried Forward approach [27,28].

### Statistical analysis

Statistical analysis of the safety data included group-specific comparisons of the following variables: (i) total number of TEAEs, (ii) number of TEAEs experienced per subject, (iii) number of TEAEs classified as {mild / moderate / severe}, (iv) relationship of TEAEs to treatment classified as {not related / unlikely / possible / probable / definite} and (v) number of TEAEs classified as {mild and unlikely to be related to the investigated treatment / mild and possibly related to the investigated treatment / moderate and unlikely to be related to the investigated treatment / moderate and possibly related to the investigated treatment}. According to the protocol of the present study these comparisons were performed for the following time periods: from BL to W24 post-treatment (considering only data of the former study [12]), from BL to FSV in the present study, and from BL to SSV in the present study (each considering data of the present and the former studies [12]). Comparisons were performed using Chi-square test or Chi-square test for trend, respectively.

Statistical analysis of the efficacy data included calculation of the group specific mean, standard error of the mean and median as well as group-specific comparisons of the following variables: (i) ASES Total score, (ii) SF-36 Total score, (iii) VAS pain score and (iv) tear volume measured on MRIs. According to the protocol of the present study these comparisons were performed at BL and at W24 and W52 post-treatment (data of the former study [12]) as well as at FSV and SSV (data of the present study). Given the non-parametric nature of all efficacy data after imputation according to the estimand, comparisons were performed using the Mann-Whitney test.

In all analyses, a p value < 0.05 was considered statistically significant. Calculations were performed using GraphPad Prism (Version 9.4.1 for Windows; GraphPad Software, San Diego, CA, USA).

## RESULTS

### Long-term safety of treating sPTRCT with injection of either UA-ADRCs or corticosteroid

The subjects in the UA-ADRCs group reported a total number of 58 TEAEs (35 TEAEs during the former study [12] and 23 TEAEs during the present study) (details in Table S1). The subjects in the corticosteroid group reported a total number of 25 TEAEs (12 TEAEs during the former study [12] and 13 TEAEs during the present study) (details in Table S2).

No TEAE that occurred during the present and the former studies [12] was classified as probably or definitely related to the investigated treatment. Furthermore, all severe TEAEs that occurred during the present and the former studies [12] were classified as not related to the investigated treatment.

All subjects reported experiencing at least one TEAE. The number of subjects who experienced 1 / 2 / 3 / 4 / 6 / 7 / 8 / 10 / 12 TEAEs in the present and the former studies [12] was 1 / 0 / 2 / 3 / 1 / 1 / 2 / 1 / 0 in the UA-ADRCs group (5.3 ± 2.7; median, 4) and 0 / 1 / 1 / 2 / 0 / 0 / 0 / 0 / 1 in the corticosteroid group (5.0 ± 1.8 (mean ± SEM); median, 4). These data were not significantly different between the groups (Chi-square test for trend; p = 0.778) (details in Table S3).

The number of TEAEs classified as {mild / moderate / severe} in the present and the former studies [12] was 38 / 16 / 4 in the UA-ADRCs group and 17 / 8 / 0 in the corticosteroid group. These data were not significantly different between the groups (Chi-square test for trend; p = 0.497) (details in Tables S4–S6).

The relationship of TEAEs to treatment classified as {not related / unlikely / possible / probable / definite} in the present and the former studies [12] was 48 / 6 / 4 / 0 / 0 in the UA-ADRCs group and 20 / 3 / 2 / 0 / 0 in the corticosteroid group. These data were not significantly different between the groups (Chi-square test; p = 0.956) (details in Tables S7–S9).

The number of TEAEs classified as {mild and unlikely to be related to the investigated treatment / mild and possibly related to the investigated treatment / moderate and unlikely to be related to the investigated treatment / moderate and possibly related to the investigated treatment} in the present and the former studies [12] was 4 / 3 / 2 /1 in the UA-ADRCs group and 3 / 0 / 0 / 2 in the corticosteroid group. These data were not significantly different between the groups (Chi-square test for trend; p = 0.757) (details in Table S10).

The four severe TEAEs that occurred in the UA-ADRCs group during the present and the former studies [12] were non ST elevation myocardial infarction, ST elevation myocardial infarction, ganglion cyst of the non-index shoulder, and pain in the index shoulder. None of these severe TEAEs were related to treatment (details in Tables S11–S13).

### Long-term efficacy of treating sPTRCT with injection of either UA-ADRCs or corticosteroid

Four of the 11 subjects (36.4%) in the UA-ADRCs group and three of the five subjects (60.0%) in the corticosteroid group developed additional pathologies of the index shoulder (next to sPTRCT) and/or received additional injections into or surgery of the index shoulder (next to injection of either UA-ADRCs or corticosteroid) during the present and the former studies [12]. For one of the 11 subjects (9%) in the UA-ADRCs group and two of the five subjects (40%) in the corticosteroid group these additional pathologies were considered treatment failure (details in Tables S14 and S15). After these intercurrent events, individual data related to the efficacy of the investigated treatment were either missing or unsuitable for assessing treatment outcome if they had been collected after the intercurrent event during the present and the former studies [12]. These missing data were imputed according to the estimand of the present study outlined in Appendix 2.

The individual ASES Total scores, SF-36 Total scores and VAS pain scores as a function of time post-treatment are shown in Figs S16–S18; imputation of missing data is indicated in these figures. Eight of the 11 subjects (72.7%) in the UA-ADRCs group but only one of the five subjects (20%) in the corticosteroid group reached an individual ASES Total score of at least 90 at any time of the present and the former studies [12]. An ASES Total score of 100 (representing no pain and maximum function) was reached by five of the 11 subjects (45.5%) in the UA-ADRCs group but none of the five subjects (0%) in the corticosteroid group at any time of the present and the former studies [12].

Statistical analysis demonstrated that compared with the subjects in the corticosteroid group, the subjects in the UA-ADRCs group had (i) a significantly higher mean ASES Total score at W24 and W52 post-treatment as well as at SSV (i.e., at 40.6 ± 1.9 months post-treatment), (ii) a significantly higher mean SF-36 Total score at W24 post-treatment, and (iii) a significantly higher mean VAS Pain score at W24 and W52 post-treatment (Fig. 2A-C and Tables S16–S18).

**FIGURE 2.**
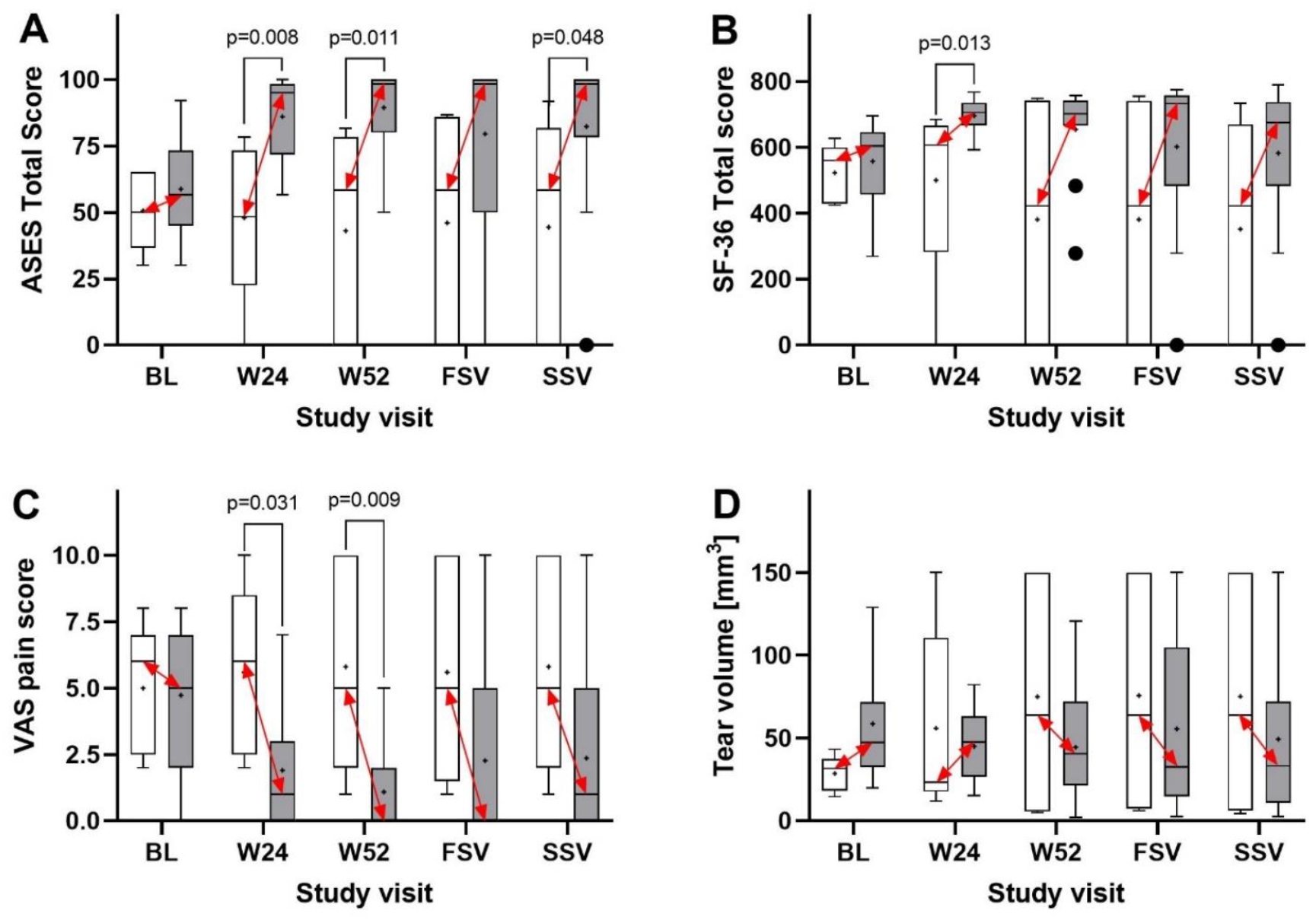
Tukey boxplots of (A) ASES Total score, (B) SF-36 Total score, (C) VAS Pain score (collected together with the ASES score) and (D) tear volume measured on MRIs of subjects treated with injection of either UA-ADRCs (gray bars) or corticosteroid (open bars). The red double-arrows indicate corresponding median values. P-values <0.05 are indicated in (A-C); all p-values are provided in Tables S16–S18. Abbreviations: BL, baseline; W24 / W52, study visits scheduled in the former study [12] at 24 and 52 weeks post-treatment; FSV, first study visit of the present study at 33.2 ± 1.0 (mean ± standard deviation) months post-treatment; SSV, second study visit of the present study at 40.6 ± 1.9 months post-treatment.

### Partial thickness rotator cuff tear size as a function of time after treatment with injection of either UA-ADRCs or corticosteroid

The individual tear size as a function of time post-treatment is shown in Fig. S19; imputation of missing data is indicated in this figure. Statistical analysis demonstrated no significant differences between the subjects in the UA-ADRCs group and the subjects in the corticosteroid group (Fig. 2D and Table S19).

### Detection of hyperintense structures on PD FS T2 coronal MRI scans of the index shoulder at the position of the supraspinatus tendon after injection of UA-ADRCs, but not after injection of corticosteroid, at 24 weeks post-treatment but not at baseline

The PD FS T2 coronal MRI scans of the index shoulder of 10 of 11 subjects (90.9%) in the UA-ADRCs group and none of the subjects (0%) in the corticosteroid group showed hyperintense structures at the position of the supraspinatus tendon at 24 weeks post-treatment but not at baseline. A representative example is shown in Fig. 3; all PD FS T2 coronal MRI scans of Subjects A1-A11 (injection of UA-ADRCs) and Subjects C1-C4 (injection of corticosteroid) are provided in Figs S1–S15. No MRI scans of Subject C5 are shown because this subject was not enrolled in the present study, and the study protocol did not allow to re-assess the MRI scans of this subject generated during the former study [12].

**FIGURE 3.**
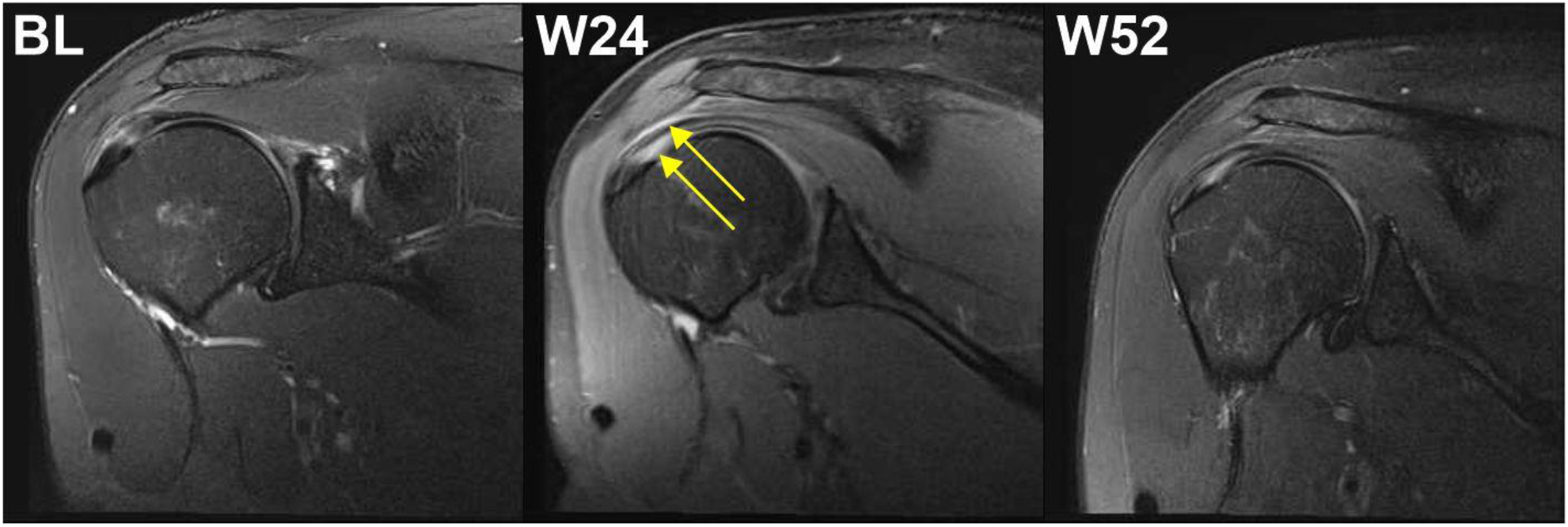
Proton density weighted, fat saturated, T2-weighted, coronal MRI scans of the index shoulder of Subject A4 (injection of UA-ADRCs), showing hyperintense structures at the position of the supraspinatus tendon at 24 weeks post-treatment (yellow arrows) but not at baseline. Abbreviations: BL, baseline; W24 / W52, study visits scheduled in the former study [12] at 24 and 52 weeks post-treatment.

### No relationship between treatment outcome and baseline data

Figures S20 and S21 show individual ASES Total scores as a function of time post-treatment together with individual data at baseline (ASES Total score, tear volume, age and body mass index, as well as (in case of subjects who were treated with injection of UA-ADRCs) cell yield and cell viability. No relationship between treatment outcome and baseline data was found, including those data characterizing UA-ADRCs that can be collected with a clinical test.

## DISCUSSION

To assess the relevance of the results of the present study in accordance with the current state of knowledge, Table 2 summarizes the essential details of all previously published clinical studies on the management of partial-thickness and full-thickness rotator cuff tears with stem cells [12,29–37]. In most of these studies, stem cells were applied to improve the outcome of surgical treatment. Furthermore, next to the former study [12], treatment of partial-thickness rotator cuff tears was only investigated in three other studies [34–36], and only three other studies were randomized controlled trials [32,36,37]. The mean number of subjects treated with stem cells in the studies listed in Table 2 (excluding the former study [12]) was 19.1 ± 5.0 (mean ± SEM) (median, 13.5; range, 7-45).

**TABLE 2.** Essential details of all published clinical studies on the management of rotator cuff tears with stem cells.

| Reference | [29] | [30] | [31] | [32] | [33] | [34,35*] | [12] | [36] | [37] |
| --- | --- | --- | --- | --- | --- | --- | --- | --- | --- |
| Year of publication | 2012 | 2014 | 2015 | 2016 | 2017 | 2018 | 2020 | 2022 | 2022 |
| RCT | No | No | No | Yes | No | No | Yes | Yes | Yes |
| n (SCs / C) | 14 / - | 45 / - | 8 / - | 8 / 5 | 35 / 35 | 13 / 3+3 | 11 / 5 | 7 / 8 | 23 / 23 |
| Tear type | n.s. | F | F | F | F | P | P | P | F |
| Only SCs | No | No | No | No | No | Yes | Yes | Yes | No |
| Type of SCs | BM | BM | BM | BM | UA-ADRCs | ADSCs | UA-ADRCs | ADSCs | MFF |
| Autologous SCs | Yes | Yes | Yes | Yes | Yes | Yes | Yes | No | Yes |
| Surgery adjunct | Yes | Yes | Yes | Yes | Yes | No | No | No | Yes |
| Arthroscopy | n/a | n/a | n/a | n/a | n/a | Yes | No | No | n/a |
| Stat Plan | No | Yes (b) | No | Yes (w) | Yes (b/w) | Yes (b) | Yes (b/w) | Yes (b/w) | Yes (b/w) |
| Follow-up | M12 | M120 | M6 | M12 | M28** | M6 / M24 | M12 | M24 | M24 |
| Outcome | n/a | n/a | n/a | *** | No | n/a | Yes | No | Yes |

Abbreviations: RCT, randomized controlled trial; n, number of subjects; SCs, stem cells; C, control treatment; n.s., not specified; F, full-thickness rotator cuff tears; P, partial-thickness rotator cuff tears; BM, bone marrow-derived mesenchymal stem cells; UA-ADRCs, fresh, uncultured, unmodified, autologous, adipose-derived regenerative cells; ADSCs, cultured adipose-derived stem cells; MFF, microfragmented fat; n/a, not applicable; Stat Plan, statistical analysis of differences between (b) and/or within (w) the groups to assess the therapeutic outcome achieved; M12 / M120 / M6 / M29 / M24, 12 / 120 / 6 / 29 / 24 months post-treatment; \*, M24 follow-up study of [24]; \*\*, 28.3 ± 3.8 months in the injection group and 28.8 ± 4.2 months in the conventional group; \*\*\*, this study was stopped due to adverse effects observed in both groups; most probably the reason was the use of a xenogenic scaffold (OrthADAPT Boimplant; Synovis Orthopedic & Woundcare Inc., Irvine, CA, USA) in both groups.

With respect to the long-term safety we evidenced that treatment of sPTRCT with injection of UA-ADRCs did not result in serious adverse events by 40.6 ± 1.9 months post-treatment. There were no greater risks connected with injection of UA-ADRCs than those connected with injection of corticosteroid in treatment of sPTRCT. In summary, the results of the present study suggest that the use of UA-ADRCs in subjects with sPTRCT is safe. The only other study listed in Table 2 (excluding the former study [12]) in which UA-ADRCs were applied [33] did not address the safety of the procedure.

Regarding the long-term efficacy, we evidenced that the subjects in the UA-ADRCs group had significantly higher mean ASES Total scores than the subjects in the corticosteroid group at W24 and W52 post-treatment as well as at the second study visit of the present study, significantly higher SF-36 Total scores at W24 post-treatment, and significantly higher VAS-Pain scores at W24 and W52 post-treatment. These findings were in line with the findings of the former study [12] that treatment of sPTRCT with injection of UA-ADRCs leads to improved shoulder function. We hypothesize that the negative outcome (i.e., no significant difference in mean data between the groups) observed for the ASES Total score at the first study visit of the present study as well as the SF-36 Total score and the VAS Pain score at the first and second study visits of the present study were consequent to the small sample size. Using an adequate sample size of n=246 subjects an ongoing pivotal clinical trial is currently testing the hypothesis that treatment of sPTRCT with injection of UA-ADRCs is more effective than treatment of sPTRCT with injection of corticosteroid [38].

We would like to point to the following, additional results of the present and the former studies [12]: (i) The results obtained in the former study [12] after treatment of sPTRCT with injection of corticosteroid were in line with other studies which investigated the efficacy of treating sPTRCT with injection of corticosteroid [39–43] (details in Tables S20 and S21). (ii) Six subjects in the UA-ADRCs group but no subject in the corticosteroid group reached an ASES Total score of 100 over time after treatment (Fig. S16). Among these six subjects in the UA-ADRCs group, five reported an ASES Total score of 100 both at the end of the former study [12] and throughout the present study (Fig. S16). (iii) One subject in the corticosteroid group (Subject C5 in Figs S16–S18) developed a full thickness tear during the former study [12], and another subject in the corticosteroid group (Subject C4 in Figs S16–S18) developed pain in the index shoulder at 1.4 months post-treatment and was treated with another injection of corticosteroid at 7.4 months post-treatment during the former study [12]. In contrast, except for one subject in the UA-ADRCs group (Subject A9 in Figs S16–S18) who reported an accident with involvement of the index shoulder at 1.0 months post-treatment, no subject in the UA-ADRCs group required additional treatment of the index shoulder during the former study [12]. In summary, these results reinforce the general need to individually examine the clinical course after an initial treatment, and to identify all possible interfering influences that could have negatively impacted the success of the therapy under study. Furthermore, these results support our hypothesis that treatment of UA-ADRCs with injection of sPTRCT is effective, and is more effective than treatment of sPTRCT with injection of corticosteroid.

The only other published study to date that investigated treatment of sPTRCT with injection of stem cells without surgery found no benefit of injection of cultured adipose-derived stem cells (ADSCs) for 24 months post-treatment, and the results obtained after injection of ADSCs did not differ from the results obtained after injection of saline [36]. This negative result could have been caused by at least three circumstances: (i) the use of allogeneic cells, with the possible inability of new cells derived from the stem cells to integrate into the host tissue because of immunological incompatibility [14]; (ii) the need for culturing the cells, with the possible reduction of the life span of the cells by shortening the telomeres following repetitive cell divisions, and possible negative effects on the safety of the cells as a medicinal product [13]; and (iii) the selection of a single cell type, with the consequence of limited functionality of the cells [14,44]. All this is prevented by the use of fresh UA-ADRCs in the present and the former studies [12], and may explain the discrepancy between the negative result in [36] and the positive results in the present and the former studies [12].

It is beyond the scope of the present study to provide a comprehensive explanation why selection of stem cells (i.e., the use of cultured ADSCs or cultured MSCs in general) is inferior to the use of fresh UA-ADRCs in treatment of musculoskeletal pathologies. Here we report just three of the most important reasons: (i) unlike cultured ADSCs, fresh UA-ADRCs express those growth factors that are needed to stimulate cultured ADSCs towards tenogenic differentiation in culture [45]; (ii) these growth factors are expressed by M2 macrophages [46–49], and M2 macrophages are contained in the UA-ADRCs used in the present and the former studies [12,20], but are missing in any cultured stem cells; and (iii) M2 macrophages are mainly involved in anti-inflammatory responses [50,51], and the presence of M2 macrophages in UA-ADRCs may explain the very early treatment success observed after treating sPTRCT with UA-ADRCs in the former study [12], which cannot be explained by the formation of new tendon tissue (Fig. S16). In summary, there are a number of possible explanations of the discrepancy between the negative result in [36] and the positive results in the present and the former studies [12] with respect to treatment of sPTRCT with injection of stem cells.

Regarding the analysis of MRIs pre- and post-injection, we found no significant improvement of the mean tear volume over time, nor any significant difference between the results obtained after injection of UA-ADRCs and those obtained after injection of corticosteroid (Table S18). Of note, these findings are not in line with the results related to the long term efficacy (improvement in ASES Total score) outlined above. The main reason for this discrepancy may be the mechanisms of action of UA-ADRCs in tendon repair. Initially one could assume that UA-ADRCs would mainly fill the gap in the tendon tissue caused by a partial-thickness tear. However, the location of the hyperintense structures in PD FS T2 MRI scans at the position of the supraspinatus tendon present at 24 weeks post-treatment but not at baseline in 10 of the 11 subjects in the UA-ADRCs group (Figs S1–S11) and none of the subjects in the corticosteroid group (Figs S12–S15) indicate that this may not be the case. Rather, these hyperintense structures in PD FS T2 MRI scans may indicate formation of new tendon tissue following injection of UA-ADRCs in a different location than the original tear, possibly primarily following individual biomechanical requirements. This may explain why subjects who are suffering from sPTRCT experience fast (the former study [12]) and lasting (the present study) recovery from pain and impaired function without disappearance of the rotator cuff tears on MRI scans even at 41 months post-treatment.

The presence of hyperintense structures in PD FS T2 MRI scans at the position of a tendon with partial-thickness tear a few months after injection of UA-ADRCs has only been reported in a recent single case report [52]. Without additional investigations, it is unclear whether these hyperintense structures in PD FS T2 MRI scans indeed represent formation of new tendon tissue. These investigations must be performed on biopsies of tendons with partial-thickness tear that were treated with injection of UA-ADRCs. On the other hand, there are two indications supporting the hypothesis that these hyperintense structures in PD FS T2 MRI scans indeed represent formation of new tendon tissue: (i) the analysis of the biopsy reported in the recent case report [52] indicated newly formed tendon tissue which did not resemble scar tissue (the biopsy was taken at the position of the hyperintense structure found in the corresponding MRI scans ten weeks post-treatment); and (ii) this biopsy showed a dense network of newly formed microvessels next to the position of newly formed tendon tissue [52]. Blood flow in these newly formed microvessels may indeed explain the occurrence of hyperintense structures in PD FS T2 MRI scans after treatment of sPTRCT with injection of UA-ADRCs. Furthermore, the full or partial disappearance of these hyperintense structures in PD FS T2 MRI scans at 52 weeks post-treatment (Figs S1–S11) may indicate that tendon regeneration was complete, or almost complete, at this time.

In summary, the results of the present study indicate that treatment success after treating sPTRCT with UA-ADRCs cannot be assessed using measurements of tear volume on MRI scans. On the other hand, PD FS T2 MRI scans taken a few months after treatment of sPTRCT with injection of UA-ADRCs may allow to “watch the UA-ADRCs at work”. The latter finding may inform researchers about optimal times for taking biopsies in future research into the mechanisms of action of UA-ADRCs in tendon repair.

Based on the outcome of the analysis shown in Fig. S20 we hypothesize that individual treatment success after treating sPTRCT with injection of UA-ADRCs cannot be predicted based on the following, individual values at baseline: ASES Total score, tear volume, age and BMI, as well as on the cell yield and cell viability of the final cell suspension. This finding is important because it may render individual bedside testing of the final cell suspension in clinical use of UA-ADRCs irrelevant.

It is currently unknown whether individual treatment success after treating sPTRCT with injection of UA-ADRCs can be predicted using the colony forming unit (CFU) assay [15] and/or determination of cell surface markers using fluorescence-activated cell scanning [20]. In any case, these analyses take between several days (determination of surface markers) and more than two weeks (CFU assay). Thus, they are not suitable for clinical testing of the final cell suspension in clinical use of UA-ADRCs.

## Limitations

The limitations of the present study are the same as the limitations of the former study [12]: only a small sample of subjects suffering from sPTRCT was investigated, only a limited number of clinical examination methods was applied, no power analysis was carried out, and neither the subjects nor the physicians who performed treatment and the assessors who performed baseline and follow-up examinations were blinded (only the physicians who analyzed the MRI scans were blinded). We believe that the ongoing clinical trial [21] will demonstrate with sufficient statistical power that treatment of sPTRCT with injection of UA-ADRCs is more effective than treatment of sPTRCT with injection of corticosteroid.

## Conclusions

The present investigation further supports treatment of sPTRCT with injection of UA-ADRCs. Once this therapy is approved in the US, clinicians should consider injection of UA-ADRCs instead of injection of corticosteroids. In the long run treatment of sPTRCT with injection of UA-ADRCs may delay or even prevent surgical treatment of sPTRCT.

## Data Availability

All data produced in the present study are available upon reasonable request to the authors.

## Availability of data and materials

The datasets used and analyzed during the current study are available from the corresponding author on reasonable request, taking into account any confidentiality.

## Authors’ contributions

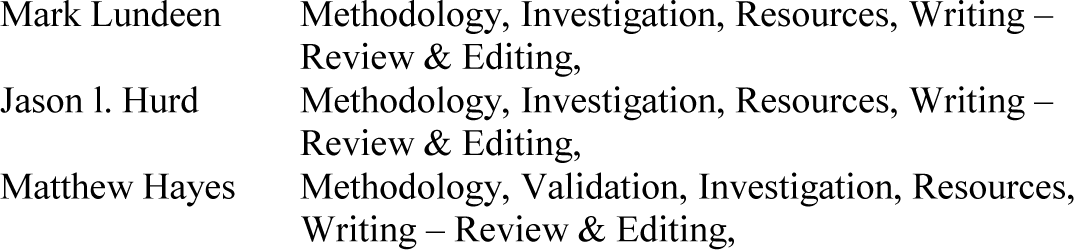
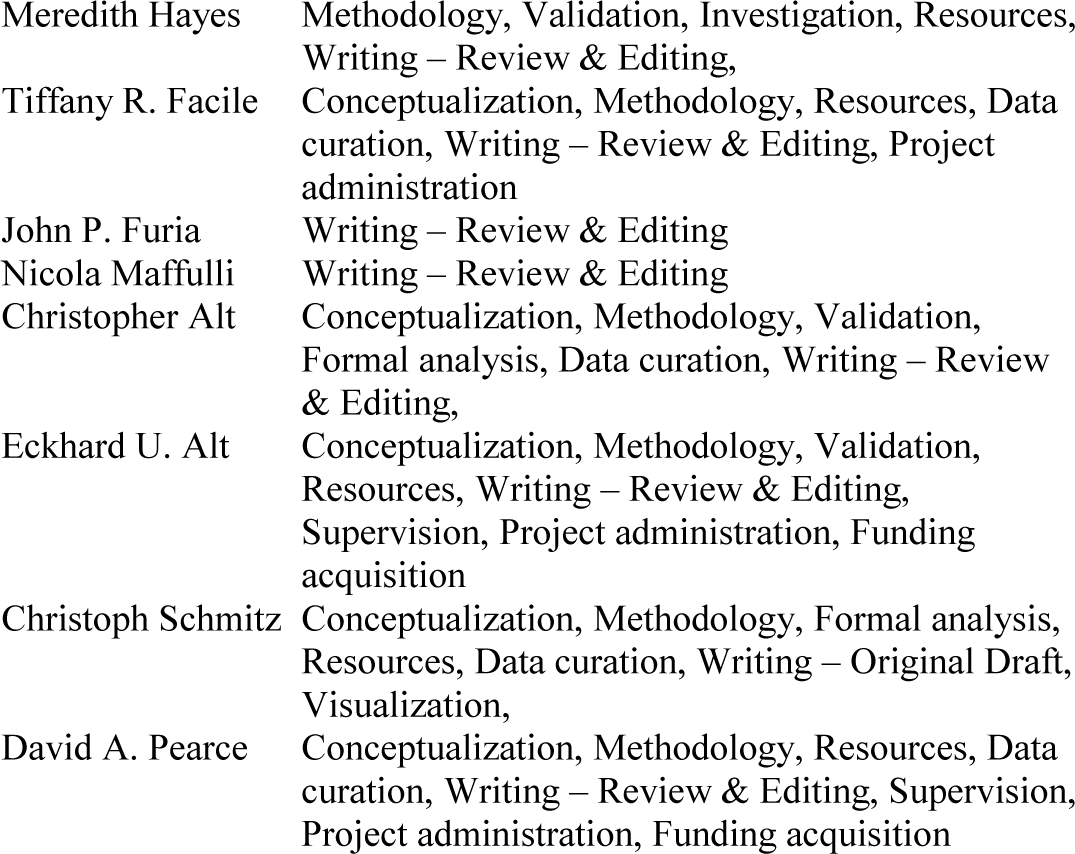

## Consent for publication

We have obtained consent for publication.

## Appendix 1

Note: throughout this document the term “former study” refers to the following study:

Hurd JL, Facile TR, Weiss J, et al. Safety and efficacy of treating symptomatic, partial-thickness rotator cuff tears with fresh, uncultured, unmodified, autologous, adipose-derived regenerative cells (UA-ADRCs) isolated at the point of care: a prospective, randomized, controlled first-in-human pilot study. *J Orthop Surg Res* 2020;**15**:122.

### Part 1 Original MRI scans

**FIGURE S1.**
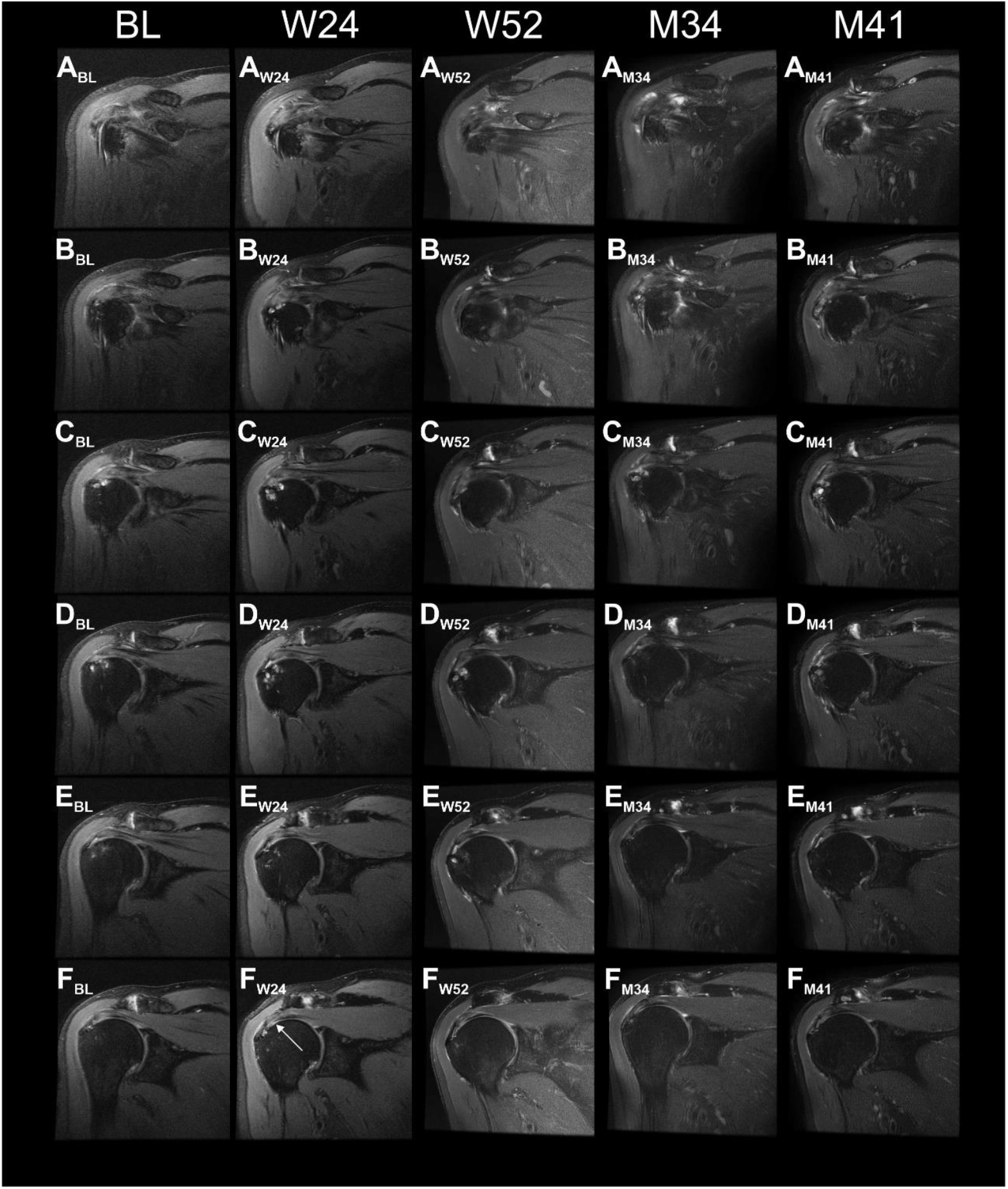
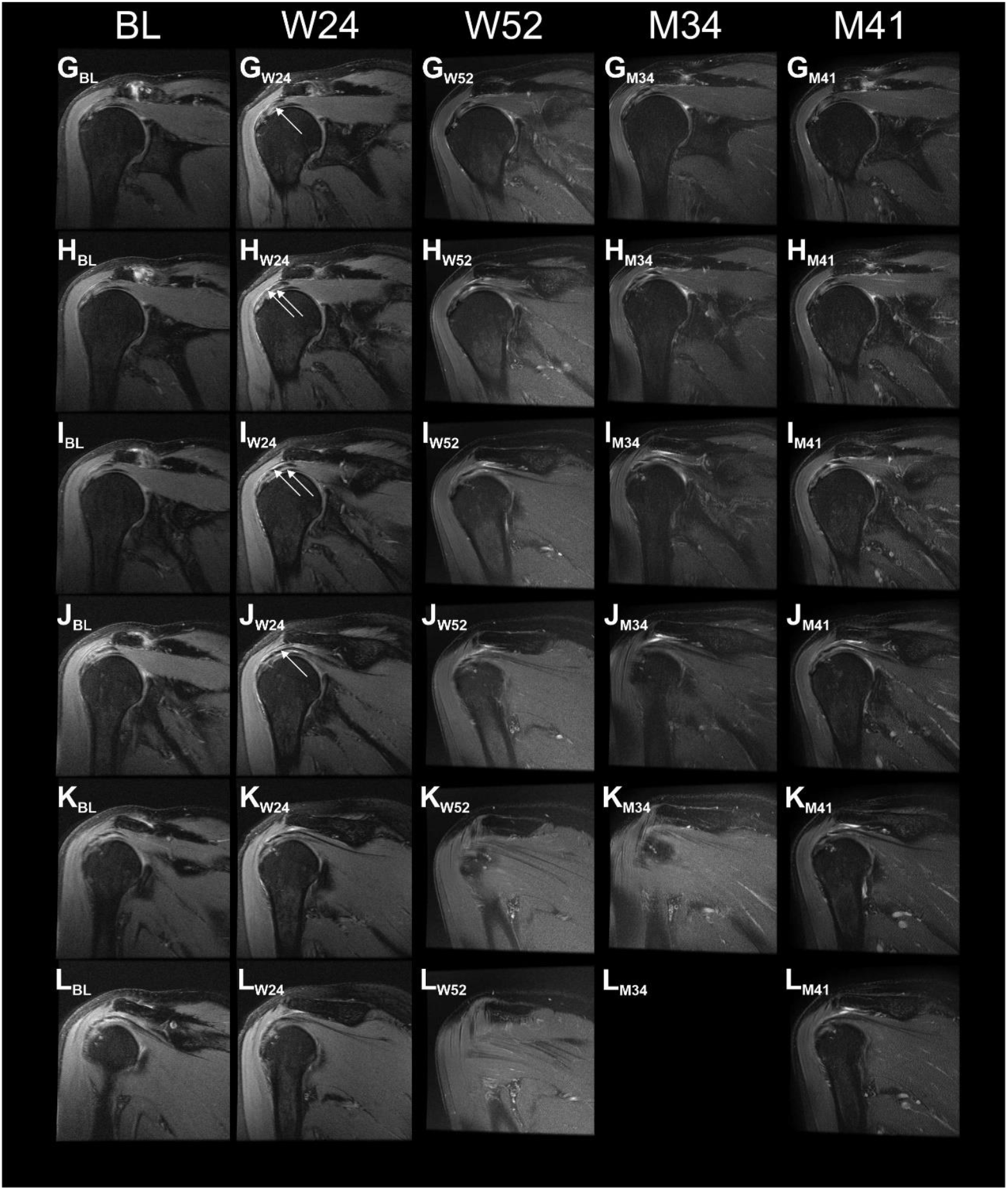
Proton density weighted, fat saturated, T2-weighted, coronal magnetic resonance imaging (MRI) scans of the index shoulder of Subject A1 treated with injection of UA-ADRCs, generated during the present and the former studies. Panels A-L show the same (or nearly the same) image planes at different times, with Panels A showing the most ventral image plane and Panels L the most dorsal image plane. The arrows in Panels F_W24_, G_W24_, H_W24_, I_W24_ and J_W24_ indicate a hyperintense structure at the position of the supraspinatus tendon that was found at 24 weeks post-treatment but not at baseline. Abbreviations: BL, baseline; W24 / W52, 24 / 52 weeks post-treatment; M34 / M41, 34 / 41 months post-treatment.

**FIGURE S2.**
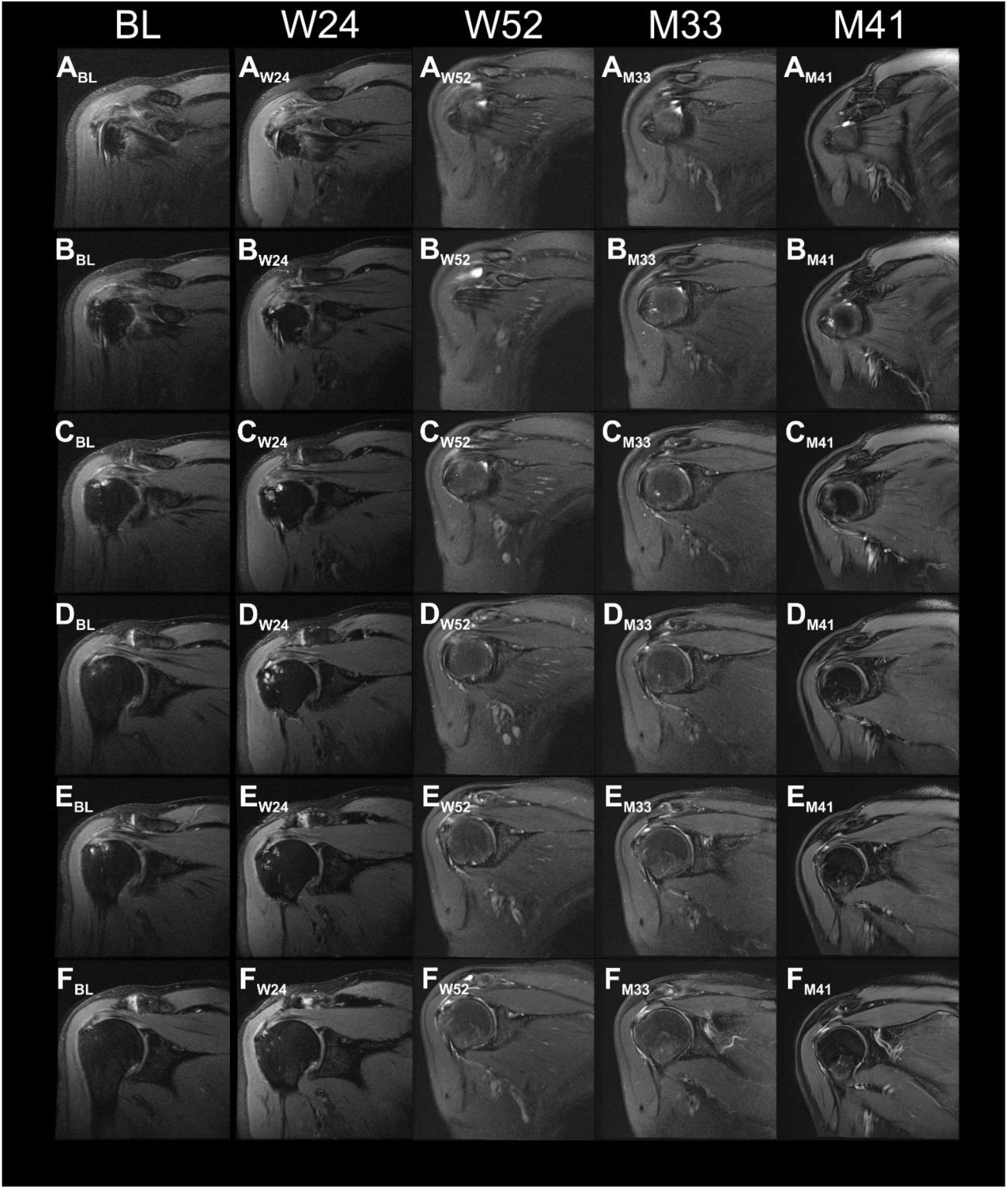
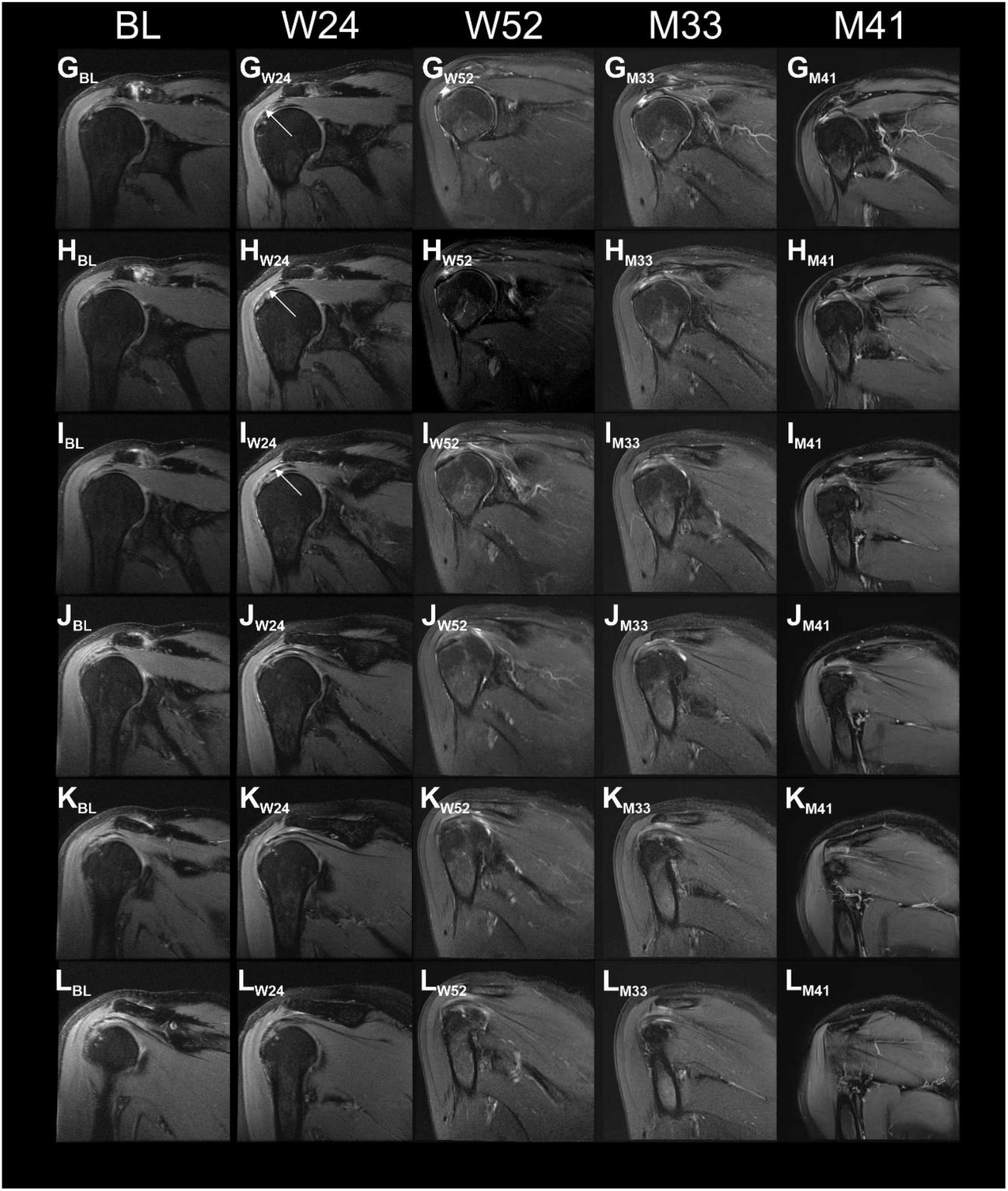
Proton density weighted, fat saturated, T2-weighted, coronal magnetic resonance imaging (MRI) scans of the index shoulder of Subject A2 treated with injection of UA-ADRCs, generated during the present and the former studies. Panels A-L show the same (or nearly the same) image planes at different times, with Panels A showing the most ventral image plane and Panels L the most dorsal image plane. The arrows in Panels G_W24_, H_W24_ and I_W24_ indicate a hyperintense structure at the position of the supraspinatus tendon that was found at 24 weeks post-treatment but not at baseline. Abbreviations: BL, baseline; W24 / W52, 24 / 52 weeks post-treatment; M33 / M41, 33 / 41 months post-treatment.

**FIGURE S3.**
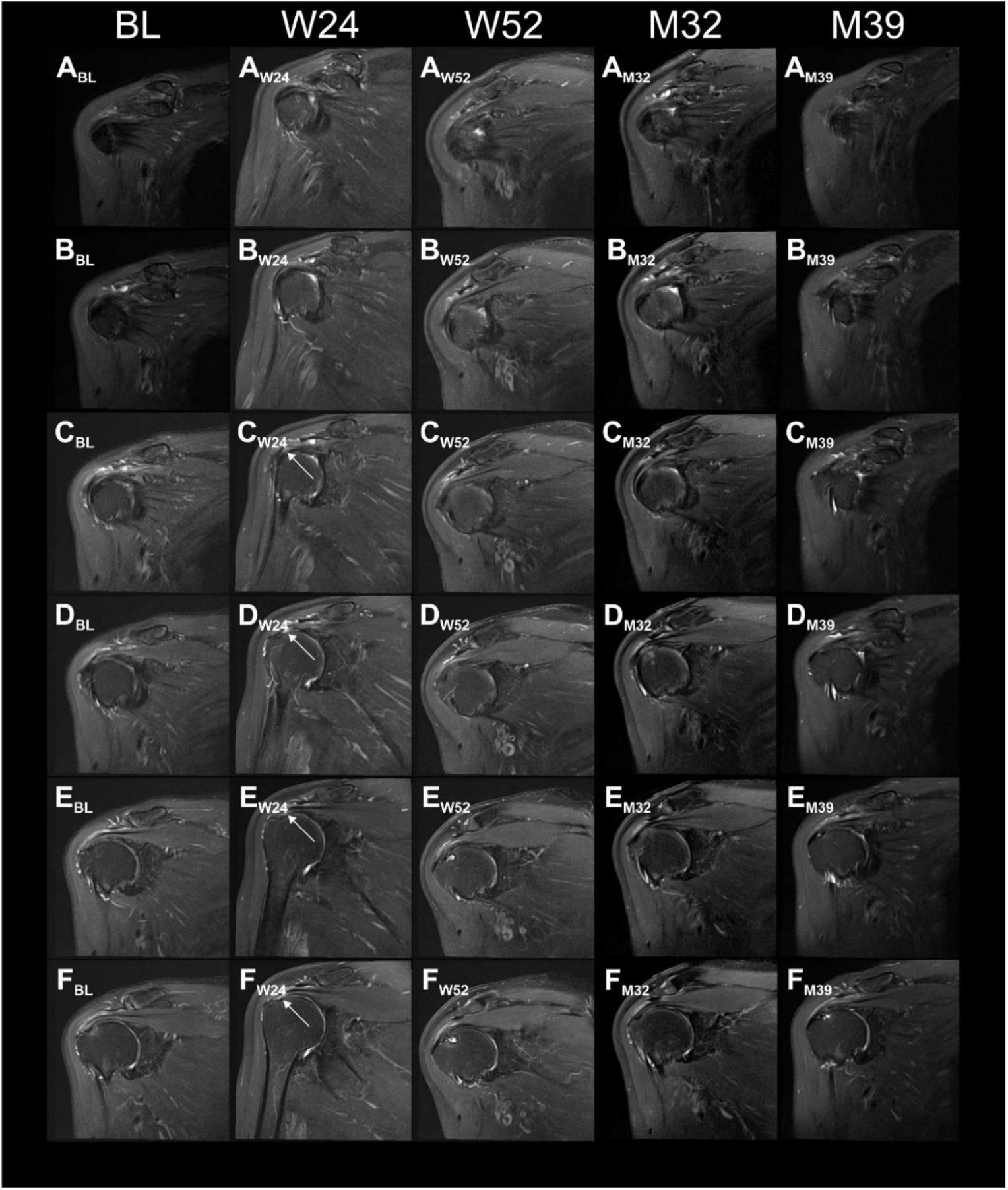
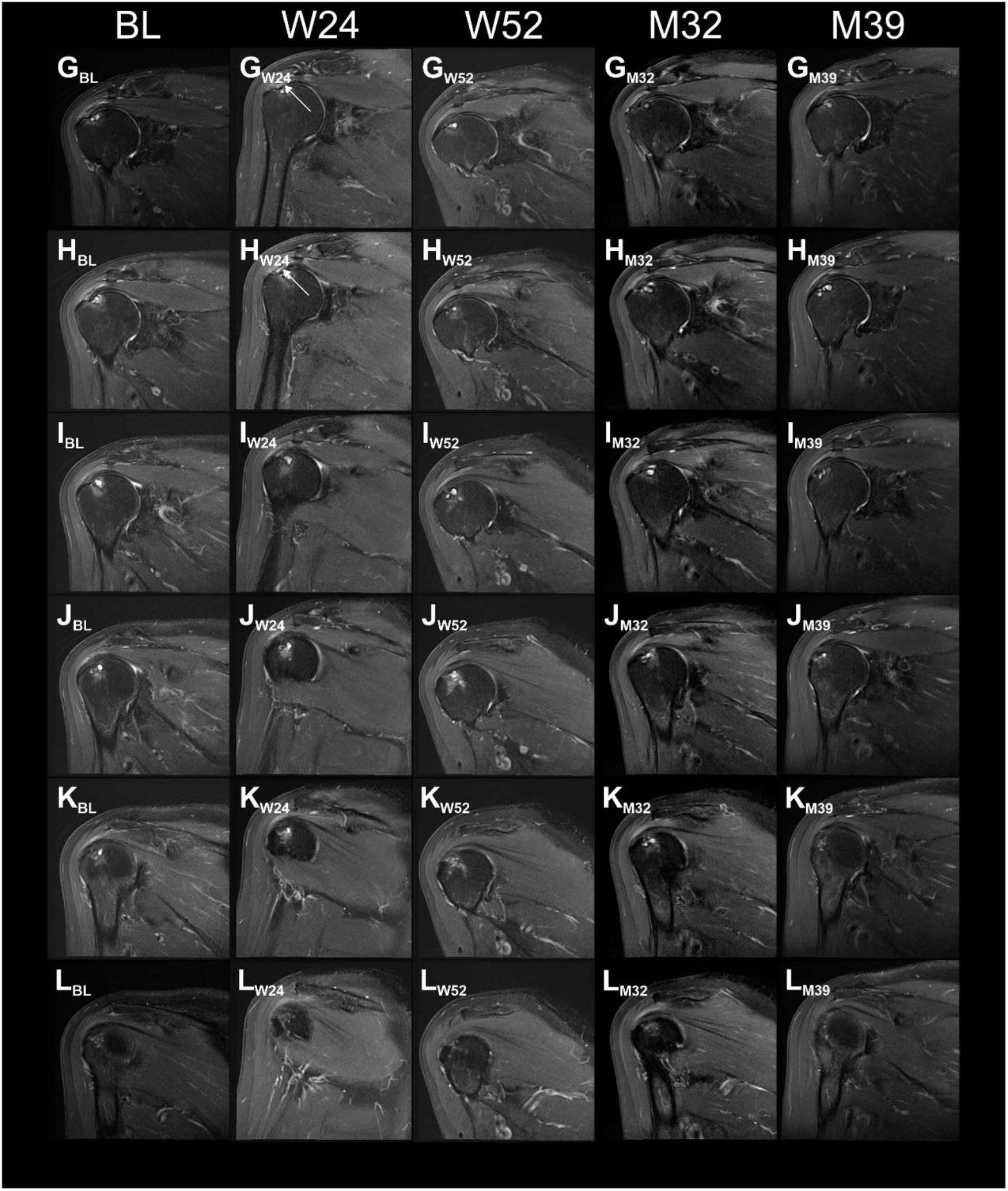
Proton density weighted, fat saturated, T2-weighted, coronal magnetic resonance imaging (MRI) scans of the index shoulder of Subject A3 treated with injection of UA-ADRCs, generated during the present and the former studies. Panels A-L show the same (or nearly the same) image planes at different times, with Panels A showing the most ventral image plane and Panels L the most dorsal image plane. The arrows in Panels C_W24_, D_W24_, E_W24_, F_W24_, G_W24_ and H_W24_ indicate a hyperintense structure at the position of the supraspinatus tendon that was found at 24 weeks post-treatment but not at baseline. Abbreviations: BL, baseline; W24 / W52, 24 / 52 weeks post-treatment; M32 / M39, 32 / 39 months post-treatment.

**FIGURE S4.**
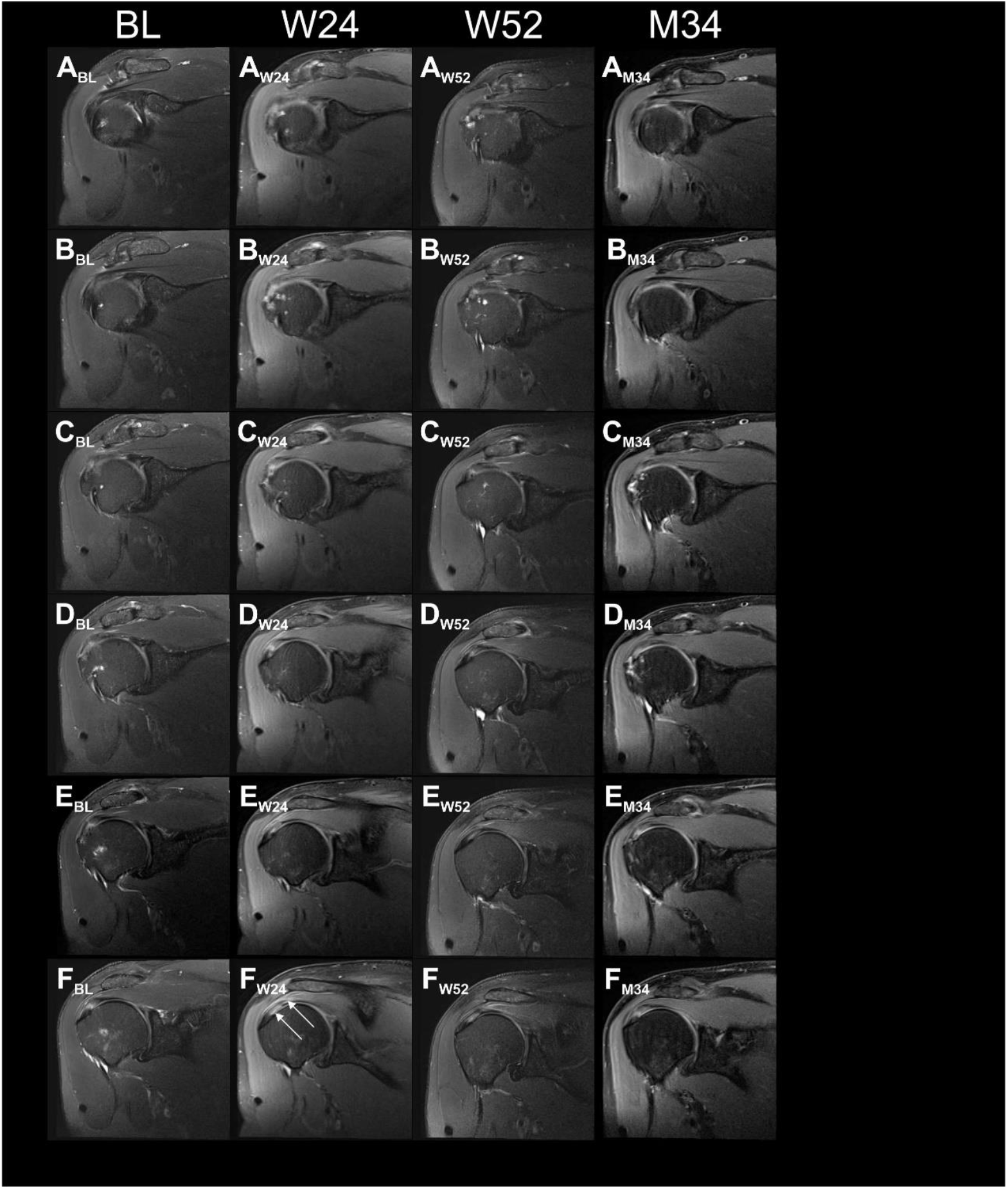
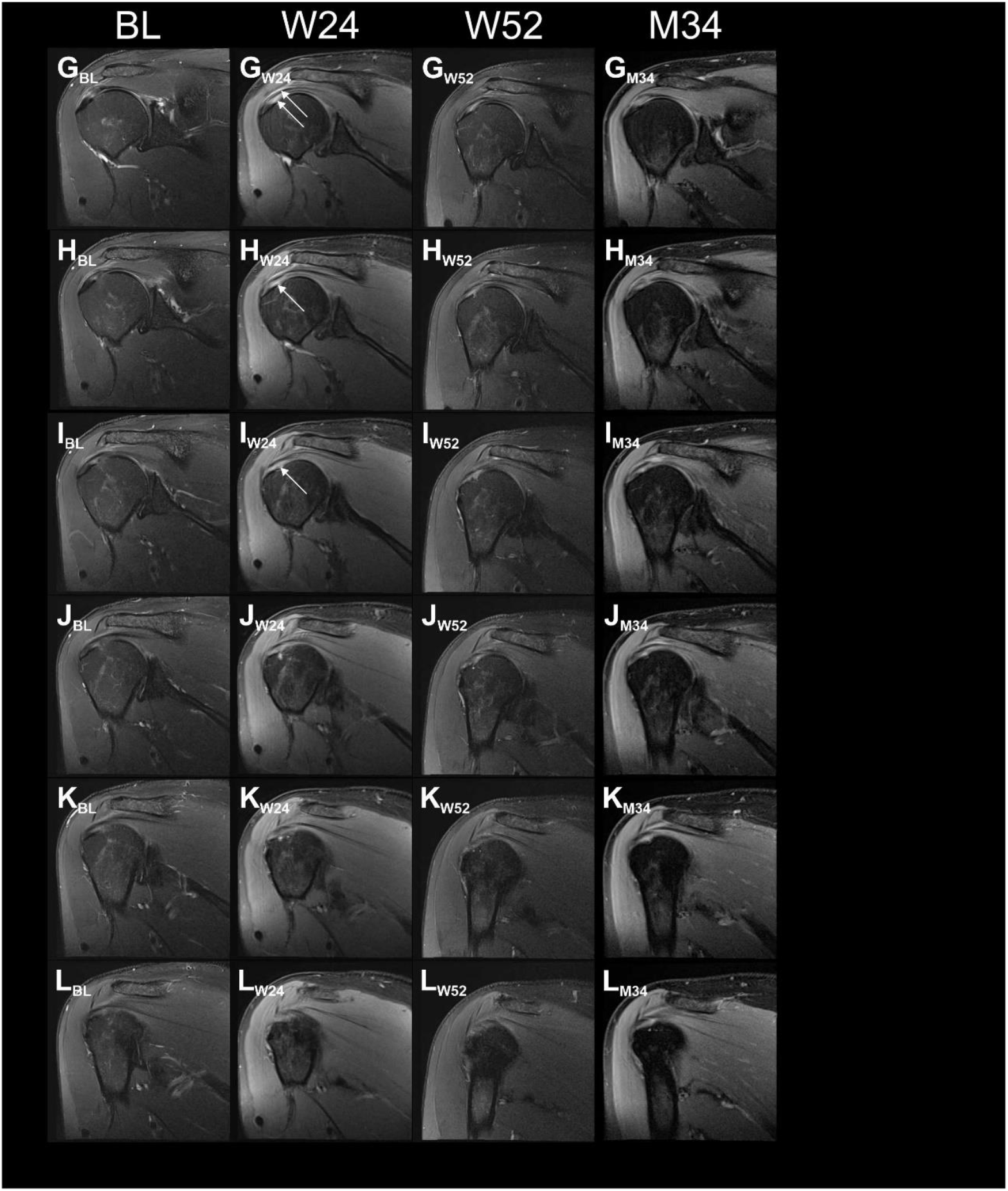
Proton density weighted, fat saturated, T2-weighted, coronal magnetic resonance imaging (MRI) scans of the index shoulder of Subject A4 treated with injection of UA-ADRCs, generated during the present and the former studies. Panels A-L show the same (or nearly the same) image planes at different times, with Panels A showing the most ventral image plane and Panels L the most dorsal image plane. The arrows in Panels F_W24_, G_W24_, H_W24_ and I_W24_ indicate a hyperintense structure at the position of the supraspinatus tendon that was found at 24 weeks post-treatment but not at baseline. Abbreviations: BL, baseline; W24 / W52, 24 / 52 weeks post-treatment; M34, 34 months post-treatment (no MRI was performed during the second visit of Subject A7).

**FIGURE S5.**
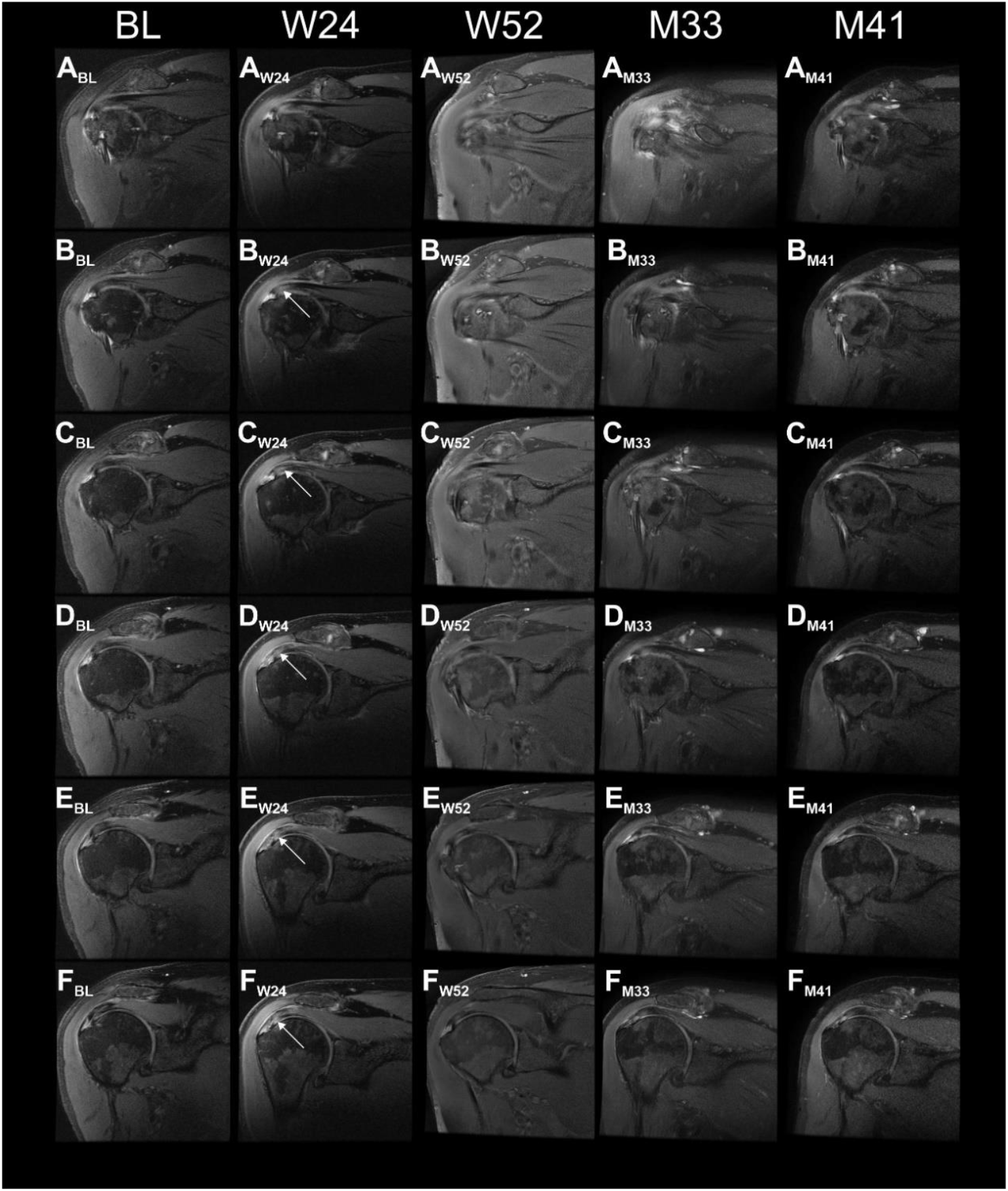
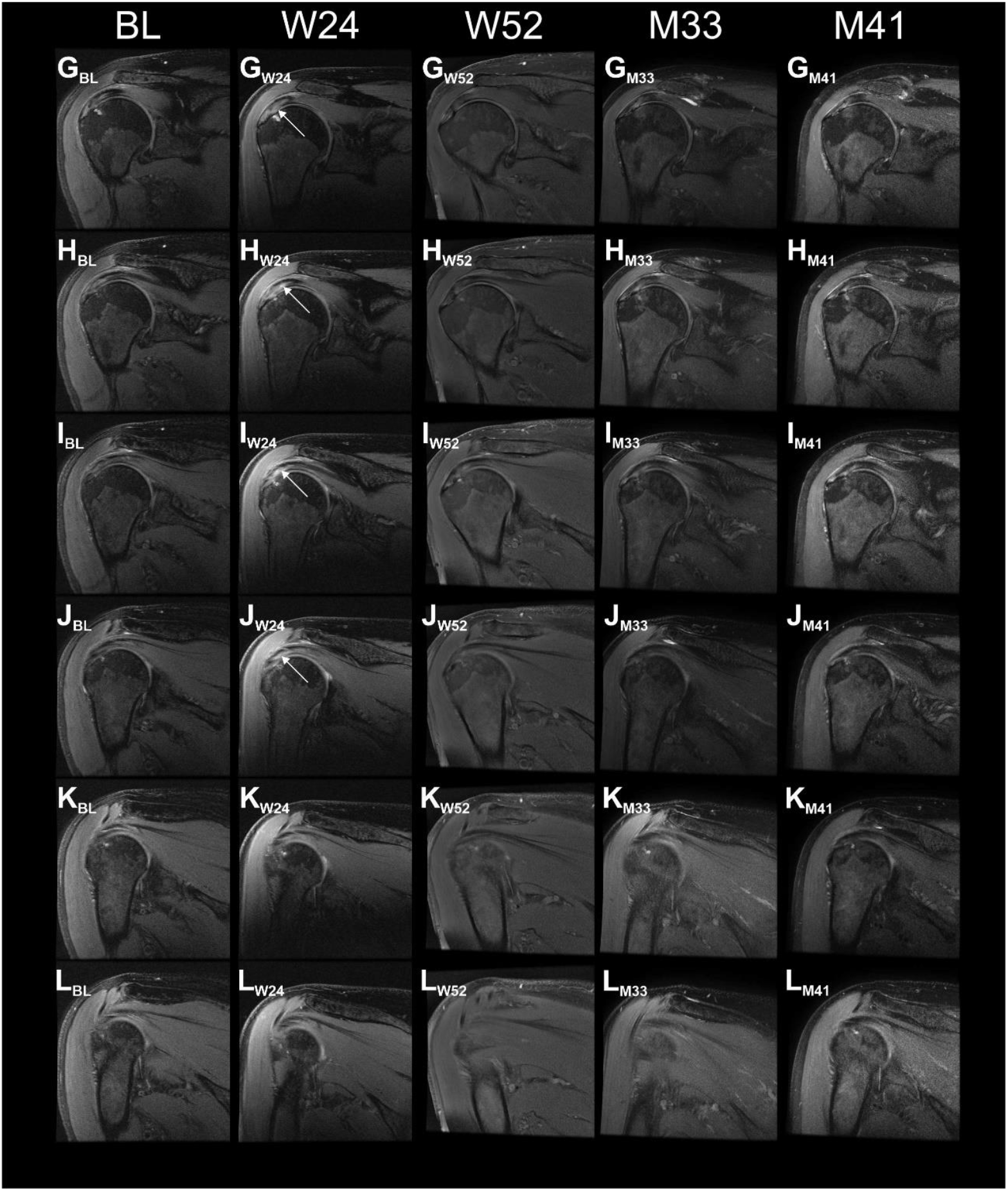
Proton density weighted, fat saturated, T2-weighted, coronal magnetic resonance imaging (MRI) scans of the index shoulder of Subject A5 treated with injection of UA-ADRCs, generated during the present and the former studies. Panels A-L show the same (or nearly the same) image planes at different times, with Panels A showing the most ventral image plane and Panels L the most dorsal image plane. The arrows in Panels B_W24_, C_W24_, D_W24_, E_W24_, F_W24_, G_W24_ H_W24_, I_W24_ and J_W24_ indicate a hyperintense structure at the position of the supraspinatus tendon that was found at 24 weeks post-treatment but not at baseline. Abbreviations: BL, baseline; W24 / W52, 24 / 52 weeks post-treatment; M33 / M41, 33 / 41 months post-treatment.

**FIGURE S6.**
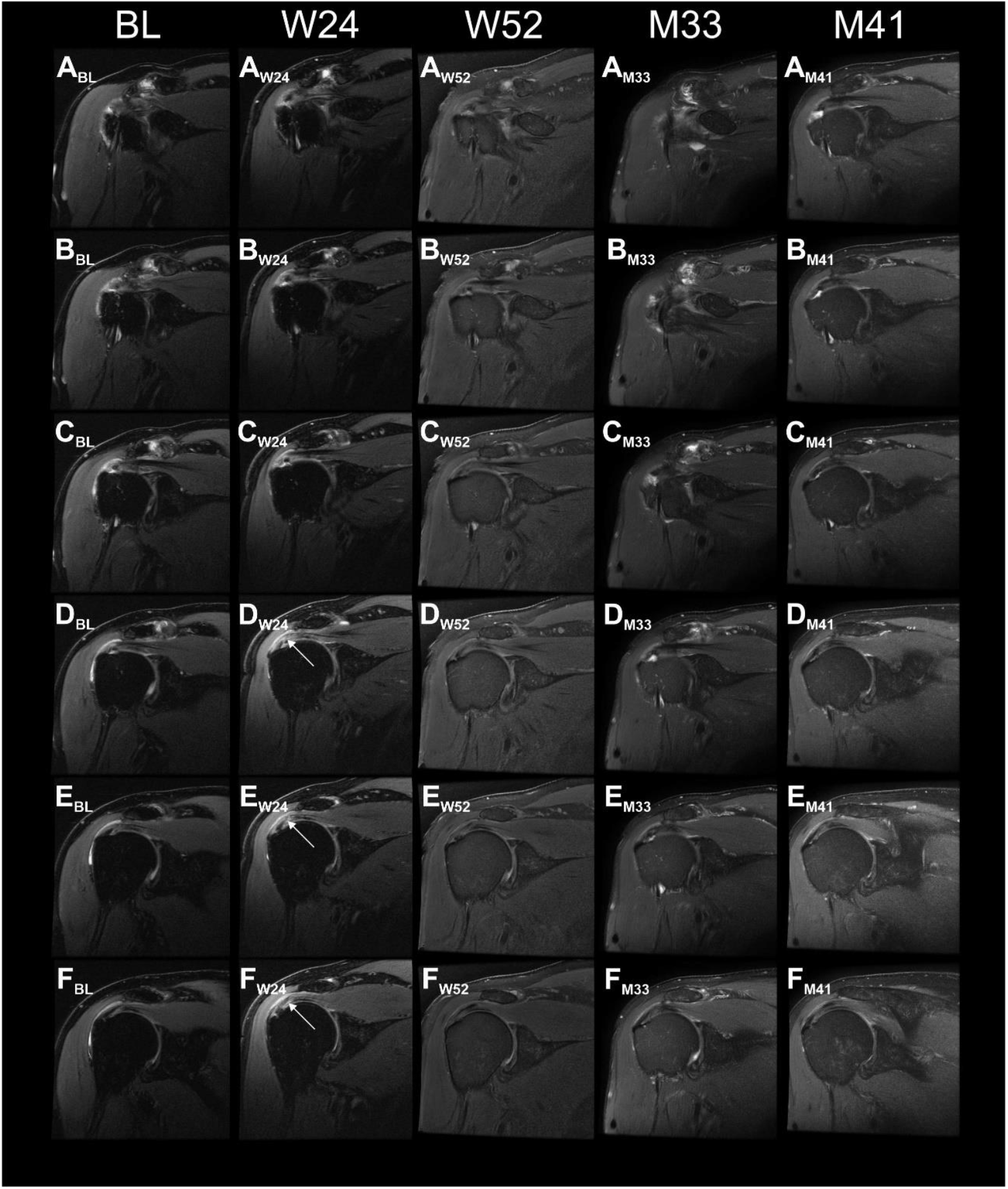
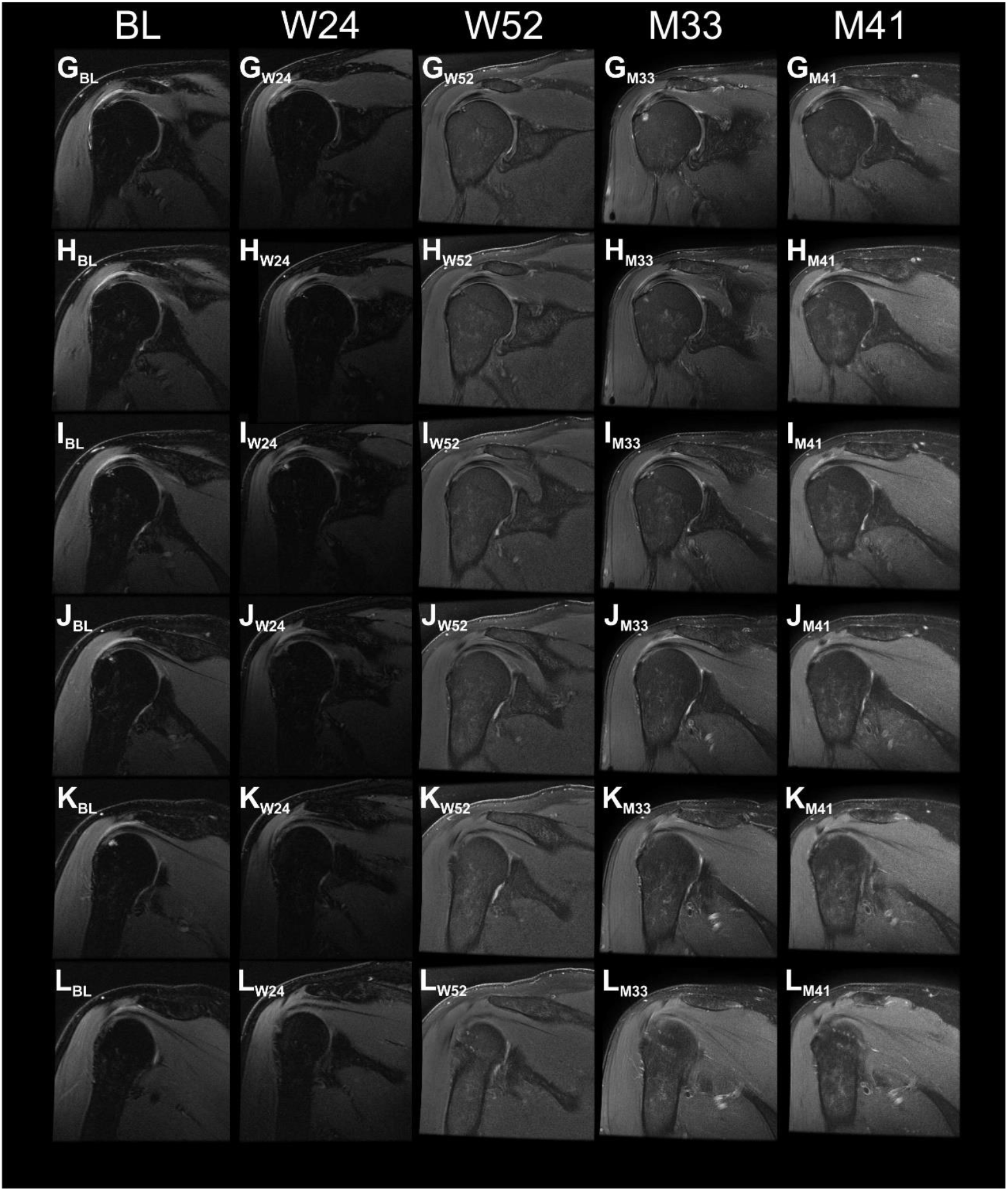
Proton density weighted, fat saturated, T2-weighted, coronal magnetic resonance imaging (MRI) scans of the index shoulder of Subject A6 treated with injection of UA-ADRCs, generated during the present and the former studies. Panels A-L show the same (or nearly the same) image planes at different times, with Panels A showing the most ventral image plane and Panels L the most dorsal image plane. The arrows in Panels D_W24_, E_W24_ and F_W24_ indicate a hyperintense structure at the position of the supraspinatus tendon that was found at 24 weeks post-treatment but not at baseline. Abbreviations: BL, baseline; W24 / W52, 24 / 52 weeks post-treatment; M33 / M41, 33 / 41 months post-treatment.

**FIGURE S7.**
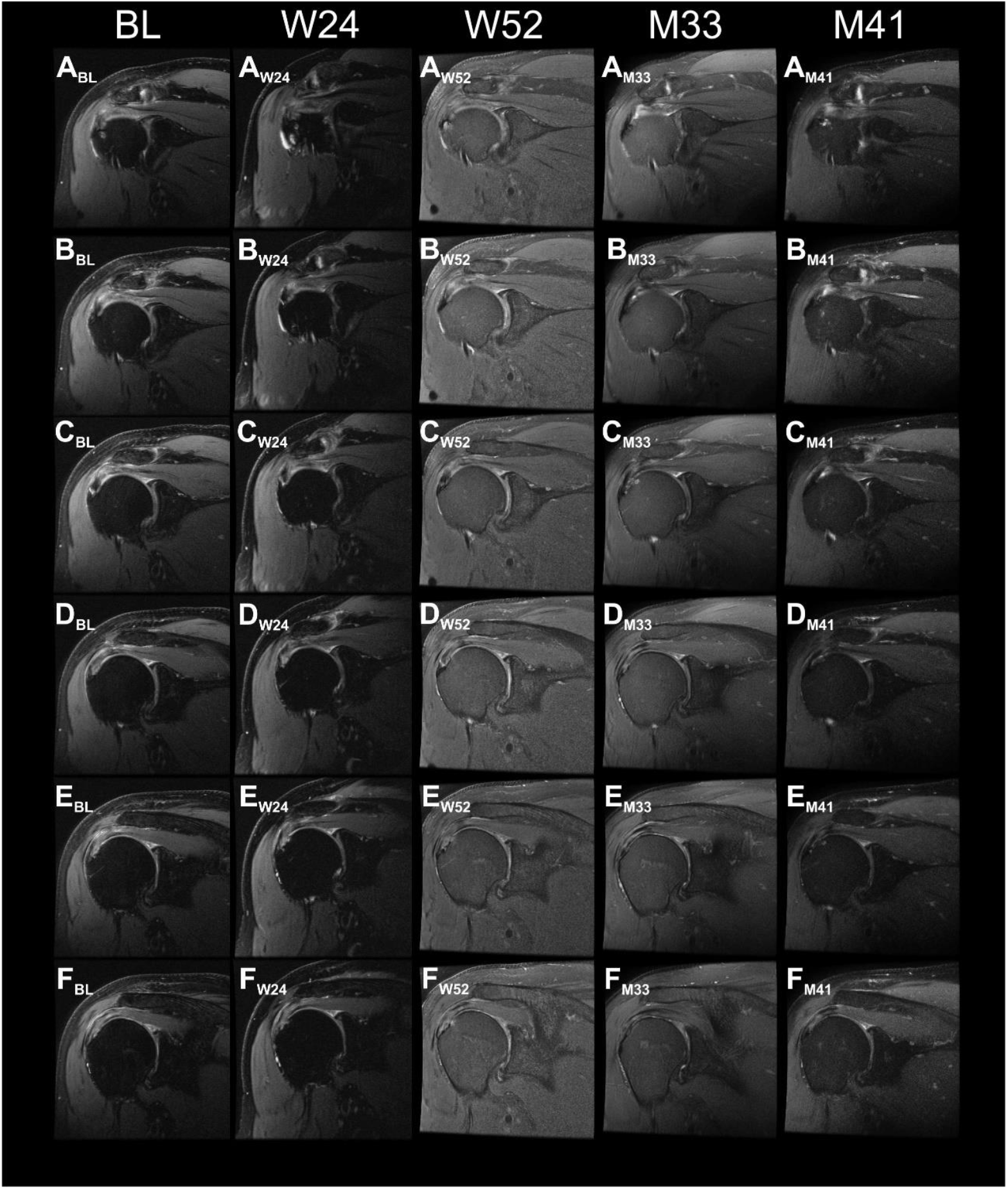
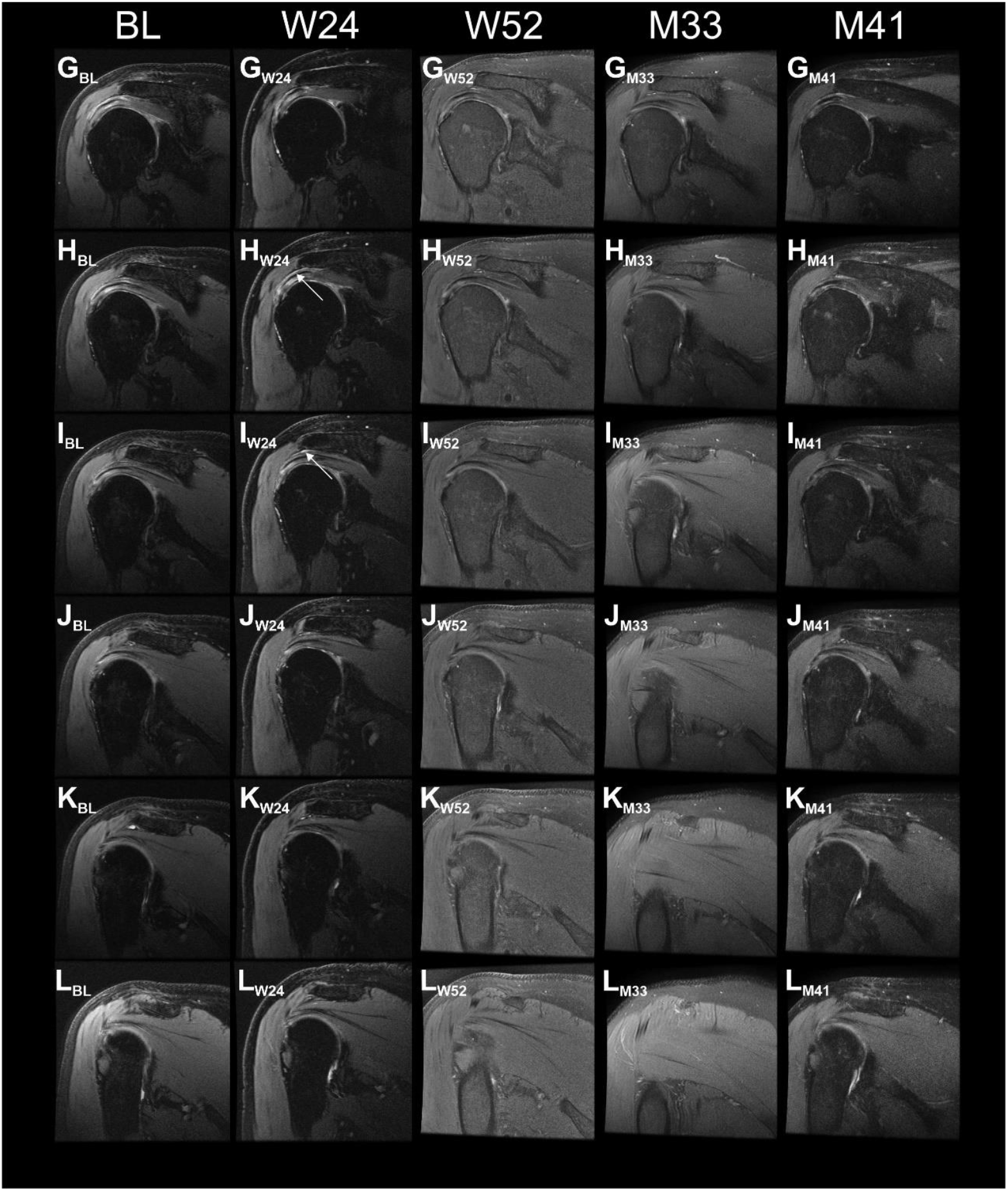
Proton density weighted, fat saturated, T2-weighted, coronal magnetic resonance imaging (MRI) scans of the index shoulder of Subject A7 treated with injection of UA-ADRCs, generated during the present and the former studies. Panels A-L show the same (or nearly the same) image planes at different times, with Panels A showing the most ventral image plane and Panels L the most dorsal image plane. The arrows in Panels H_W24_ and I_W24_ indicate a hyperintense structure at the position of the supraspinatus tendon that was found at 24 weeks post-treatment but not at baseline. Abbreviations: BL, baseline; W24 / W52, 24 / 52 weeks post-treatment; M33 / M41, 33 / 41 months post-treatment.

**FIGURE S8.**
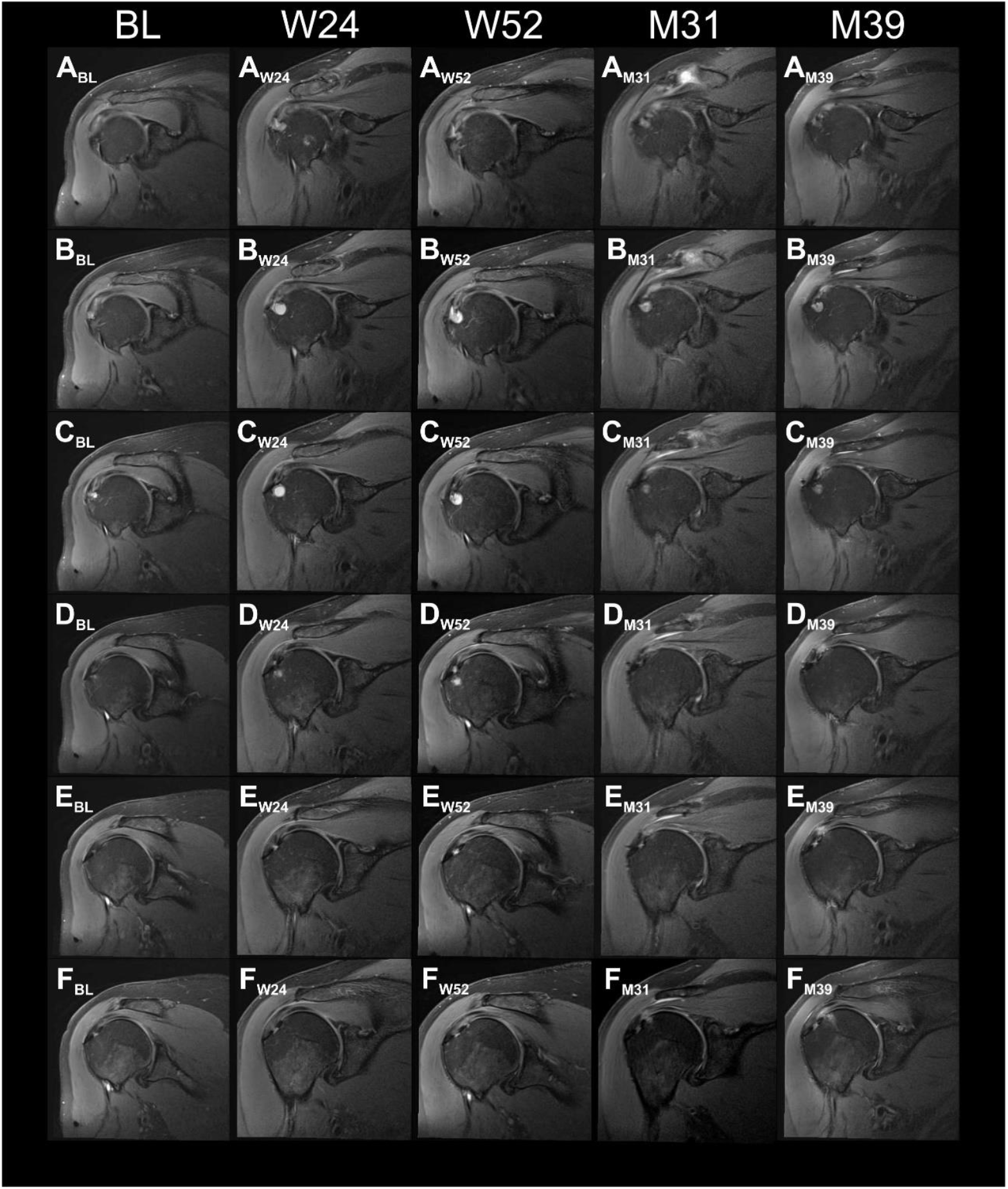
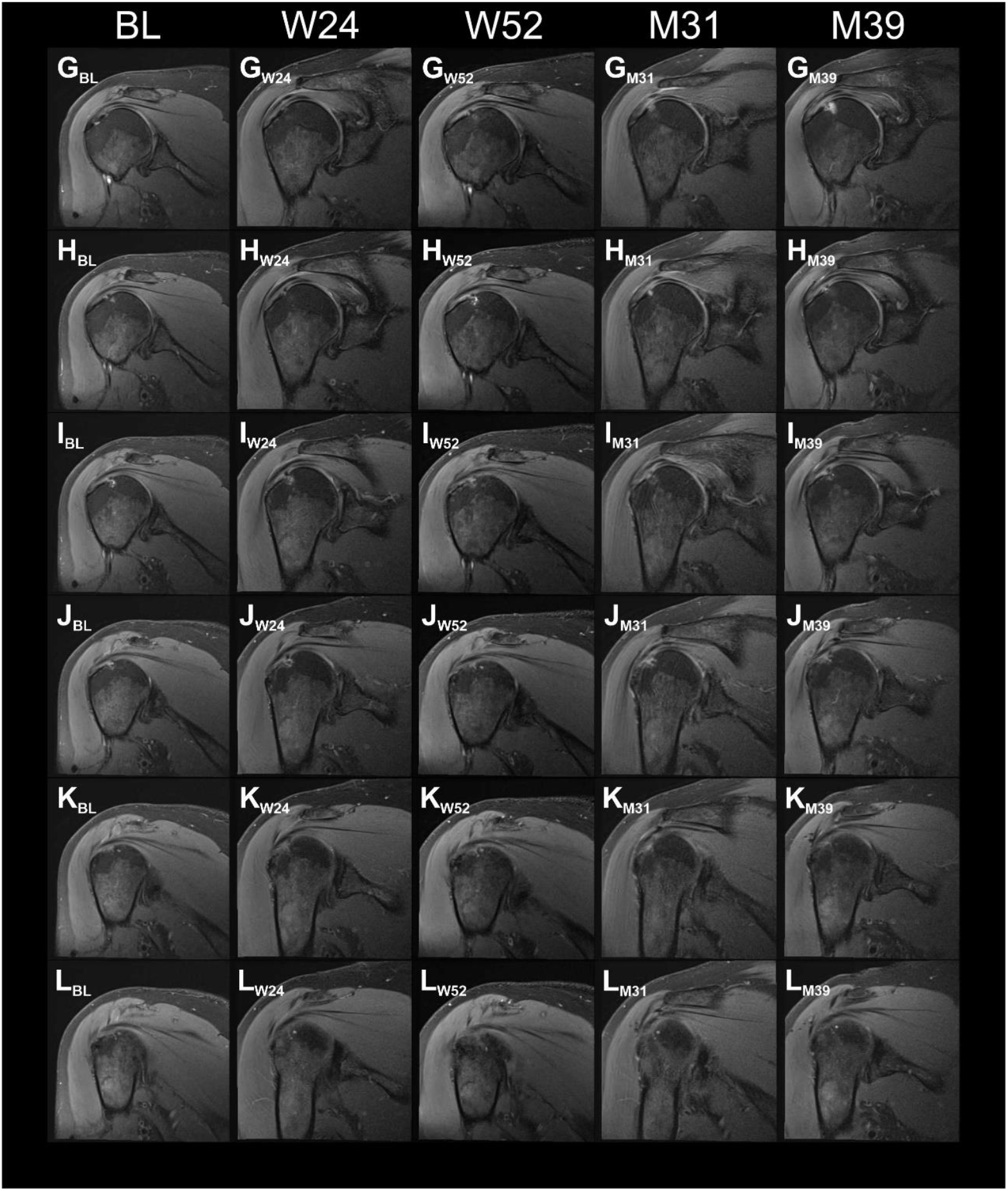
Proton density weighted, fat saturated, T2-weighted, coronal magnetic resonance imaging (MRI) scans of the index shoulder of Subject A8 treated with injection of UA-ADRCs, generated during the present and the former studies. Panels A-L show the same (or nearly the same) image planes at different times, with Panels A showing the most ventral image plane and Panels L the most dorsal image plane. Abbreviations: BL, baseline; W24 / W52, 24 / 52 weeks post-treatment; M33 / M41, 31 / 39 months post-treatment.

**FIGURE S9.**
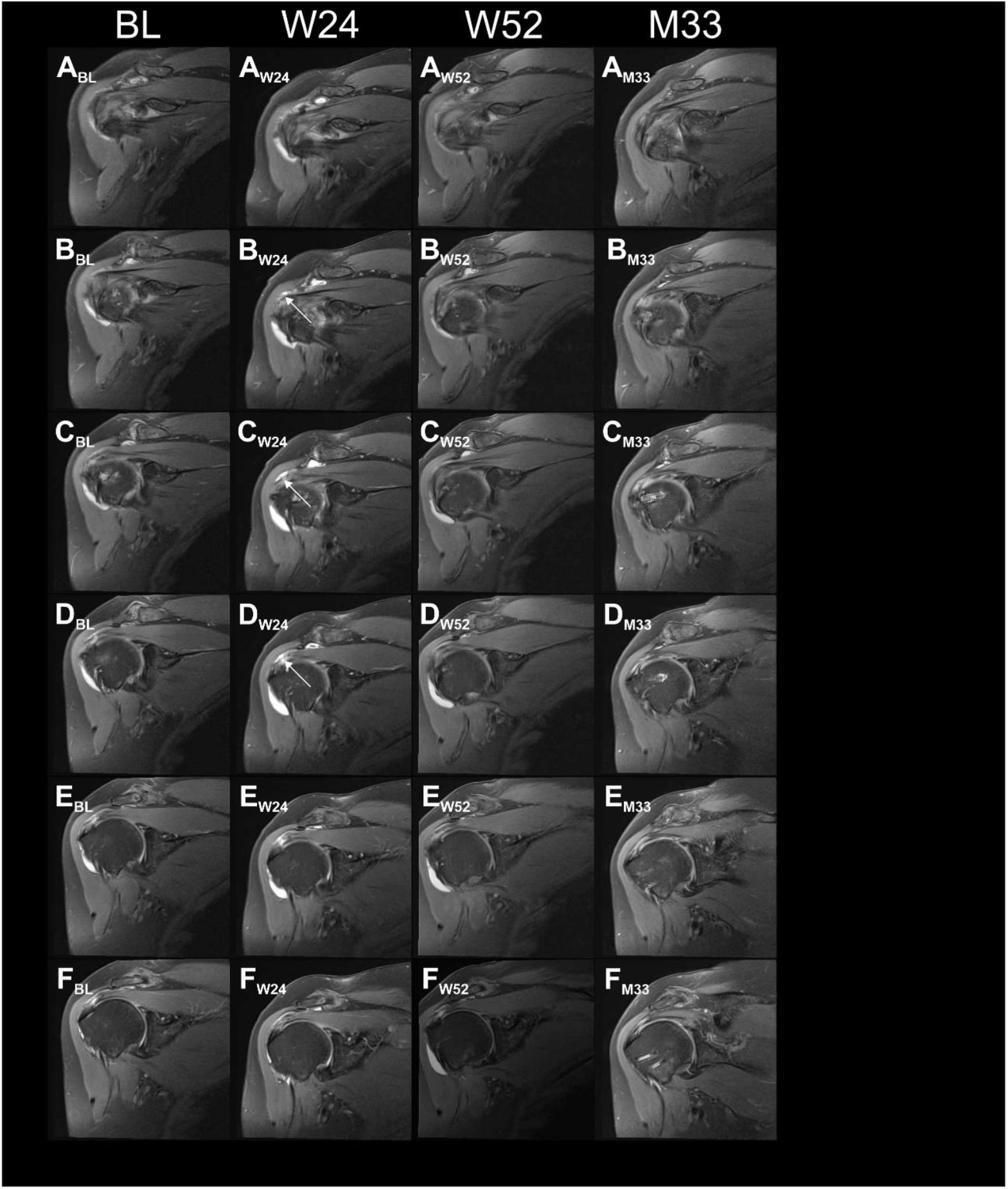
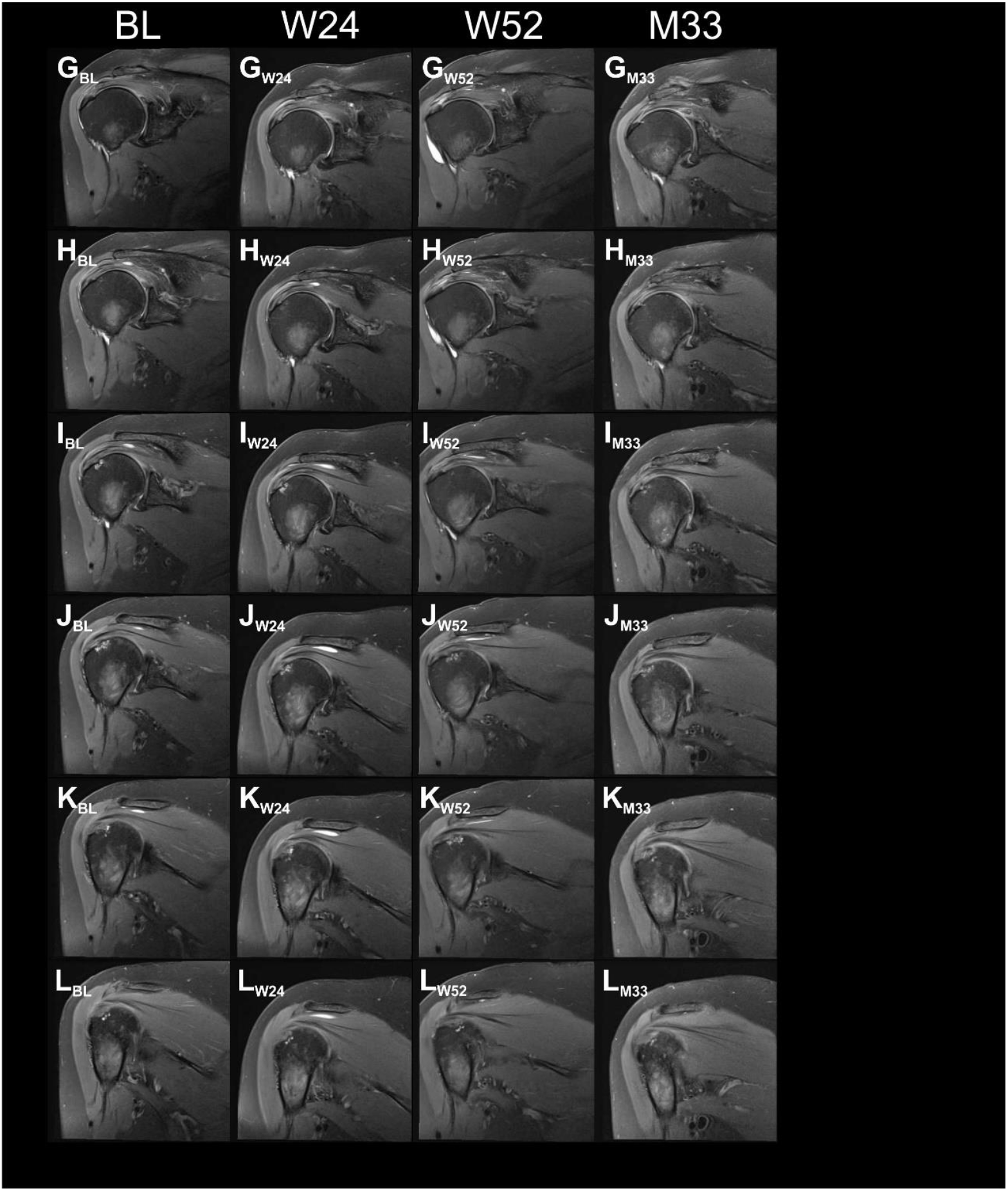
Proton density weighted, fat saturated, T2-weighted, coronal magnetic resonance imaging (MRI) scans of the index shoulder of Subject A9 treated with injection of UA-ADRCs, generated during the present and the former studies. Panels A-L show the same (or nearly the same) image planes at different times, with Panels A showing the most ventral image plane and Panels L the most dorsal image plane. The arrows in Panels B_W24_, C_W24_ and D_W24_ indicate a hyperintense structure at the position of the supraspinatus tendon that was found at 24 weeks post-treatment but not at baseline. Abbreviations: BL, baseline; W24 / W52, 24 / 52 weeks post-treatment; M33, 33 months post-treatment (no MRI was performed during the second visit of Subject A9).

**FIGURE S10.**
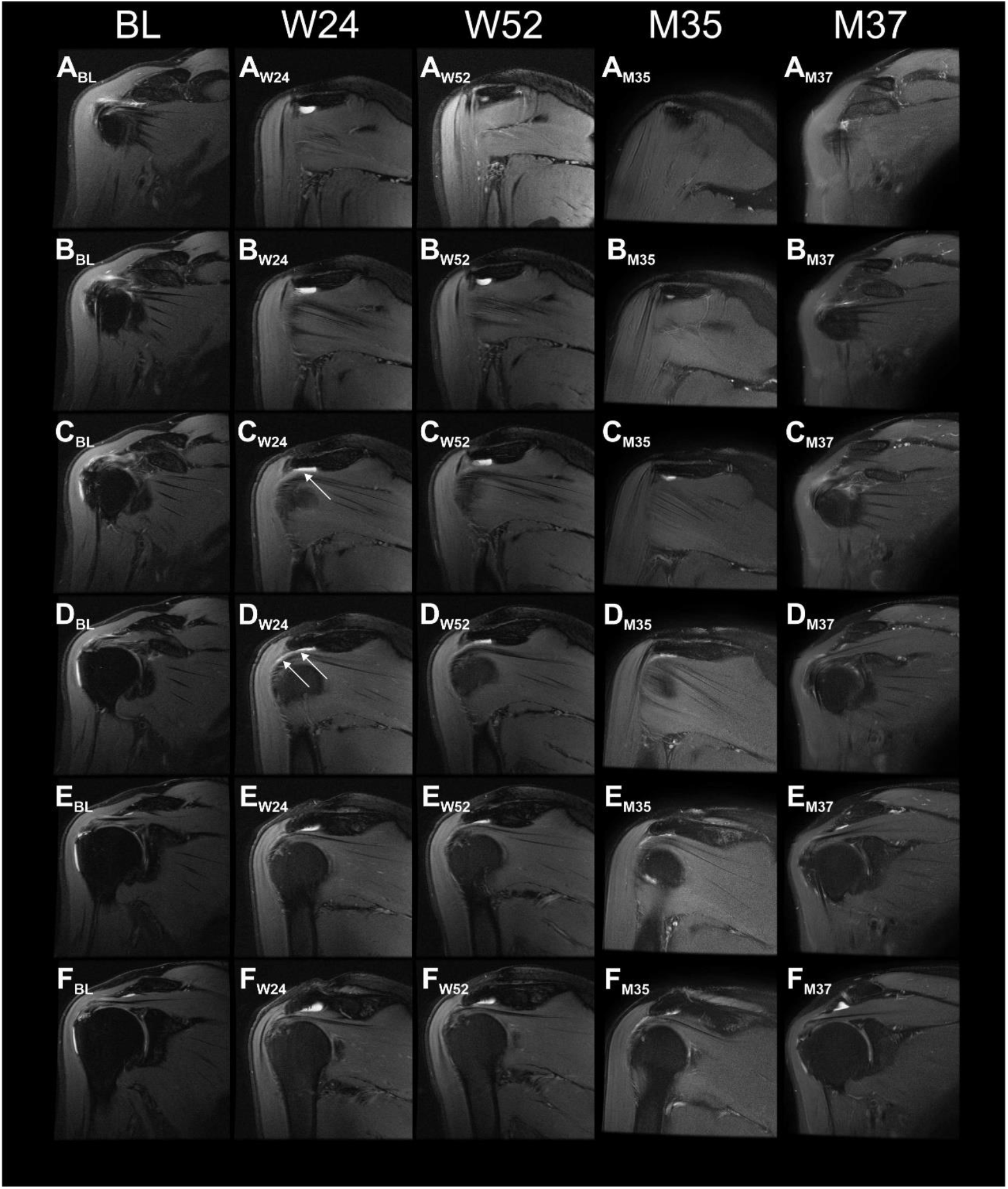
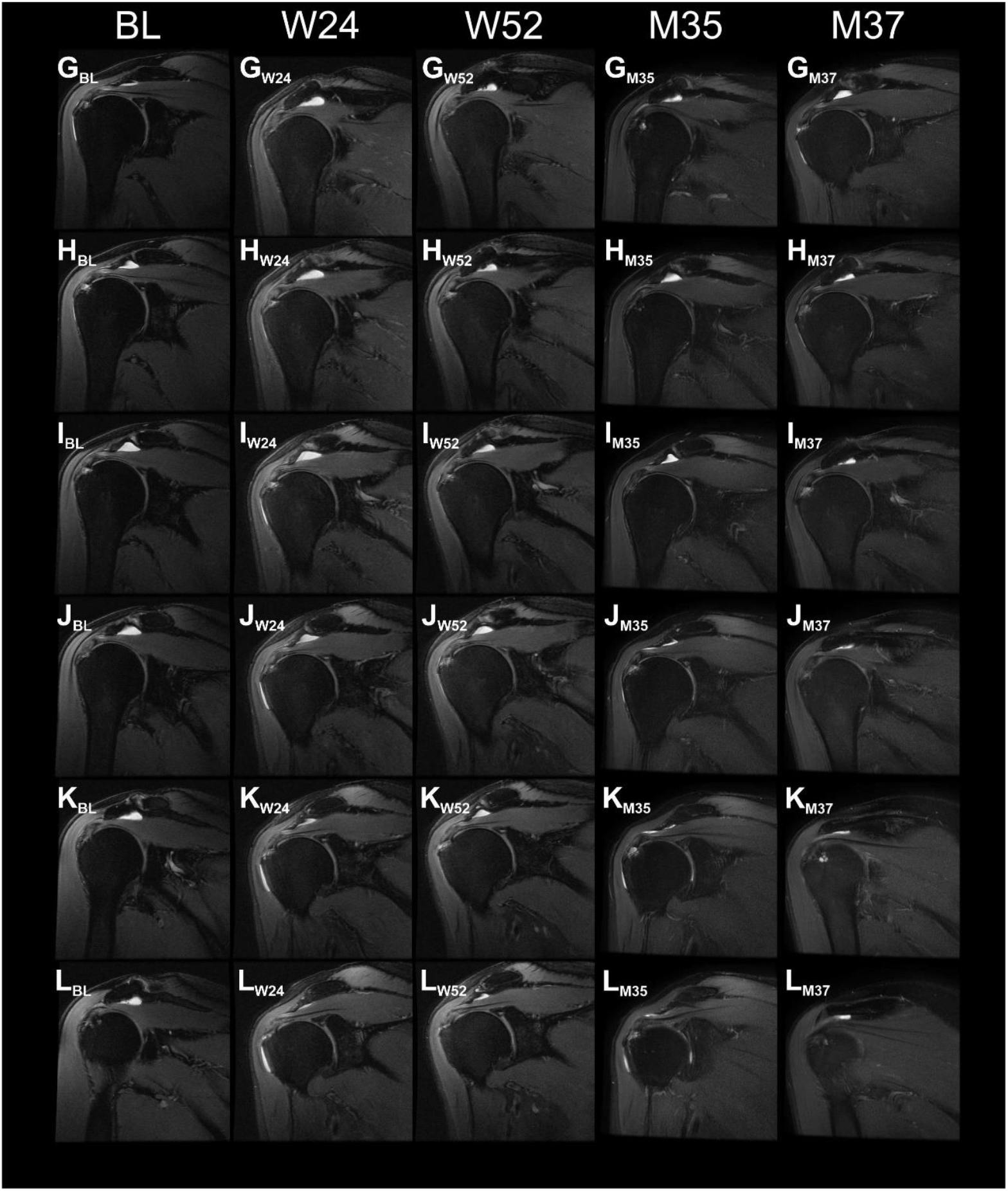
Proton density weighted, fat saturated, T2-weighted, coronal magnetic resonance imaging (MRI) scans of the index shoulder of Subject A10 treated with injection of UA-ADRCs, generated during the present and the former studies. Panels A-L show the same (or nearly the same) image planes at different times, with Panels A showing the most ventral image plane and Panels L the most dorsal image plane. The arrows in Panels C_W24_ and D_W24_ indicate a hyperintense structure at the position of the supraspinatus tendon that was found at 24 weeks post-treatment but not at baseline. Abbreviations: BL, baseline; W24 / W52, 24 / 52 weeks post-treatment; M35 / M41, 35 / 41 months post-treatment.

**FIGURE S11.**
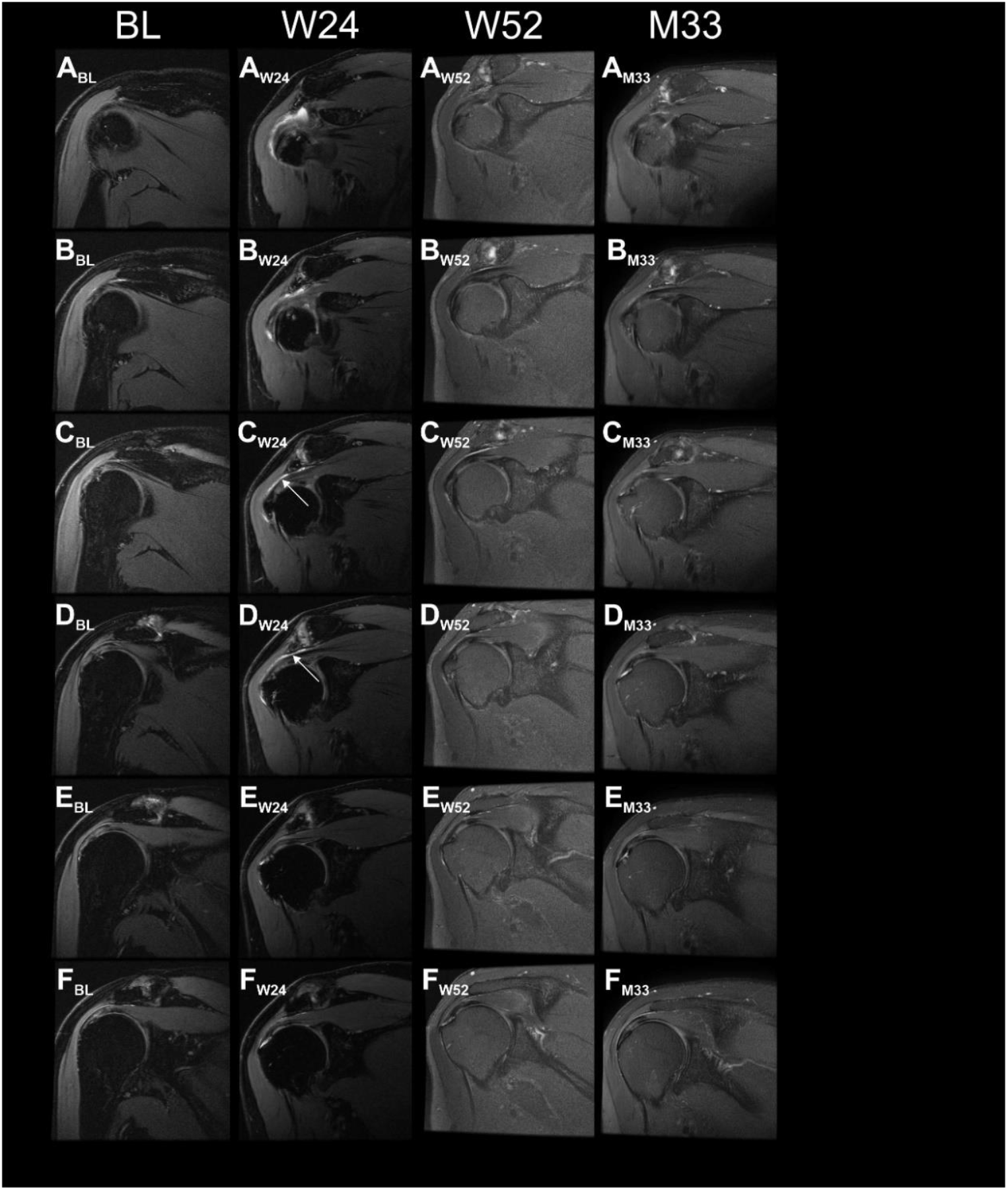
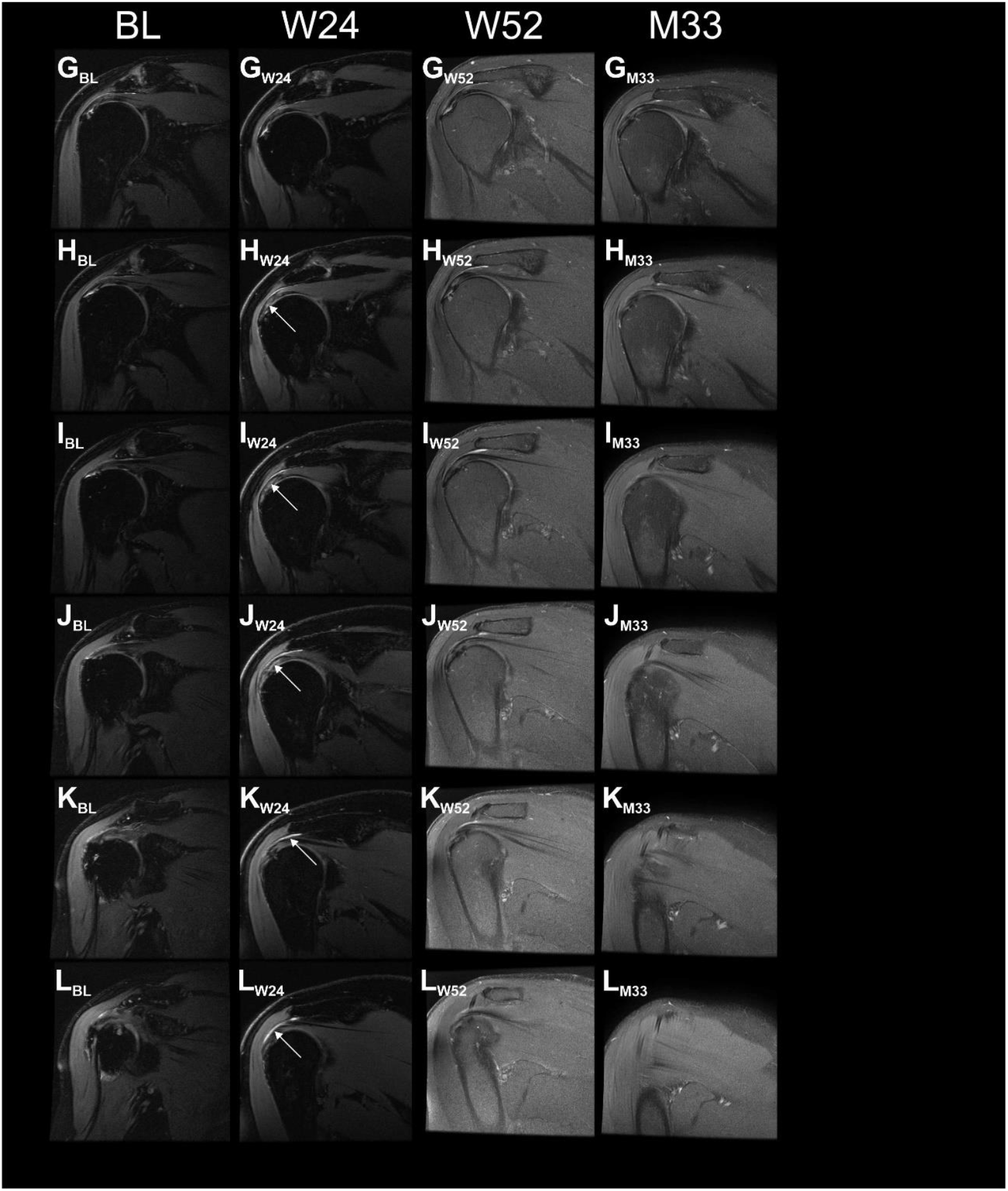
Proton density weighted, fat saturated, T2-weighted, coronal magnetic resonance imaging (MRI) scans of the index shoulder of Subject A11 treated with injection of UA-ADRCs, generated during the present and the former studies. Panels A-L show the same (or nearly the same) image planes at different times, with Panels A showing the most ventral image plane and Panels L the most dorsal image plane. The arrows in Panels C_W24_, D_W24_, H_W24_, I_W24_, J_W24_, K_W24_ and L_W24_ indicate a hyperintense structure at the position of the supraspinatus tendon that was found at 24 weeks post-treatment but not at baseline. Abbreviations: BL, baseline; W24 / W52, 24 / 52 weeks post-treatment; M33, 33 months post-treatment (note that no MRI was performed during the second visit of Subject A3).

**FIGURE S12.**
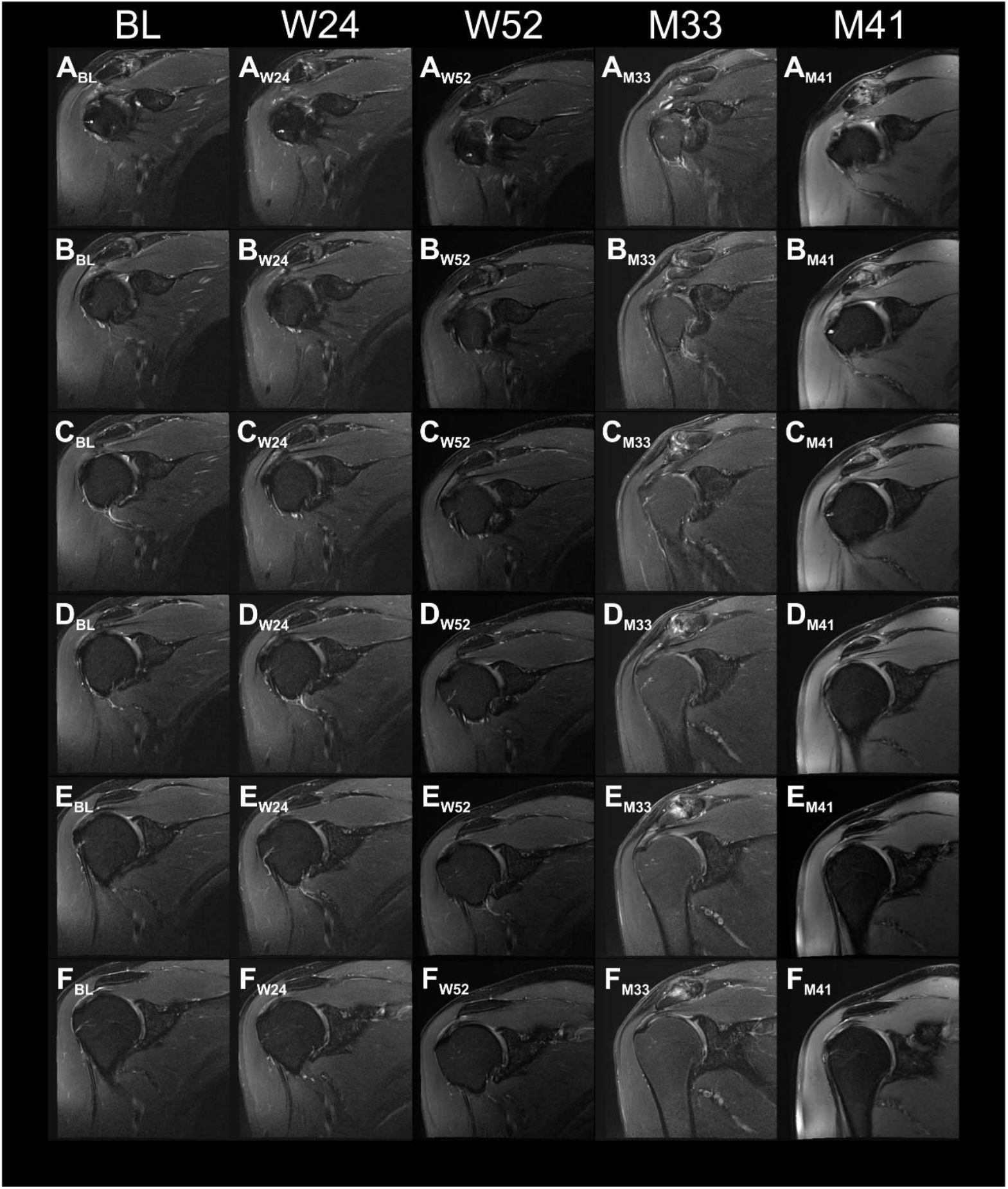
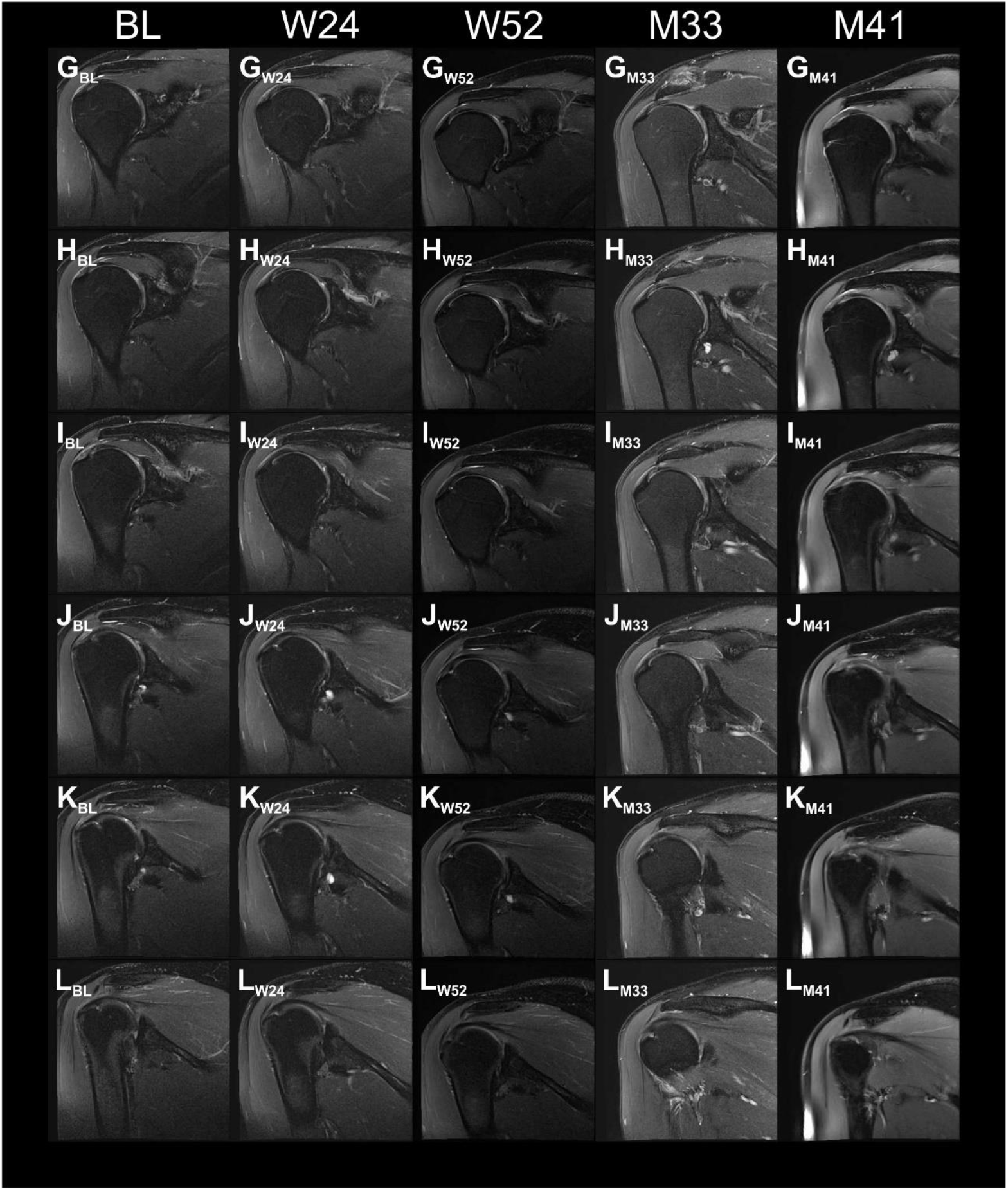
Proton density weighted, fat saturated, T2-weighted, coronal magnetic resonance imaging (MRI) scans of the index shoulder of Subject C1 treated with injection of corticosteroid, generated during the present and the former studies. Panels A-L show the same (or nearly the same) image planes at different times, with Panels A showing the most ventral image plane and Panels L the most dorsal image plane. Abbreviations: BL, baseline; W24 / W52, 24 / 52 weeks post-treatment; M33 / M41, 33 / 41 months post-treatment.

**FIGURE S13.**
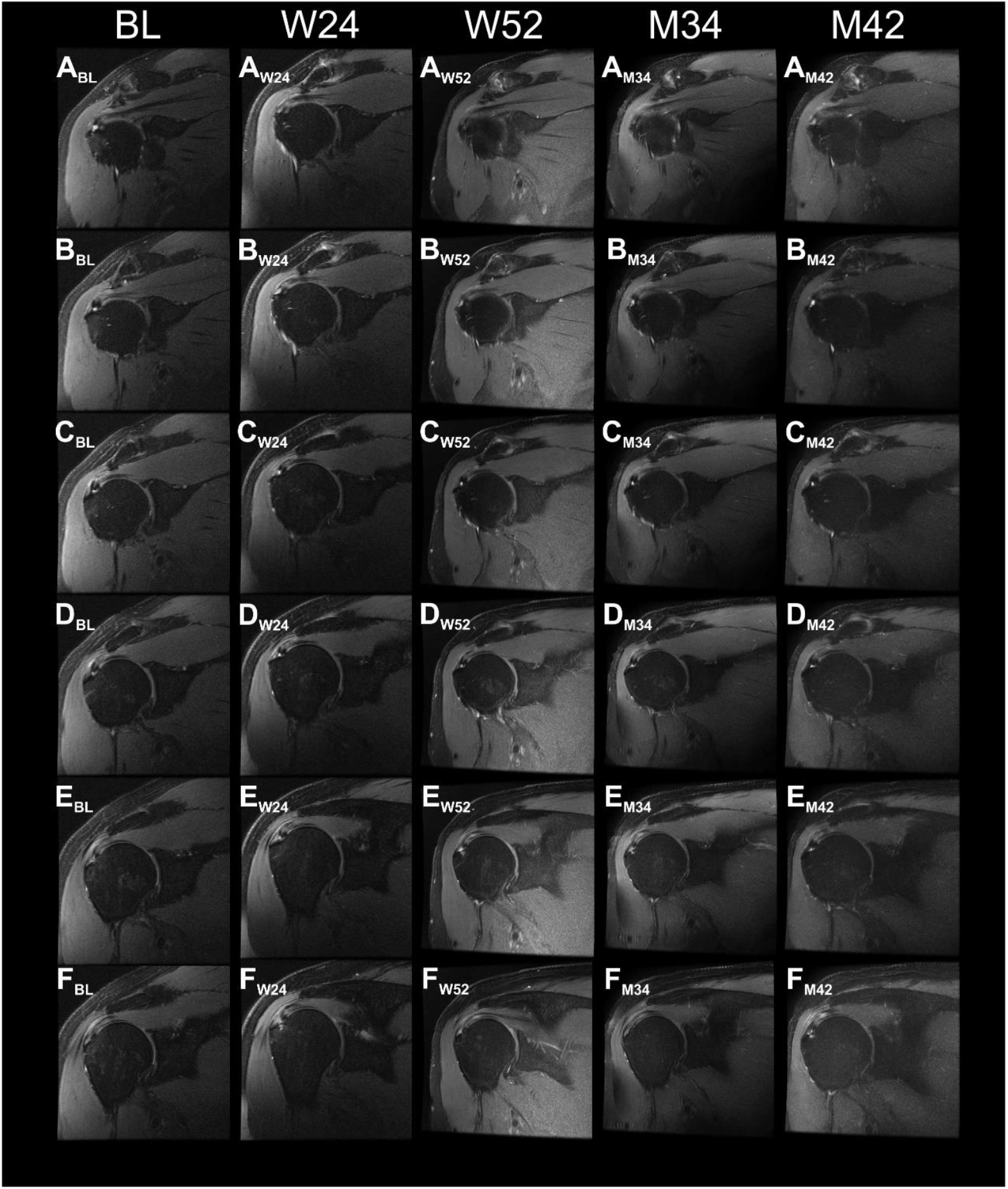
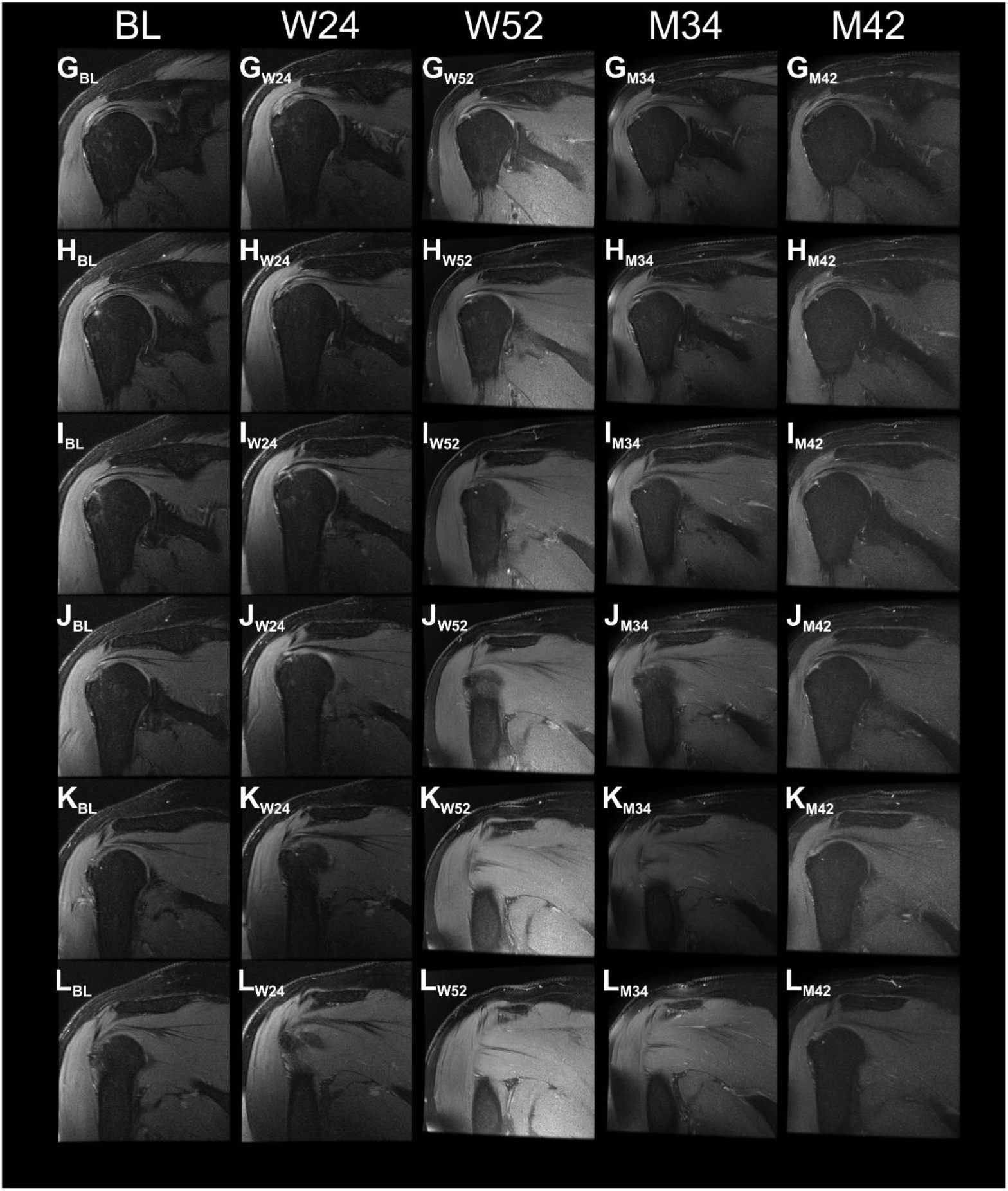
Proton density weighted, fat saturated, T2-weighted, coronal magnetic resonance imaging (MRI) scans of the index shoulder of Subject C2 treated with injection of corticosteroid, generated during the present and the former studies. Panels A-L show the same (or nearly the same) image planes at different times, with Panels A showing the most ventral image plane and Panels L the most dorsal image plane. Abbreviations: BL, baseline; W24 / W52, 24 / 52 weeks post-treatment; M34 / M42, 34 / 42 months post-treatment.

**FIGURE S14.**
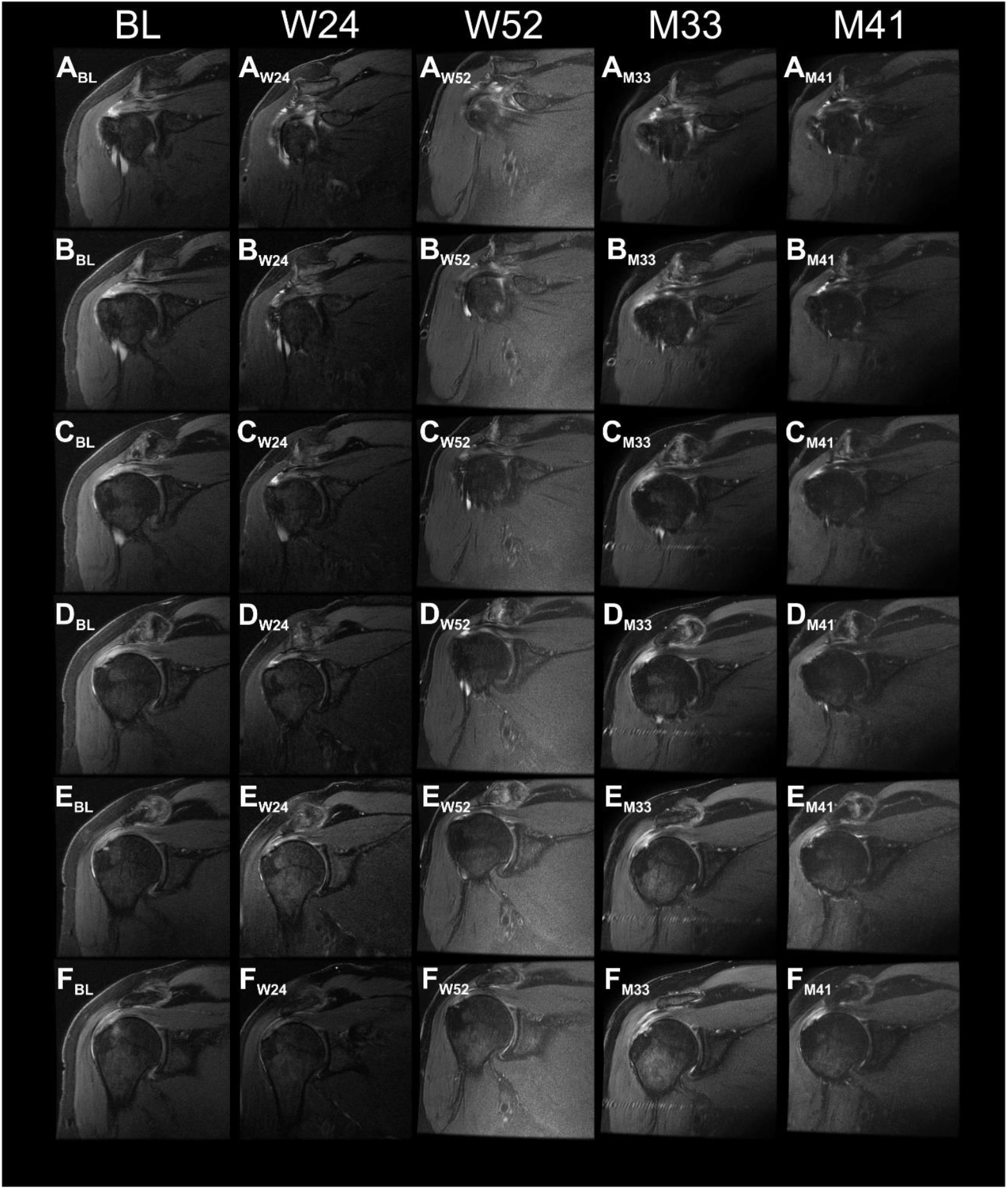
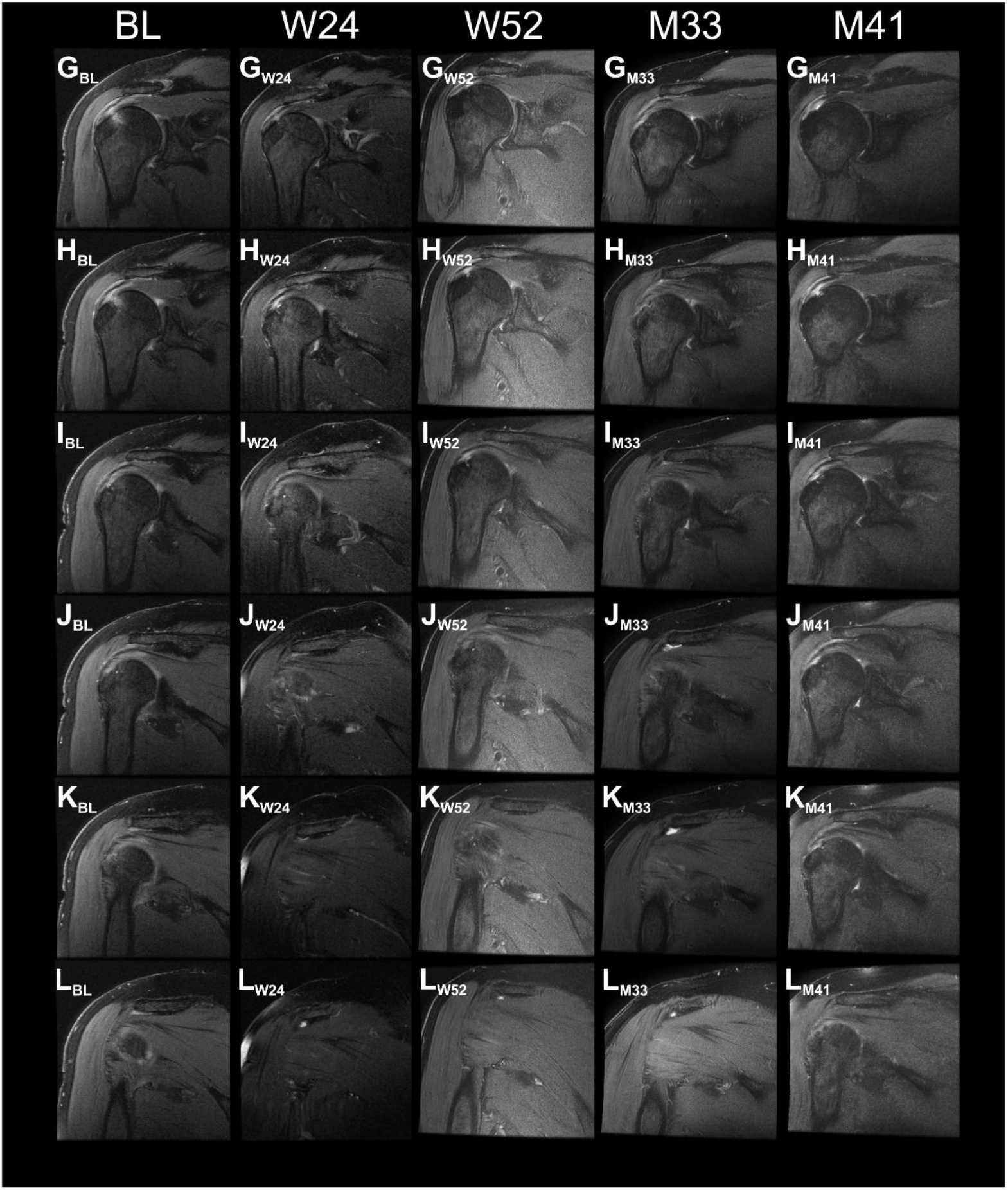
Proton density weighted, fat saturated, T2-weighted, coronal magnetic resonance imaging (MRI) scans of the index shoulder of Subject C3 treated with injection of corticosteroid, generated during the present and the former studies. Panels A-L show the same (or nearly the same) image planes at different times, with Panels A showing the most ventral image plane and Panels L the most dorsal image plane. Abbreviations: BL, baseline; W24 / W52, 24 / 52 weeks post-treatment; M33 / M41, 33 / 41 months post-treatment.

**FIGURE S15.**
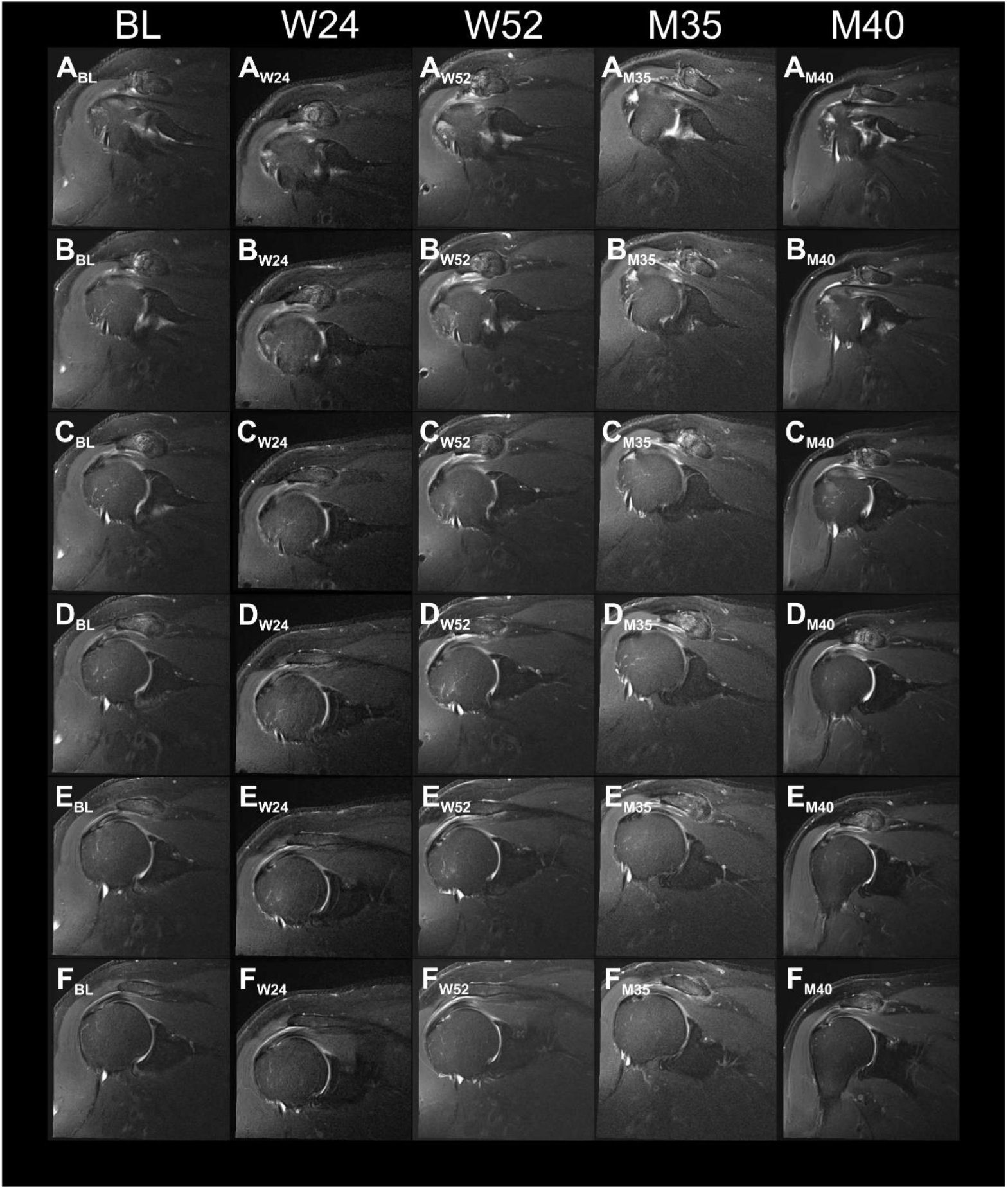
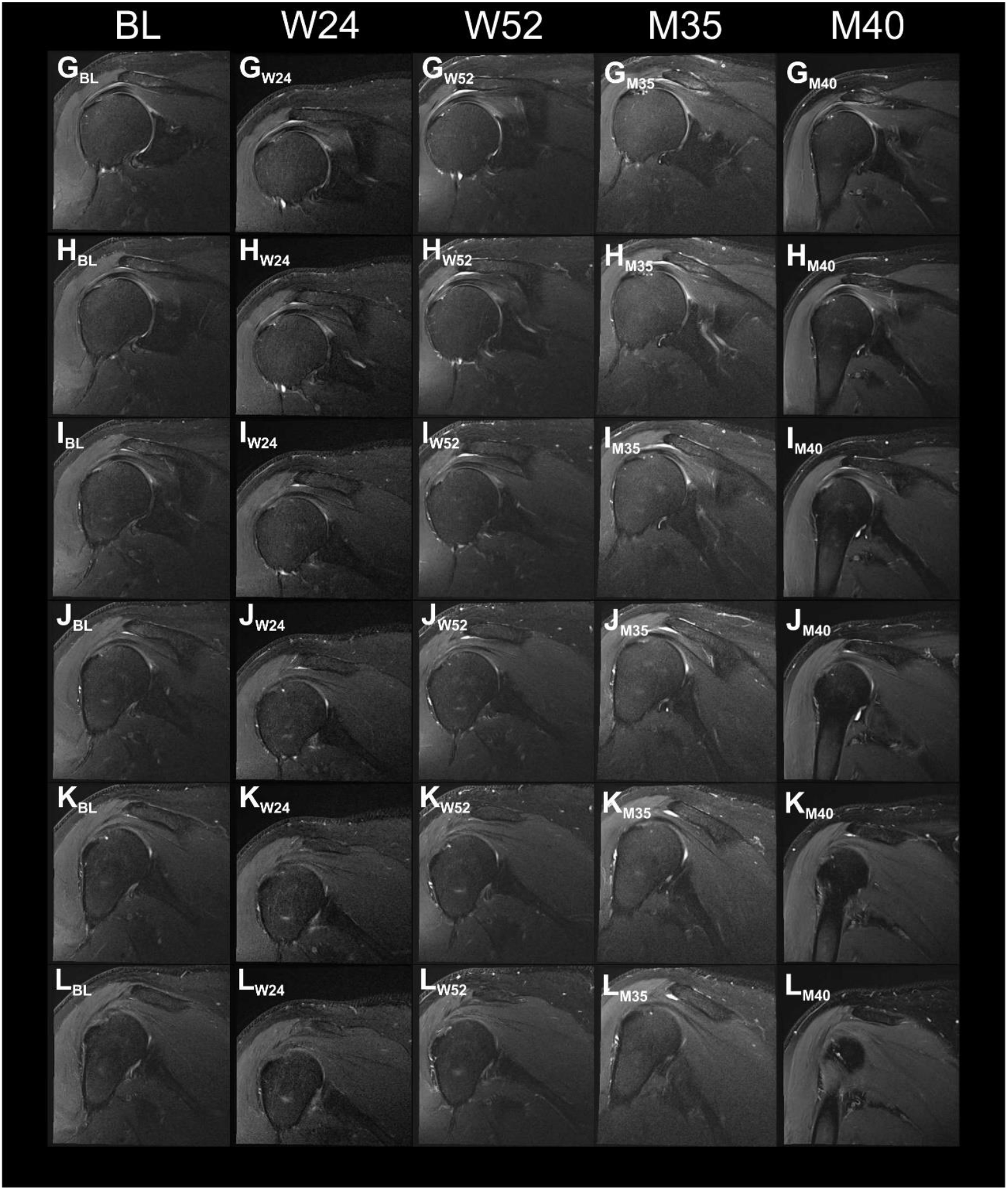
Proton density weighted, fat saturated, T2-weighted, coronal magnetic resonance imaging (MRI) scans of the index shoulder of Subject C4 treated with injection of corticosteroid, generated during the present and the former studies. Panels A-L show the same (or nearly the same) image planes at different times, with Panels A showing the most ventral image plane and Panels L the most dorsal image plane. Abbreviations: BL, baseline; W24 / W52, 24 / 52 weeks post-treatment; M35 / M40, 35 / 40 months post-treatment.

### Part 2 Treatment-emergent adverse events that occurred during the present and the former studies

#### 2.1. Details of all treatment-emergent adverse events that occurred during the present and the former studies

Tables S1 and S2 provide details of all treatment emergent adverse events (TEAEs) reported in the present and the former studies, stratified by the relation to the investigated treatment, severity and month post-treatment during which the TEAEs occurred.

**TABLE S1.** Treatment-emergent adverse events reported by those subjects who were treated with injection of UA-ADRCs. Adverse events related to the index shoulder are given boldface.

| Subject | M | Study | R | Severity | Descriptions/Comments |
| --- | --- | --- | --- | --- | --- |
| A11 | 0.1 | F | Possible | Moderate | **Index arm pain<sup>a</sup>** |
| A3 | 0.0 | F | Possible | Mild | Left mid-abdomen pain<sup>a</sup> |
| A5 | 0.0 | F | Possible | Mild | Pain mid-abdomen<sup>a</sup> |
| A2 | 0.5 | F | Possible | Mild | Cold<sup>a</sup> |
| A2 | 0.9 | F | Unlikely | Moderate | Shortness of Breath<sup>a</sup> |
| A5 | 1.2 | F | Unlikely | Moderate | Pulp necrosis with asymptomatic apical periodontitis Tooth #15<sup>a</sup> |
| A4 | 0.7 | F | Unlikely | Mild | **Worsening index shoulder pain<sup>a</sup>** |
| A1 | 1.4 | F | Unlikely | Mild | **Worsening index shoulder pain<sup>a</sup>** |
| A11 | 18.7 | P | Unlikely | Mild | **Index shoulder pain<sup>a</sup>** |
| A1 | 25.5 | P | Unlikely | Mild | **Index shoulder pain<sup>a</sup>** |
| A4 | 3.0 | F | Not Related | Severe | Non ST elevation myocardial infarction<sup>b</sup> |
| A4 | 4.1 | F | Not Related | Severe | ST elevation myocardial infarction<sup>b</sup> |
| A9 | 7.4 | F | Not Related | Severe | **Index shoulder pain<sup>b</sup>** |
| A7 | 34.4 | P | Not Related | Severe | Ganglion cyst of the non-index shoulder<sup>b</sup> |
| A7 | 1.3 | F | Not Related | Moderate | Concussion |
| A6 | 1.9 | F | Not Related | Moderate | Worsening tendonitis left wrist |
| A9 | 7.6 | F | Not Related | Moderate | Bronchitis |
| A7 | 8.0 | F | Not Related | Moderate | Esophageal dysphagia |
| A7 | 9.7 | F | Not Related | Moderate | Chronic obstructive lung disease |
| A1 | 9.8 | F | Not Related | Moderate | Diverticulitis |
| A7 | 12.0 | F | Not Related | Moderate | Shortness of breath |
| A3 | 33.5 | P | Not Related | Moderate | **Index shoulder pain<sup>c</sup>** |
| A4 | 34.2 | P | Not Related | Moderate | Interstitial pneumonia |
| A8 | 34.9 | P | Not Related | Moderate | Alcoholic relapse |
| A4 | 36.8 | P | Not Related | Moderate | Right leg pain |
| A8 | 38.7 | P | Not Related | Moderate | Dyspnea |
| A7 | 40.4 | P | Not Related | Moderate | Muscle spasms |
| A9 | -0.3 | F | Not Related | Mild | Asthmatic bronchitis |
| A9 | 0.3 | F | Not Related | Mild | Left knee pain |
| A8 | 0.6 | F | Not Related | Mild | Dysuria |
| A9 | 1.0 | F | Not Related | Mild | Contusion of right elbow |
| A3 | 1.1 | F | Not Related | Mild | Worsening lower back pain |
| A7 | 1.6 | F | Not Related | Mild | Sinusitis |
| A2 | 2.1 | F | Not Related | Mild | Worsening allergic rhinitis |
| A7 | 2.8 | F | Not Related | Mild | Abdominal discomfort |

**TABLE S1 (cont.)**
| Subject | M | Study | R | Severity | Descriptions/Comments |
| --- | --- | --- | --- | --- | --- |
| A11 | 3.9 | F | Not Related | Mild | Left hand pain |
| A10 | 4.1 | F | Not Related | Mild | Toenail thickening and discoloration |
| A10 | 4.1 | F | Not Related | Mild | Seborrheic keratosis |
| A2 | 4.2 | F | Not Related | Mild | Essential hypertension |
| A1 | 4.4 | F | Not Related | Mild | Pharyngitis |
| A8 | 4.6 | F | Not Related | Mild | Relapse of alcohol dependence |
| A11 | 4.6 | F | Not Related | Mild | Staph infection of fourth ray |
| A7 | 5.5 | F | Not Related | Mild | Acute pain left shoulder |
| A8 | 6.8 | F | Not Related | Mild | Left foot pain |
| A11 | 31.4 | P | Not Related | Mild | influenza |
| A9 | 32.7 | P | Not Related | Mild | **Device material fragmentation in index shoulder<sup>b</sup>** |
| A10 | 34.1 | P | Not Related | Mild | Epigastric pain |
| A4 | 34.3 | P | Not Related | Mild | Sleep apnea |
| A9 | 35.7 | P | Not Related | Mild | Bronchitis |
| A11 | 36.7 | P | Not Related | Mild | Sleep disturbance |
| A11 | 36.7 | P | Not Related | Mild | Tinnitus |
| A11 | 36.7 | P | Not Related | Mild | Hearing loss |
| A8 | 37.2 | P | Not Related | Mild | Acute cystitis |
| A9 | 39.5 | P | Not Related | Mild | Bursal hematoma left knee |
| A4 | 40.4 | P | Not Related | Mild | Benign prostatic hyperplasia |
| A5 | 40.6 | P | Not Related | Mild | Cataract |
| A5 | 40.9 | P | Not Related | Mild | Back pain |
| A7 | 41.4 | P | Not Related | Mild | Baclofen allergy |

Abbreviations: UA-ADRCs, fresh, uncultured, unmodified, autologous, adipose-derived regenerative cells; M, month post-treatment when the treatment-emergent adverse event started; F, former study; P, present study; R, relation to the investigated treatment.
<sup>a</sup>, outlined in detail in Table S8.
<sup>b</sup>, outlined in detail in Table S13.
<sup>c</sup>, this subject reported at 33.3 months post-treatment that he was doing very well until about 30.3 months post-treatment. At this time he moved into a new house and was doing a lot of work that involved repetitive use of his right (index) arm. He started to have increased pain in his right shoulder. Because of this he had a recent subacromial corticosteroid injection (i.e., before the first study visit). At the time of the first study visit he had no pain in his right shoulder.

**TABLE S2.** Treatment-emergent adverse events reported by those subjects who were treated with injection of corticosteroid. Adverse events related to the index shoulder are given boldface.

| Subject | M | Study | R | Severity | Descriptions/Comments |
| --- | --- | --- | --- | --- | --- |
| C4 | 1.4 | F | Possible | Moderate | **Anterior pain in index shoulder<sup>a</sup>** |
| C2 | 1.6 | F | Possible | Moderate | **Worsening index shoulder pain<sup>a</sup>** |
| C2 | 0.1 | F | Unlikely | Mild | Pharyngitis<sup>a</sup> |
| C5<sup>b</sup> | 2.1 | F | Unlikely | Mild | **Worsening index shoulder pain<sup>a</sup>** |
| C3 | 5.2 | F | Unlikely | Mild | **Worsening index shoulder pain<sup>a</sup>** |
| C3 | 1.0 | F | Not Related | Moderate | Biceps tear at elbow |
| C2 | 5.4 | F | Not Related | Moderate | Right upper broken tooth |
| C1 | 36.1 | P | Not Related | Moderate | Multilevel lumbar degenerative disc disease |
| C2 | 38.2 | P | Not Related | Moderate | Actinic keratoses |
| C2 | 38.2 | P | Not Related | Moderate | Squamous cell carcinoma of skin, left neck |
| C2 | 38.2 | P | Not Related | Moderate | Basal cell carcinoma, left inferior neck |
| C5<sup>b</sup> | 0.7 | F | Not Related | Mild | Coronary artery disease |
| C3 | 1.1 | F | Not Related | Mild | Left finger sprain |
| C2 | 3.7 | F | Not Related | Mild | Hip Pain |
| C1 | 4.8 | F | Not Related | Mild | Chest pain |
| C2 | 4.8 | F | Not Related | Mild | Cough |
| C2 | 32.3 | P | Not Related | Mild | **Index shoulder pain<sup>c</sup>** |
| C4 | 32.3 | P | Not Related | Mild | Upper respiratory infection |
| C3 | 34.9 | P | Not Related | Mild | Benign skin lesion |
| C1 | 35.3 | P | Not Related | Mild | Otitis media |
| C4 | 36.5 | P | Not Related | Mild | Hypogammaglobulinemia<sup>a</sup> |
| C1 | 37.2 | P | Not Related | Mild | **Index shoulder pain<sup>d</sup>** |
| C2 | 37.8 | P | Not Related | Mild | Nausea |
| C2 | 40.5 | P | Not Related | Mild | Lymphadenopathy |
| C2 | 40.5 | P | Not Related | Mild | Atrial fibrillation |

Abbreviations: M, month post-treatment when the treatment-emergent adverse event started; F, former study; P, present study; R, relation to the investigated treatment.
<sup>a</sup>, outlined in detail in Table S9.
<sup>b</sup>, this subject was enrolled in the former study but not in the present study.
<sup>c</sup>, this subject fell off a ladder at 32.3 months post-treatment. He reported increased pain and discomfort and difficulty with overhead activity. An MRI performed during the second study visit at 33.3 months post-treatment revealed a full thickness tear of anterior superior fibers of the supraspinatus tendon. An MRI performed during the second study visit at 41.2 months post-treatment showed a near full thickness tear of the anterosuperior fibers of the supraspinatus tendon.
<sup>d</sup>, this subject reported pain in his left shoulder at 37.2 months post-treatment that kept him awake at night; he was unable to sleep on his left side.

#### 2.2. Statistical analysis of all treatment-emergent adverse events that occurred during the present and the former studies

The total number of TEAEs was 83, of which 58 (69.9%) occurred in the UA-ADRCs group and 25 (30.1%) in the corticosteroid group (Tables S1 and S2). Accordingly, the average number of TEAEs per subject was 5.3 ± 2.7 (mean ± standard error of the mean) in the UA-ADRCs group and 5.0 ± 1.8 in the corticosteroid group.

The distribution of these 83 TEAEs with regard to severity and relation to the investigated treatment was as follows: there were…

- four severe TEAEs (4.8%), none of which were related to the investigated treatment (all in the UA-ADRCs group),
- three moderate TEAEs probably related to the investigated treatment (3.6%) (one in the UA-ADRCs group and two in the corticosteroid group),
- three mild TEAEs probably related to the investigated treatment (3.6%) (all in the UA-ADRCs group),
- two moderate TEAEs unlikely to be related to the investigated treatment (2.4%) (all in the UA-ADRCs group),
- seven mild TEAEs unlikely to be related to the investigated treatment (8.4%) (four in the UA-ADRCs group and three in the corticosteroid group),
- 19 moderate TEAEs not related to the investigated treatment (22.9%), and
- 45 mild TEAEs not related to the investigated treatment (54.2%).

Table S3 shows group-specific numbers of subjects who experienced a certain number of TEAEs (between 0 and 12) in the present and the former studies.

**TABLE S3.**
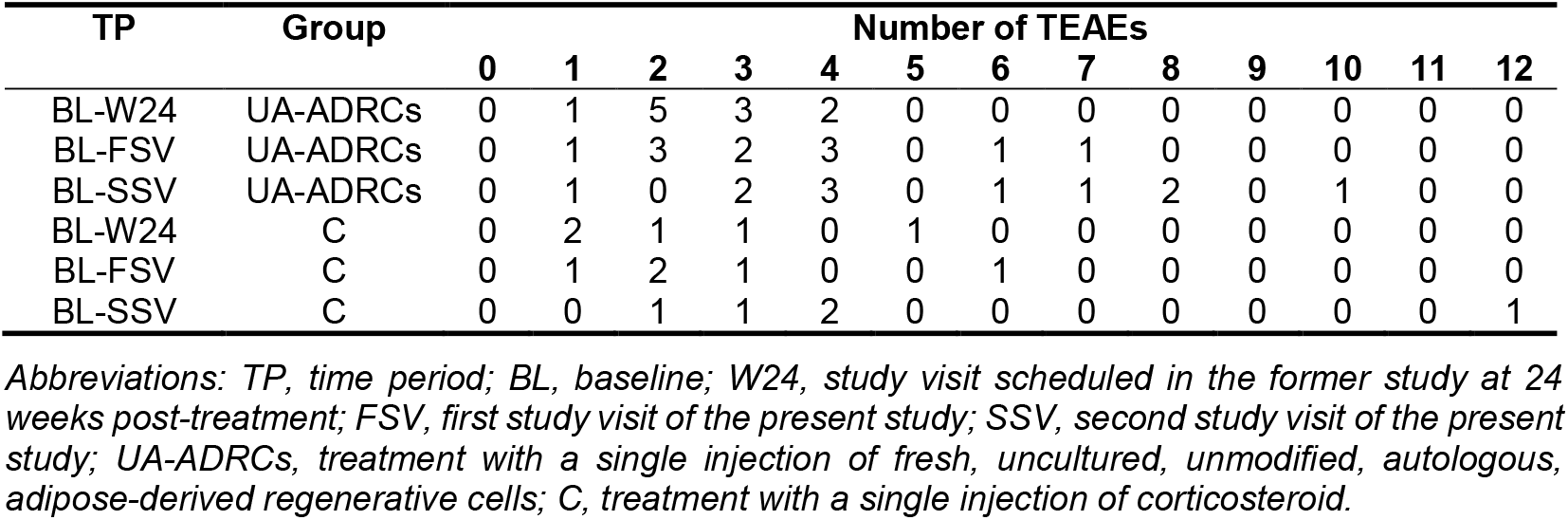
Group-specific numbers of subjects who experienced a certain number of treatment-emergent adverse events (between 0 and 12) in the present and the former studies.

| TP | Group | Number of TEAEs |  |  |  |  |  |  |  |  |  |  |  |  |
| --- | --- | --- | --- | --- | --- | --- | --- | --- | --- | --- | --- | --- | --- | --- |
|  |  | 0 | 1 | 2 | 3 | 4 | 5 | 6 | 7 | 8 | 9 | 10 | 11 | 12 |
| BL-W24 | UA-ADRCs | 0 | 1 | 5 | 3 | 2 | 0 | 0 | 0 | 0 | 0 | 0 | 0 | 0 |
| BL-FSV | UA-ADRCs | 0 | 1 | 3 | 2 | 3 | 0 | 1 | 1 | 0 | 0 | 0 | 0 | 0 |
| BL-SSV | UA-ADRCs | 0 | 1 | 0 | 2 | 3 | 0 | 1 | 1 | 2 | 0 | 1 | 0 | 0 |
| BL-W24 | C | 0 | 2 | 1 | 1 | 0 | 1 | 0 | 0 | 0 | 0 | 0 | 0 | 0 |
| BL-FSV | C | 0 | 1 | 2 | 1 | 0 | 0 | 1 | 0 | 0 | 0 | 0 | 0 | 0 |
| BL-SSV | C | 0 | 0 | 1 | 1 | 2 | 0 | 0 | 0 | 0 | 0 | 0 | 0 | 1 |

*Abbreviations: TP, time period; BL, baseline; W24, study visit scheduled in the former study at 24 weeks post-treatment; FSV, first study visit of the present study; SSV, second study visit of the present study; UA-ADRCs, treatment with a single injection of fresh, uncultured, unmodified, autologous, adipose-derived regenerative cells; C, treatment with a single injection of corticosteroid.*

For all investigated time periods there was no statistically significant difference between the groups with regard to the numbers of subjects who experienced a certain number of TEAEs (between 0 and 12) in the present and the former studies (Chi-square test for trend):

- from baseline to W24 in the former study: p = 0.809,
- from baseline to the first study visit of the present study: p = 0.488, and
- from baseline to the second study visit of the present study: p = 0.778.

Table S4 shows group-specific numbers of TEAEs that were classified as {mild / moderate / severe} in the present and the former studies. Table S5 summarizes absolute numbers of all TEAEs reported in the present and the former studies, stratified by the relation to the investigated treatment, severity and time period during which the TEAEs occurred. Table S6 summarizes the corresponding mean numbers of TEAEs per subject in each group.

**TABLE S4.** Group-specific numbers of TEAEs that were classified as {mild / moderate / severe} in the present and the former studies.

| TP | Group | Classification of TEAEs |  |  |
| --- | --- | --- | --- | --- |
|  |  | Mild | Moderate | Severe |
| BL-W24 | UA-ADRCs | 21 | 5 | 2 |
| BL-FSV | UA-ADRCs | 25 | 10 | 3 |
| BL-SSV | UA-ADRCs | 38 | 16 | 4 |
| BL-W24 | C | 8 | 4 | 0 |
| BL-FSV | C | 10 | 4 | 0 |
| BL-SSV | C | 17 | 8 | 0 |

*Abbreviations: TP, time period; BL, baseline; W24, study visit scheduled in the former study at 24 weeks post-treatment; FSV, first study visit of the present study; SSV, second study visit of the present study; UA-ADRCs, treatment with a single injection of fresh, uncultured, unmodified, autologous, adipose-derived regenerative cells; C, treatment with a single injection of corticosteroid.*

For all investigated time periods there was no statistically significant difference between the groups with regard to the numbers of TEAEs that were classified as {mild / moderate / severe} in the present and the former studies (Chi-square test for trend):

- from baseline to W24 in the former study: p = 0.951,
- from baseline to the first study visit of the present study: p = 0.468, and
- from baseline to the second study visit of the present study: p = 0.497.

**TABLE S5.** Absolute numbers of TEAEs reported in the present and the former studies, stratified by the relation to the investigated treatment, severity and time period during which the TEAEs occurred.

| TP | Group | P-se | P-mo | P-mi | U-se | U-mo | U-mi | NR-se | NR-mo | NR-mi |
| --- | --- | --- | --- | --- | --- | --- | --- | --- | --- | --- |
| BL-W24 | UA-ADRCs | 0 | 1 | 3 | 0 | 2 | 2 | 2 | 2 | 16 |
| BL-FSV | UA-ADRCs | 0 | 1 | 3 | 0 | 2 | 4 | 3 | 7 | 18 |
| BL-SSV | UA-ADRCs | 0 | 1 | 3 | 0 | 2 | 4 | 4 | 13 | 31 |
| BL-W24 | C | 0 | 2 | 0 | 0 | 0 | 3 | 0 | 2 | 5 |
| BL-FSV | C | 0 | 2 | 0 | 0 | 0 | 3 | 0 | 2 | 7 |
| BL-SSV | C | 0 | 2 | 0 | 0 | 0 | 3 | 0 | 6 | 14 |

*Abbreviations: TP, time period; BL, baseline; W24, study visit scheduled in the former study at 24 weeks post-treatment; FSV, first study visit of the present study; SSV, second study visit of the present study; UA-ADRCs, treatment with a single injection of fresh, uncultured, unmodified, autologous, adipose-derived regenerative cells; C, treatment with a single injection of corticosteroid; P, relation to the investigated treatment probable; U, relation to the investigated treatment unlikely; NR, no relation to the investigated treatment; se, severe, mo, moderate; mi, mild.*

**TABLE S6.** Group-specific mean numbers of treatment-emergent adverse events per subject reported in the present and the former studies, stratified by the relation to the investigated treatment, severity and time period during which the TEAEs occurred.

| TP | Group | P-se | P-mo | P-mi | U-se | U-mo | U-mi | NR-se | NR-mo | NR-mi |
| --- | --- | --- | --- | --- | --- | --- | --- | --- | --- | --- |
| BL-W24 | UA-ADRCs | 0 | 0.09 | 0.27 | 0 | 0.18 | 0.18 | 0.18 | 0.18 | 1.45 |
| BL-FSV | UA-ADRCs | 0 | 0.09 | 0.27 | 0 | 0.18 | 0.36 | 0.27 | 0.64 | 1.64 |
| BL-SSV | UA-ADRCs | 0 | 0.09 | 0.27 | 0 | 0.18 | 0.36 | 0.36 | 1.18 | 2.82 |
| BL-W24 | C | 0 | 0.40 | 0 | 0 | 0 | 0.60 | 0 | 0.40 | 1.00 |
| BL-FSV | C | 0 | 0.40 | 0 | 0 | 0 | 0.60 | 0 | 0.40 | 1.40 |
| BL-SSV | C | 0 | 0.40 | 0 | 0 | 0 | 0.60 | 0 | 1.20 | 2.80 |

Abbreviations: TP, time period; BL, baseline; W24, study visit scheduled in the former study at 24 weeks post-treatment; FSV, first study visit of the present study; SSV, second study visit of the present study; UA-ADRCs, treatment with a single injection of fresh, uncultured, unmodified, autologous, adipose-derived regenerative cells; C, treatment with a single injection of corticosteroid; P, relation to the investigated treatment probable; U, relation to the investigated treatment unlikely; NR, no relation to the investigated treatment; se, severe, mo, moderate; mi, mild.

Table S7 shows group-specific numbers of TEAEs that were classified as {not related / unlikely to be related / possibly related / probably related / definitely related} to the investigated treatment in the present and the former studies. Tables S8 and S9 summarize the individual courses of all TEAEs classified as {unlikely to be / possibly} related to the investigated treatment that occurred during the present and the former studies, experienced by those subjects who were treated with injection of UA-ADRCs (Table 8) and those subjects who were treated with injection of corticosteroid (Table S9).

**TABLE S7.**
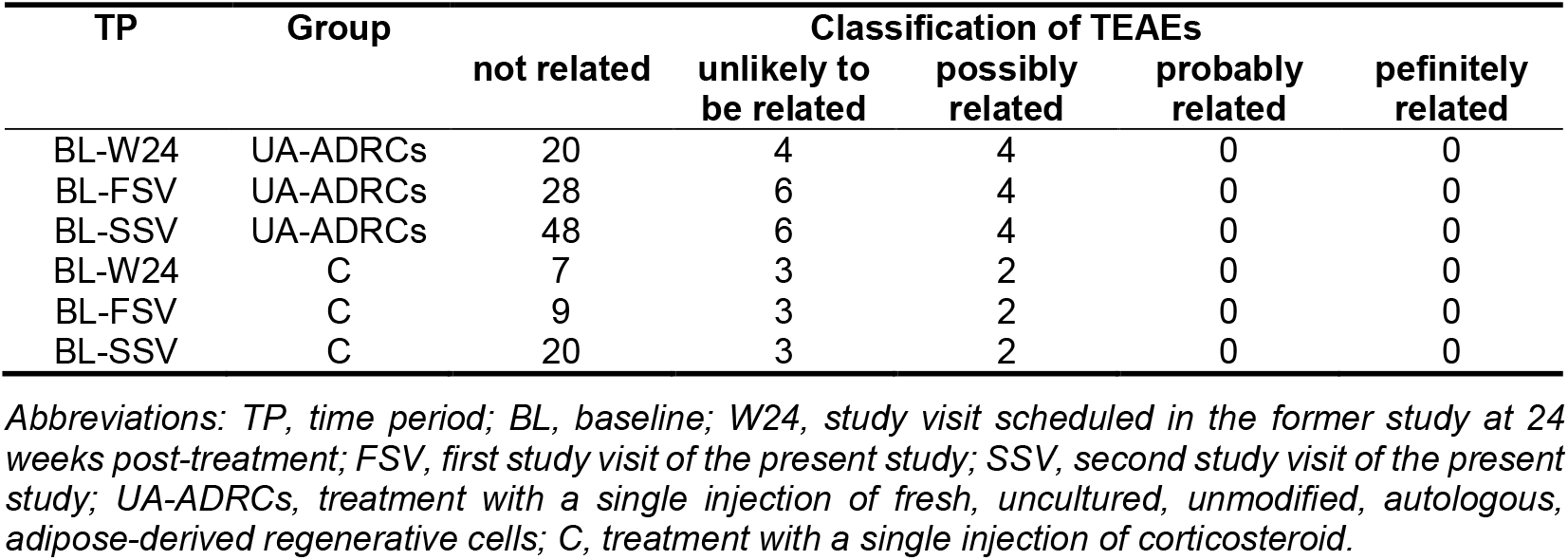
Group-specific numbers of TEAEs that were classified as {not related / unlikely to be related / possibly related / probably related / definitely related} to the investigated treatment in the present and the former studies.

For all investigated time periods there was no statistically significant difference between the groups with regard to the numbers of TEAEs that were classified as {not related / unlikely to be related / possibly related / probably related / definitely related} to the investigated treatment in the present and the former studies (Chi-square test):

- from baseline to W24 in the former study: p = 0.672,
- from baseline to the first study visit of the present study: p = 0.802, and
- from baseline to the second study visit of the present study: p = 0.956.

**TABLE S8.**
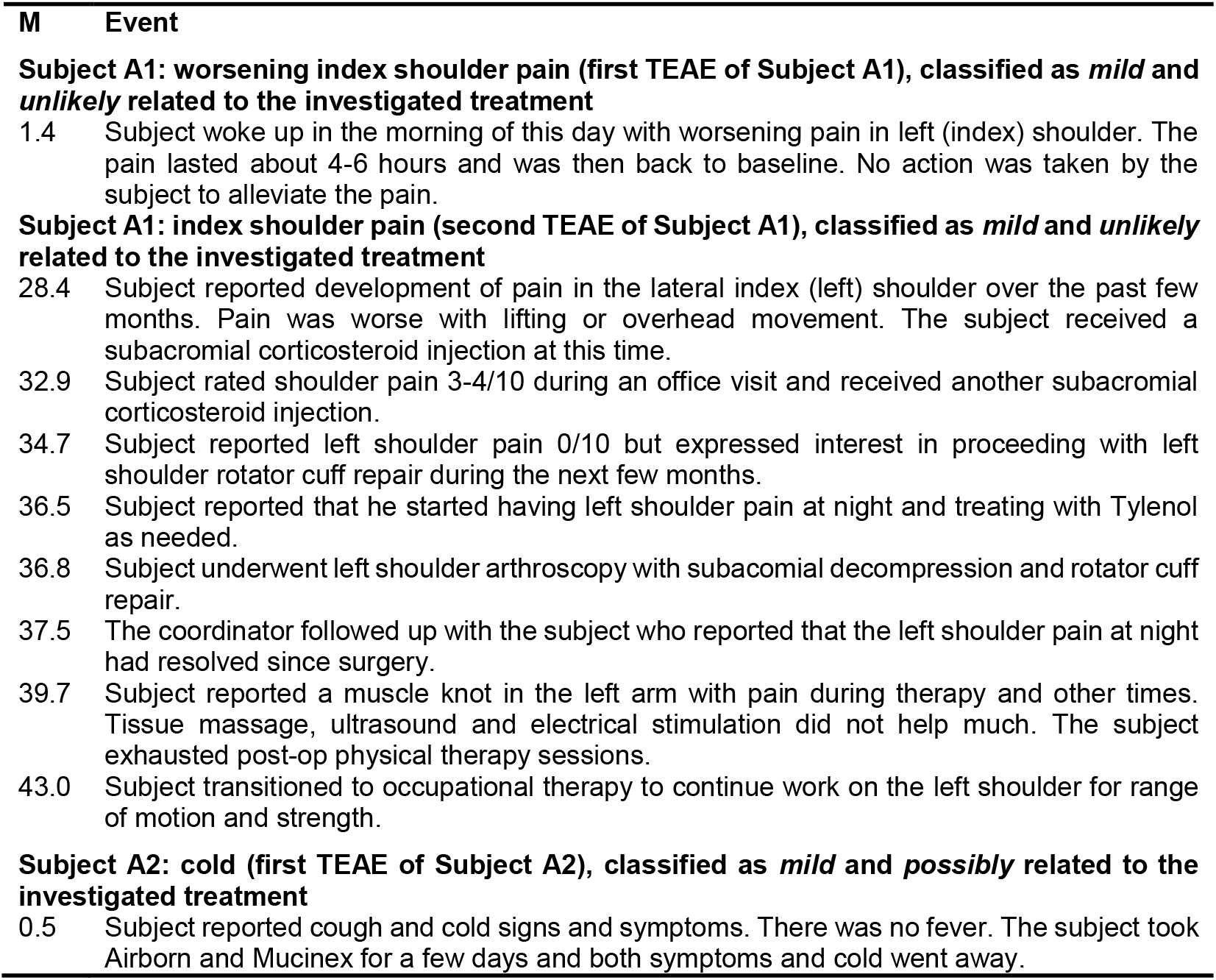
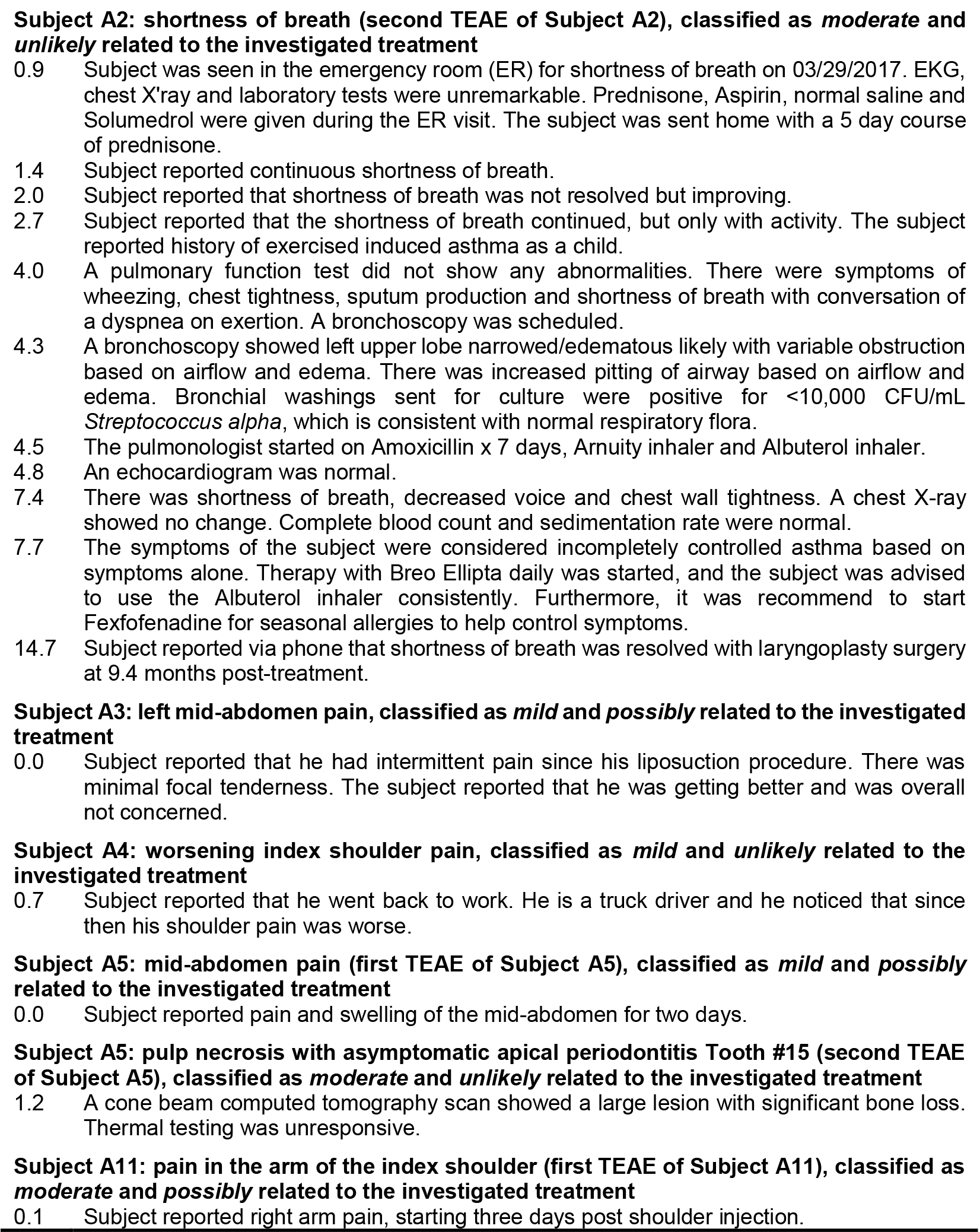
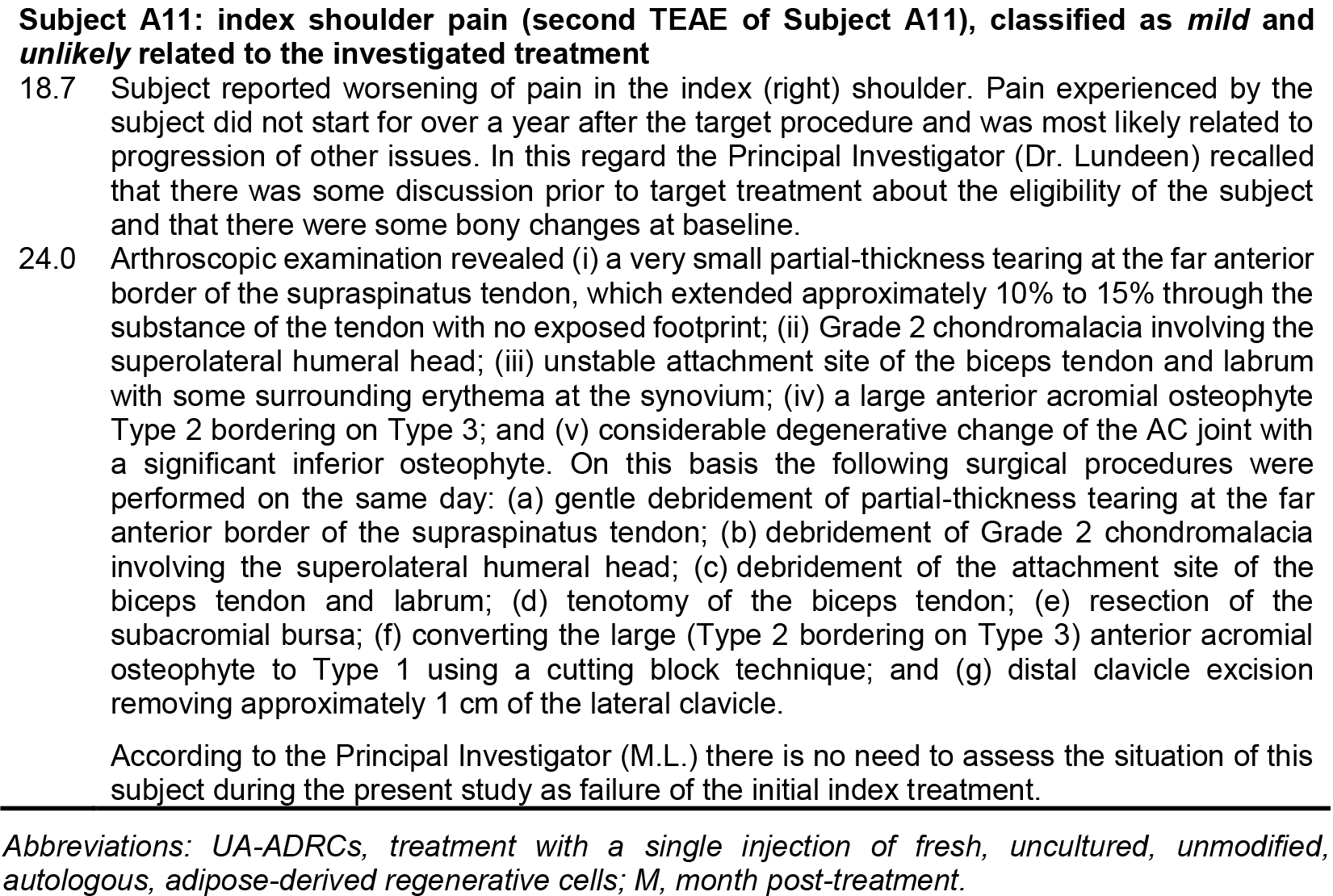
Individual courses of the treatment-related adverse events classified as {unlikely to be / possibly related} to the investigated treatment that occurred during the present and the former studies, experienced by those subjects who were treated with injection of UA-ADRCs.

**TABLE S9.**
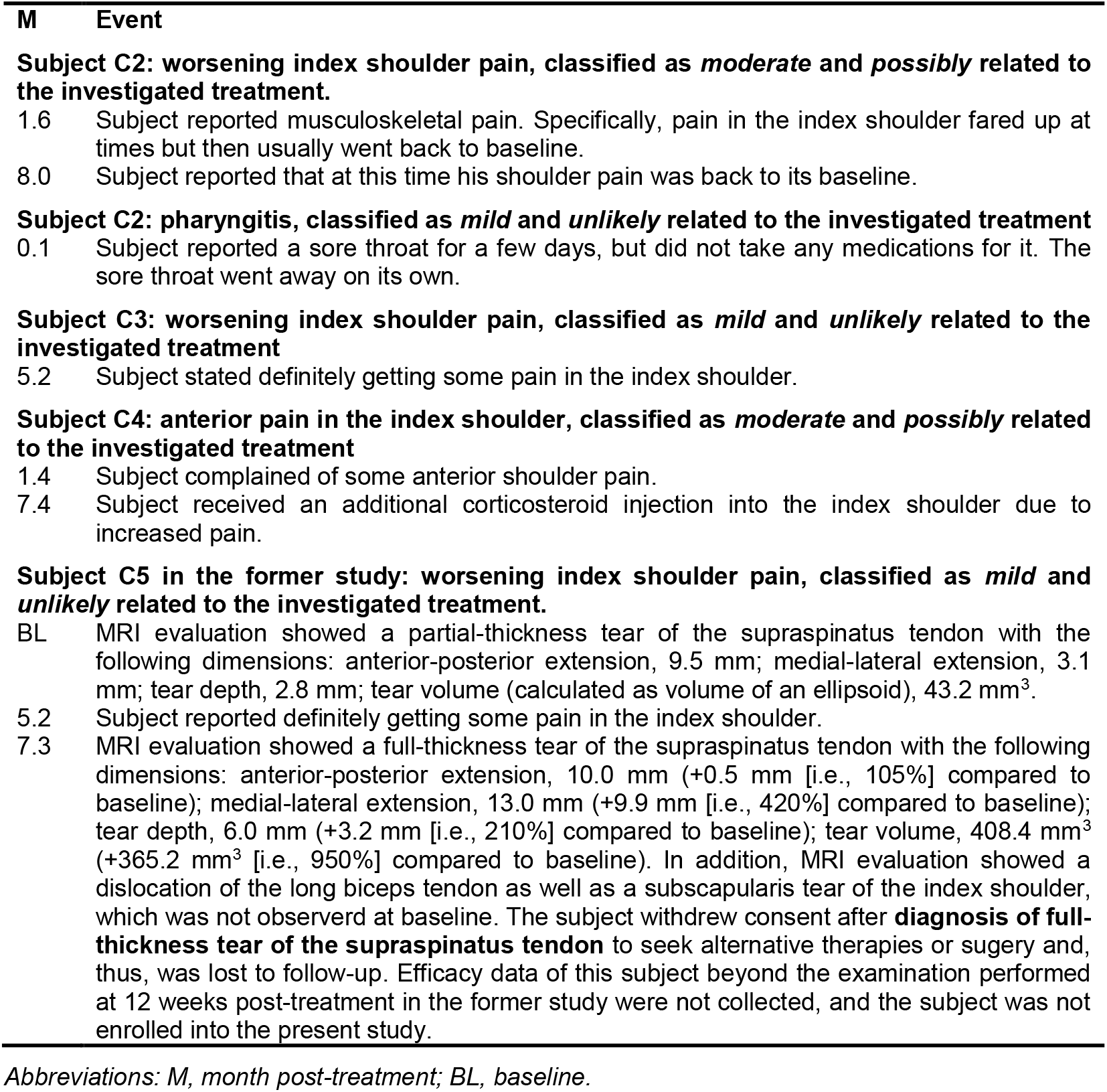
Individual courses of the treatment-related adverse events classified as {unlikely to be / possibly} related to the investigated treatment that occurred during the present and the former studies, experienced by those subjects who were treated with injection of corticosteroid.

| **M** | **Event** |
| --- | --- |
|  | **Subject C2: worsening index shoulder pain, classified as *moderate* and *possibly* related to the investigated treatment.** |
| 1.6 | Subject reported musculoskeletal pain. Specifically, pain in the index shoulder flared up at times but then usually went back to baseline. |
| 8.0 | Subject reported that at this time his shoulder pain was back to its baseline. |
|  | **Subject C2: pharyngitis, classified as *mild* and *unlikely* related to the investigated treatment** |
| 0.1 | Subject reported a sore throat for a few days, but did not take any medications for it. The sore throat went away on its own. |
|  | **Subject C3: worsening index shoulder pain, classified as *mild* and *unlikely* related to the investigated treatment** |
| 5.2 | Subject stated definitely getting some pain in the index shoulder. |
|  | **Subject C4: anterior pain in the index shoulder, classified as *moderate* and *possibly* related to the investigated treatment** |
| 1.4 | Subject complained of some anterior shoulder pain. |
| 7.4 | Subject received an additional corticosteroid injection into the index shoulder due to increased pain. |
|  | **Subject C5 in the former study: worsening index shoulder pain, classified as *mild* and *unlikely* related to the investigated treatment.** |
| BL | MRI evaluation showed a partial-thickness tear of the supraspinatus tendon with the following dimensions: anterior-posterior extension, 9.5 mm; medial-lateral extension, 3.1 mm; tear depth, 2.8 mm; tear volume (calculated as volume of an ellipsoid), 43.2 mm<sup>3</sup>. |
| 5.2 | Subject reported definitely getting some pain in the index shoulder. |
| 7.3 | MRI evaluation showed a full-thickness tear of the supraspinatus tendon with the following dimensions: anterior-posterior extension, 10.0 mm (+0.5 mm [i.e., 105%] compared to baseline); medial-lateral extension, 13.0 mm (+9.9 mm [i.e., 420%] compared to baseline); tear depth, 6.0 mm (+3.2 mm [i.e., 210%] compared to baseline); tear volume, 408.4 mm<sup>3</sup> (+365.2 mm<sup>3</sup> [i.e., 950%] compared to baseline). In addition, MRI evaluation showed a dislocation of the long biceps tendon as well as a subscapularis tear of the index shoulder, which was not observed at baseline. The subject withdrew consent after **diagnosis of full-thickness tear of the supraspinatus tendon** to seek alternative therapies or surgery and, thus, was lost to follow-up. Efficacy data of this subject beyond the examination performed at 12 weeks post-treatment in the former study were not collected, and the subject was not enrolled into the present study. |

*Abbreviations: M, month post-treatment; BL, baseline.*

Table S10 shows group-specific numbers of TEAEs that were classified as {mild and unlikely to be related to the investigated treatment / mild and possibly related to the investigated treatment / moderate and unlikely to be related to the investigated treatment / moderate and possibly related to the investigated treatment} in the present and the former studies.

**TABLE S10.** Group-specific numbers of TEAEs that were classified as {mild and unlikely to be related to the investigated treatment / mild and possibly related to the investigated treatment / moderate and unlikely to be related to the investigated treatment / moderate and possibly related to the investigated treatment} in the present and the former studies.

| TP | Group | Classification of TEAEs |  |  |  |
| --- | --- | --- | --- | --- | --- |
|  |  | mild and unlikely to be related | mild and possibly related | moderate and unlikely to be related | moderate and possibly related |
| BL-W24 | UA-ADRCs | 2 | 3 | 2 | 1 |
| BL-FSV | UA-ADRCs | 4 | 3 | 2 | 1 |
| BL-SSV | UA-ADRCs | 4 | 3 | 2 | 1 |
| BL-W24 | C | 3 | 0 | 0 | 2 |
| BL-FSV | C | 3 | 0 | 0 | 2 |
| BL-SSV | C | 3 | 0 | 0 | 2 |

*Abbreviations: TP, time period; BL, baseline; W24, study visit scheduled in the former study at 24 weeks post-treatment; FSV, first study visit of the present study; SSV, second study visit of the present study; UA-ADRCs, treatment with a single injection of fresh, uncultured, unmodified, autologous, adipose-derived regenerative cells; C, treatment with a single injection of corticosteroid.*

For all investigated time periods there was no statistically significant difference between the groups with regard to the numbers of TEAEs that were classified as {mild and unlikely to be related to the investigated treatment / mild and possibly related to the investigated treatment / moderate and unlikely to be related to the investigated treatment / moderate and possibly related to the investigated treatment} in the present and the former studies (Chi-square test for trend):

- from baseline to W24 in the former study: p = 0.941,
- from baseline to the first study visit of the present study: p = 0.757, and
- from baseline to the second study visit of the present study: p = 0.757.

#### 2.3. Severe treatment emergent adverse events

Tables S11–S13 summarize the individual courses of the four severe TEAEs that occurred during the present and the former studies, all of which were classified as not related to the investigated treatment.

**TABLE S11.** Severe TEAE experienced by Subject A7 who was treated with UA-ADRCs.

| **M** | **Event** |
| --- | --- |
| 5.5 | Subject reported acute pain in the left shoulder (i.e., not in the index shoulder). A subacromial bursitis impingement and a small undersurface tear of the rotator cuff were diagnosed, but no treatment was performed. |
| 34.4 | Subject was diagnosed with a **spinoglenoid notch cyst in the left shoulder (i.e., not in the index shoulder) (severe TEAE experienced by this subject)**, discovered by MRI. Pain at times radiated up towards the neck, and at times down the arm with some pain all the way into the hand. |
| 36.9 | Arthroscopic decompression of the cyst and repair of the posterior capsulolabral tissue was performed. |
| 37.4 | Subject reported pain 4/10 and occasional use of Tylenol and ice, as well as use of a SlingShot immobilizer. There was still some swelling present at this time, but this was decreasing. |
| 38.3 | There was still some discomfort, together with slow progress in improvement of range of motion. |
| 40.3 | The pain was nearly resolved and restricted to holding the left arm at shoulder height for extended periods of time. External rotation was lacking about 10 degrees, and internal rotation was lacking about two to three interspace levels. |
| 41.2 | Subject reported that the stopped physical therapy due to increased pain when using heavy weights. |

*Abbreviations: UA-ADRCs, treatment with a single injection of fresh, uncultured, unmodified, autologous, adipose-derived regenerative cells; M, month post-treatment.*

Data regarding the SAE of Subject A7 (this SAE occurred during the present study) was sent to the Institutional Review Board of the study site, the Data and Safety Monitoring Board of the present study and the U.S. Food and Drug Administration, and all noted that this SAE was not related to study treatment. The SAE was treated per standard of care.

**TABLE S12.**
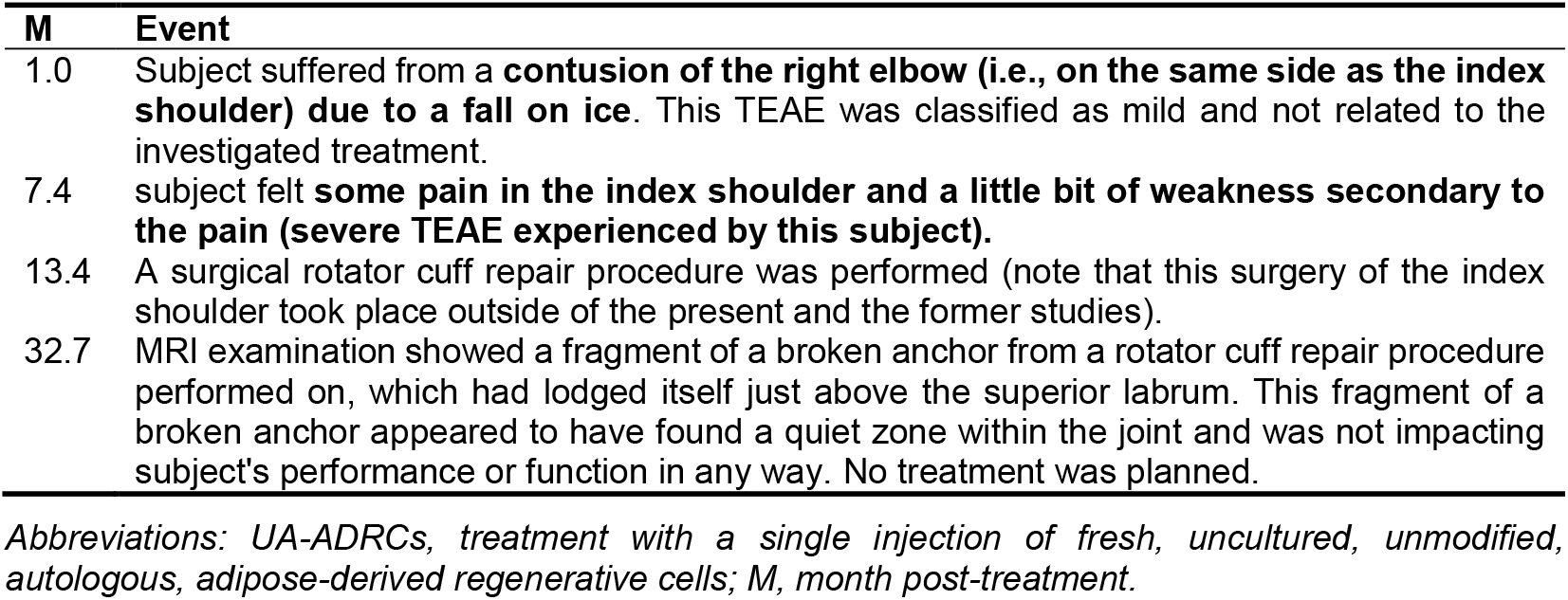
Severe TEAE experienced by Subject A9 who was treated with UA-ADRCs.

| M | Event |
| --- | --- |
| 1.0 | Subject suffered from a **contusion of the right elbow (i.e., on the same side as the index shoulder) due to a fall on ice**. This TEAE was classified as mild and not related to the investigated treatment. |
| 7.4 | subject felt **some pain in the index shoulder and a little bit of weakness secondary to the pain (severe TEAE experienced by this subject)**. |
| 13.4 | A surgical rotator cuff repair procedure was performed (note that this surgery of the index shoulder took place outside of the present and the former studies). |
| 32.7 | MRI examination showed a fragment of a broken anchor from a rotator cuff repair procedure performed on, which had lodged itself just above the superior labrum. This fragment of a broken anchor appeared to have found a quiet zone within the joint and was not impacting subject's performance or function in any way. No treatment was planned. |

Abbreviations: UA-ADRCs, treatment with a single injection of fresh, uncultured, unmodified, autologous, adipose-derived regenerative cells; M, month post-treatment.

**TABLE S13.** Severe TEAEs experienced by Subject A4 who was treated with UA-ADRCs.

| M | Event |
| --- | --- |
| 3.0 | Subject was diagnosed with a **non ST elevation myocardial infarction (first severe TEAE experienced by this subject)**. The subject had some recurring pain intermittently, but had been stable while awaiting catheter examination over the weekend. The decision to wait for catheter examination was due to platelet count of 50,000. Stress testing on the weekend indicated inferior ischemia. |
| 3.1 | ACE-inhibitor was stopped and i.v. fluids were given. Initially the subject was doing well, but developed chest pain when showering; ECG was suggestive of ST elevation. The subject was taken emergently to the catheter lab on the same day. Stents were placed in a previous vein graft to the right posterior descending artery (RPDA) and the native RPDA. The decision to split treatment into two procedures was due to concerns with his kidneys. |
| 3.1 | Stents were also placed in a previous vein graft to the lateral left anterior descending artery. The subject did well on the rest of his stay and was discharged a few days later. A cardiology follow up was planned. Atenolol was also stopped due to bradycardia concerns. |
| 4.3 | Subject had some feelings of fatigue and has noted some left sided chest discomfort and left arm pain into the antecubital area with walking and relieved with rest. Treatment with Imdur daily was started; a stress test was abnormal. The subject was diagnosed with a **ST elevation myocardial infarction (second severe TEAE experienced by this subject)**. |
| 4.6 | A cardiac angiogram showed no lesions amenable to revascularization. Adjustments to current medications were performed. |
| 6.3 | A follow up doctor visit showed that the subject was symptomatically doing well with medication changes. |

Abbreviations: UA-ADRCs, treatment with a single injection of fresh, uncultured, unmodified, autologous, adipose-derived regenerative cells; M, month post-treatment.

### Part 3 Scheduled study visits and availability of ASES Total score, VAS pain score and SF-36 Total score data

Table S14 lists the scheduled visits of the present and the former studies, during which the primary endpoint *long term efficacy of pain and function through ASES Shoulder Score and SF-36 health questionnaires* between the two groups was investigated, and indicates whether (+) or not (−) the corresponding subject developed additional pathologies of the index shoulder (next to symptomatic, partial-thickness rotator cuff tear) and/or received additional treatments on the index shoulder (next to injection of UA-ADRCs or corticosteroid) during either of these studies. The corresponding reasons are summarized in Table S15.

**TABLE S14.**
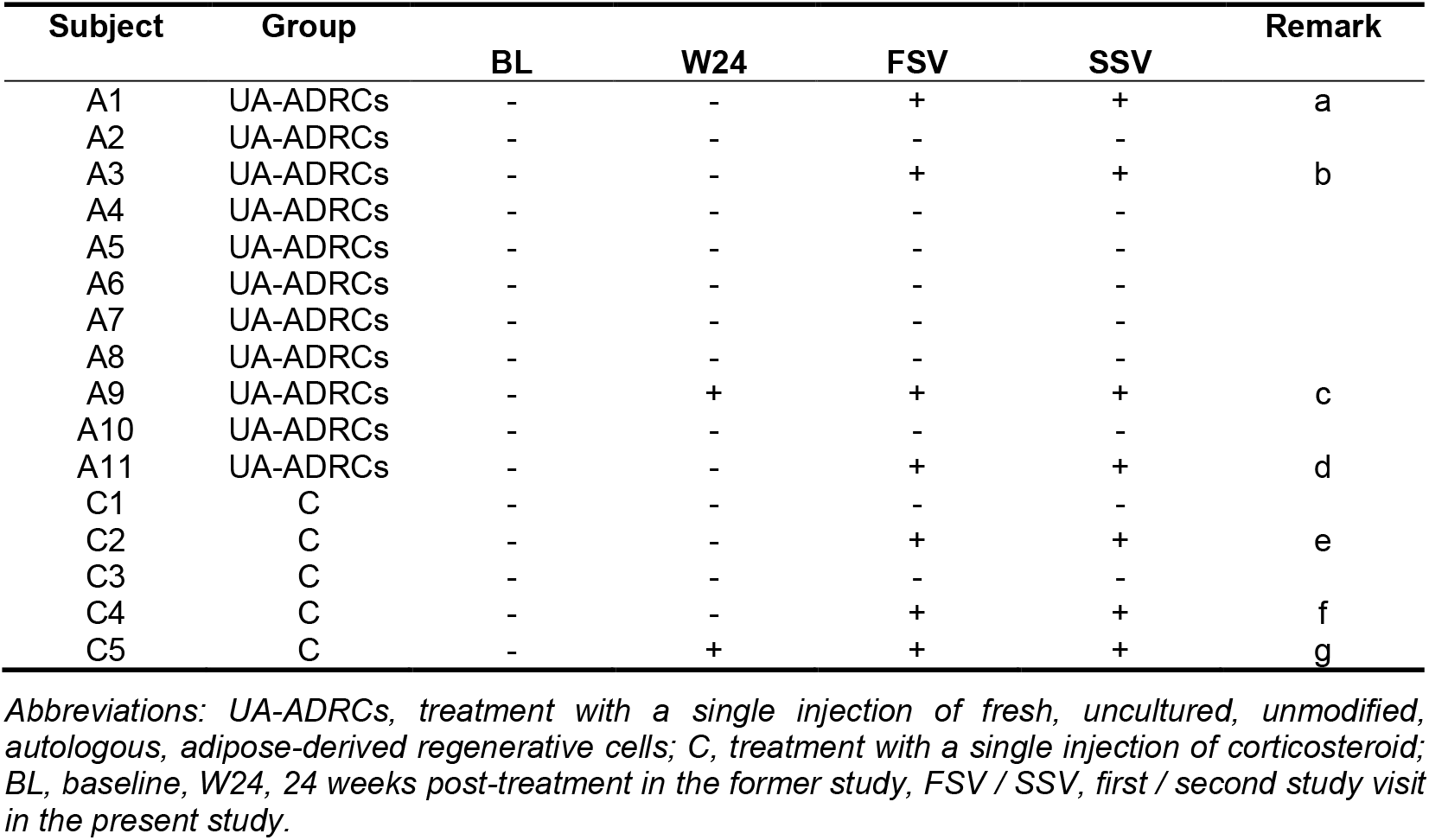
Scheduled visits of the present and the former studies, during which the primary endpoint long term efficacy of pain and function through ASES Shoulder Score and SF-36 health questionnaires between the two groups was investigated, and indication whether (+) or not (−) the corresponding subject developed additional pathologies of the index shoulder (next to symptomatic, partial-thickness rotator cuff tear) and/or received additional treatments on the index shoulder (next to injection of UA-ADRCs or corticosteroid) during either of these studies. The remarks are outlined in Table S15.

**TABLE S15.**
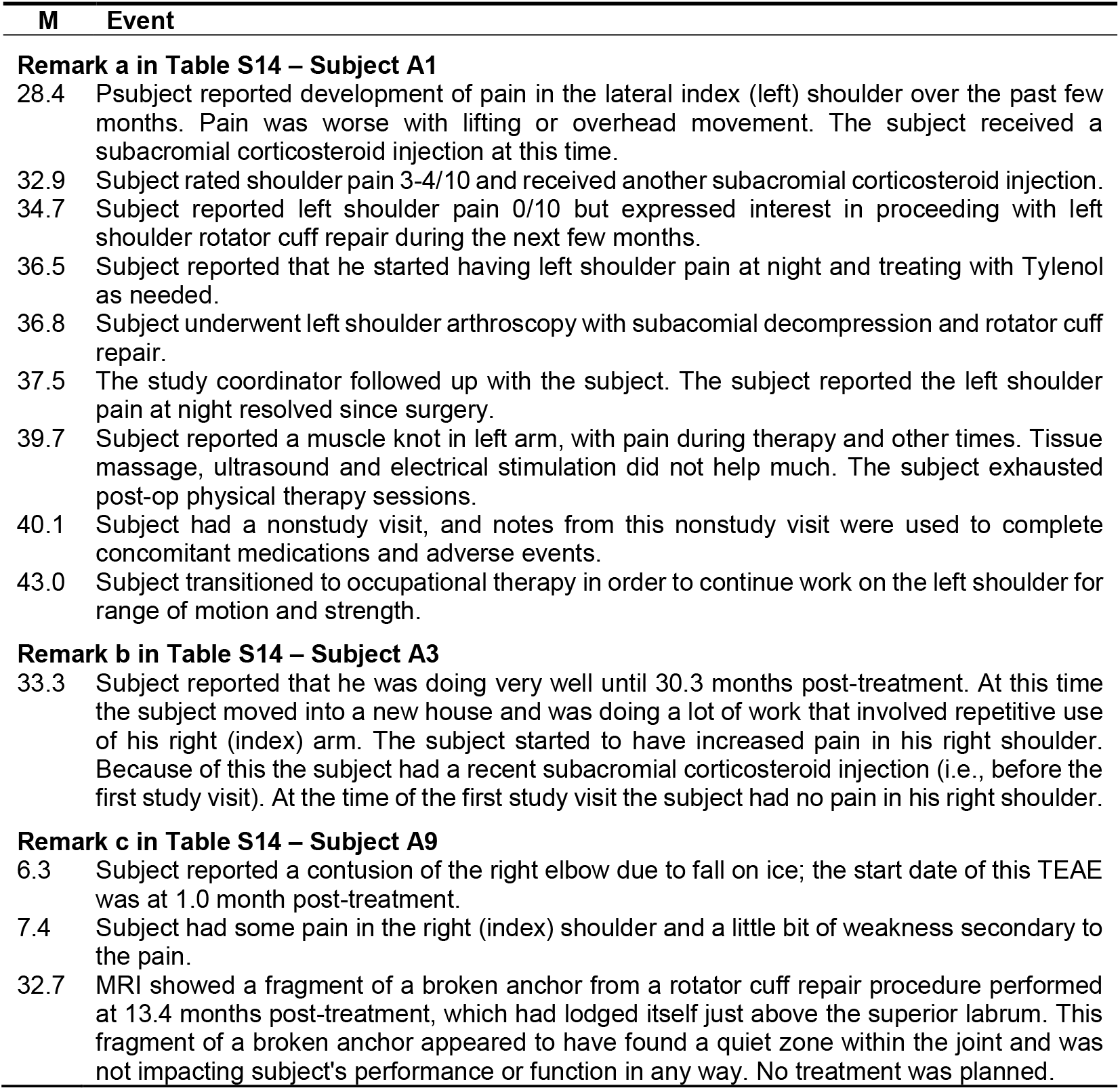
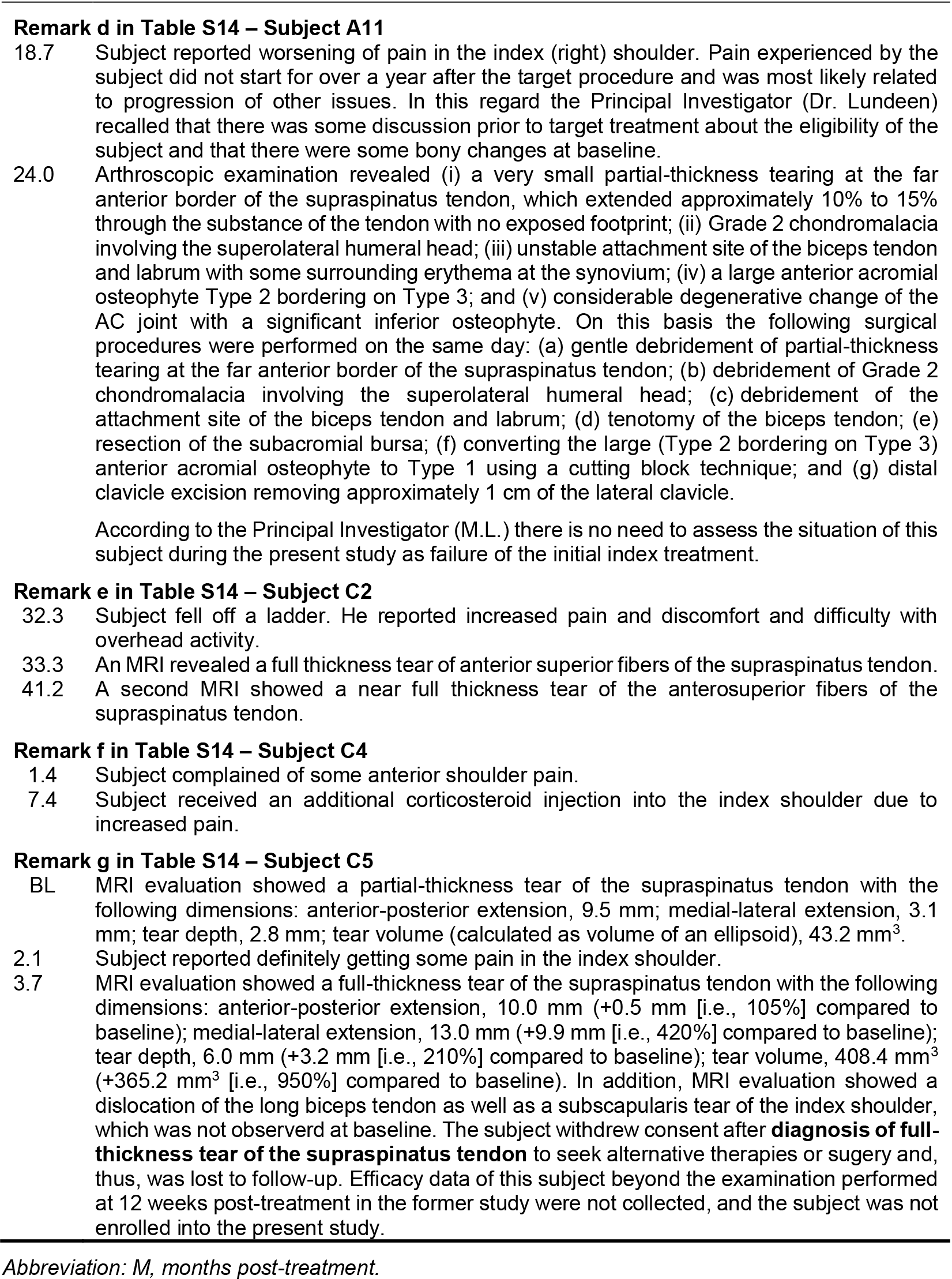
Reasons why individual subjects enrolled in the present and the former studies developed additional pathologies of the index shoulder (next to symptomatic, partial-thickness rotator cuff tear) and/or received additional treatments on the index shoulder (next to injection of UA-ADRCs or corticosteroid) during either of these studies. The remarks refer to Table S14.

### Part 4 Efficacy data as a function of time post-treatment

**FIGURE S16.**
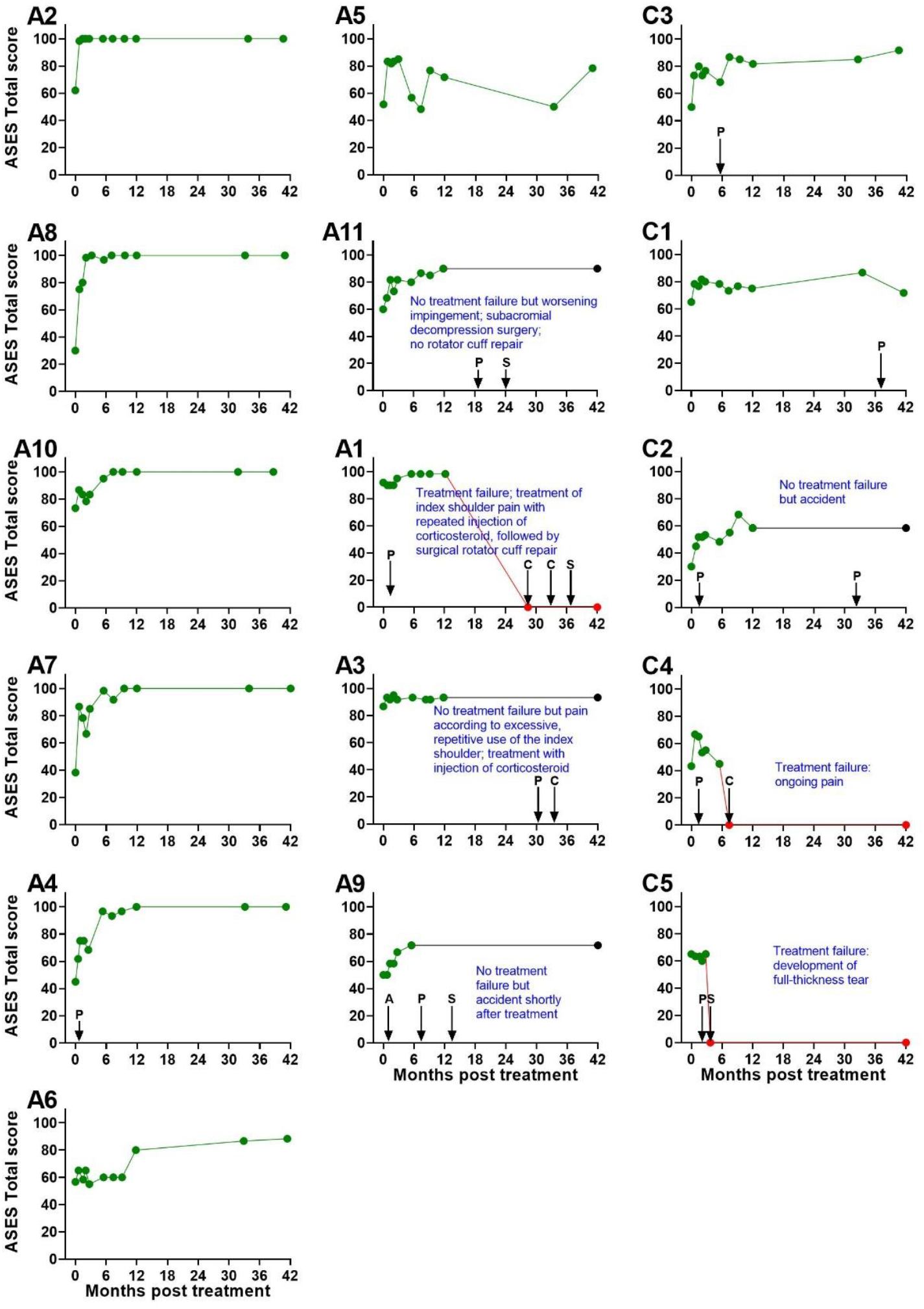
Individual ASES Total score data as a function of time post-treatment of subjects treated with injection of either UA-ADRCs (Subjects A1-A11) or corticosteroid (Subjects C1-C5). The data are arranged in descending order of individual treatment success (i.e., the data of the subjects with the hightest ASES Total score are shown in the top row of the left column (Subject A2 treated with injection of UA-ADRCs) and the right column (Subject C3 treated with injection of corticosteroid). In the graphs…

- green dots and green lines indicate data of the present and the former studies that were available and could be used to assess treatment outcome,
- black dots and black lines (Subjects A11, A13 and C2) indicate data of the present study that were imputed uisng the Last Observation Carried Forward approach and could be used to assess treatment outcome (reasons are indicated in the corresponding panels and outlined in detail in Table S15), and
- red dots and red lines (Subjects A1, C4 and C5) indicate data of the present and the former studies that were imputed as “failures” (reasons are indicated in the corresponding panels and outlined in detail in Table S15). Abbreviations: A, accident affecting the index shoulder; C, injection of corticosteroid into the index shoulder; P, pain in the index shoulder; S, surgery of the index shoulder. Abbreviations: A, accident affecting the index shoulder; C, injection of corticosteroid into the index shoulder; P, pain in the index shoulder; S, surgery of the index shoulder.

**FIGURE S17.**
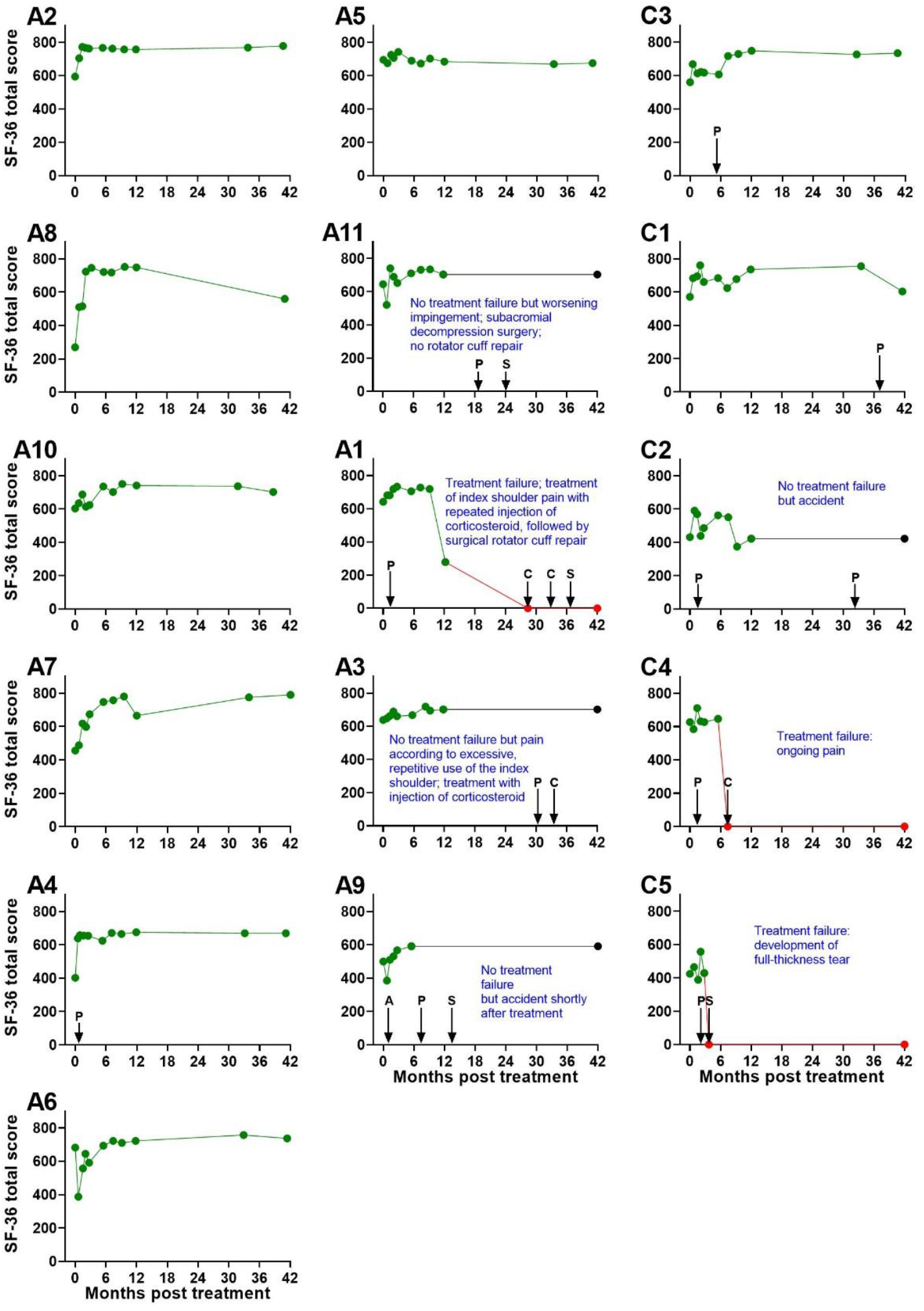
Individual SF-36 Total score as a function of time post-treatment of subjects treated with injection of either UA-ADRCs (Subjects A1-A11) or corticosteroid (Subjects C1-C5). The data are arranged in descending order of individual treatment success (i.e., the data of the subjects with the hightest <u>ASES Total scores</u> are shown in the top row of the left column (Subject A2 treated with injection of UA-ADRCs) and the right column (Subject C3 treated with injection of corticosteroid) (c.f. Figure 16). In the graphs… • green dots and green lines indicate data of the present and the former studies that were available and could be used to assess treatment outcome, • black dots and black lines (Subjects A11, A13 and C2) indicate data of the present study that were imputed uisng the Last Observation Carried Forward approach and could be used to assess treatment outcome (reasons are indicated in the corresponding panels and outlined in detail in Table S15), and • red dots and red lines (Subjects A1, C4 and C5) indicate data of the present and the former studies that were imputed as “failures” (reasons are indicated in the corresponding panels and outlined in detail in Table S15). Abbreviations: A, accident affecting the index shoulder; C, injection of corticosteroid into the index shoulder; P, pain in the index shoulder; S, surgery of the index shoulder. Abbreviations: A, accident affecting the index shoulder; C, injection of corticosteroid into the index shoulder; P, pain in the index shoulder; S, surgery of the index shoulder.

**FIGURE S18.**
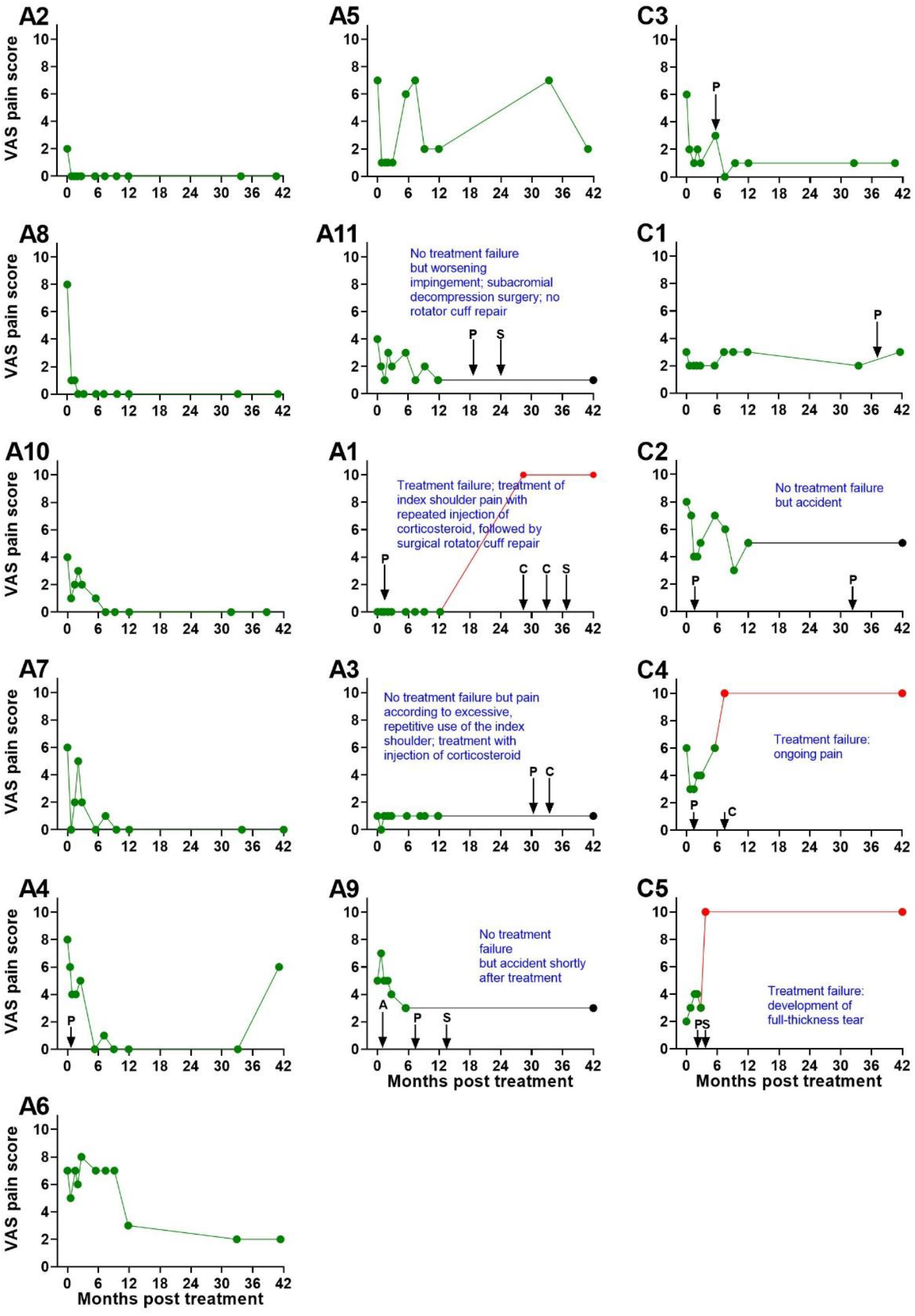
Individual VAS pain score (collected together with the ASES score) as a function of time post-treatment of subjects treated with injection of either UA-ADRCs (Subjects A1-A11) or corticosteroid (Subjects C1-C5). The data are arranged in descending order of individual treatment success (i.e., the data of the subjects with the hightest <u>ASES Total scores</u> are shown in the top row of the left column (Subject A2 treated with injection of UA-ADRCs) and the right column (Subject C3 treated with injection of corticosteroid) (c.f. Figure 16). In the graphs… • green dots and green lines indicate data of the present and the former studies that were available and could be used to assess treatment outcome, • black dots and black lines (Subjects A11, A13 and C2) indicate data of the present study that were imputed uisng the Last Observation Carried Forward approach and could be used to assess treatment outcome (reasons are indicated in the corresponding panels and outlined in detail in Table S15), and • red dots and red lines (Subjects A1, C4 and C5) indicate data of the present and the former studies that were imputed as “failures” (reasons are indicated in the corresponding panels and outlined in detail in Table S15). Abbreviations: A, accident affecting the index shoulder; C, injection of corticosteroid into the index shoulder; P, pain in the index shoulder; S, surgery of the index shoulder.

**TABLE S16.** Mean, standard error of the mean and median of the ASES Total score collected during the present and the former studies after treating subjects suffering from sPTRCT with injection of either UA-ADRCs or corticosteroid.

| Time point | Subjects treated with injection of UA-ADRCs |  | Subjects treated with injection of corticosteroid |  | D (U/C) | p |
| --- | --- | --- | --- | --- | --- | --- |
| | Mean ± SEM | Median | Mean ± SEM | Median | | |
| BL | 58.7 ± 5.8 | 56.7 | 50.6 ± 6.7 | 50 | +16.0% | 0.569 |
| W24 | 86.1 ± 4.9 | 95 | 48.0 ± 13.5 | 48.3 | +79.4% | 0.008 |
| W52 | 89.4 ± 4.9 | 98.3 | 43.0 ± 17.7 | 58.3 | +107.9% | 0.011 |
| FSV | 79.6 ± 9.9 | 98.3 | 46.0 ± 19.4 | 58.3 | +72.9% | 0.052 |
| SSV | 82.3 ± 9.4 | 98.3 | 44.3 ± 18.9 | 58.3 | +85.5 | 0.048 |

Abbreviations: sPTRCT, symptomatic, partial-thickness rotator cuff tear; UA-ADRCs, fresh, uncultured, unmodified, autologous, adipose-derived regenerative cells; D (U/C), ratio of mean values between the subjects in the UA-ADRCs group and the subjects in the corticosteroid group; p, result (p value) of Mann-Whitney test; SEM, standard error of the mean; BL, baseline; W24 / W52, study visits scheduled in the former study at 24 and 52 weeks post-treatment; FSV, first study visit of the present study; SSV, second study visit of the present study.

**TABLE S17.** Mean, standard error of the mean and median of the SF-36 Total score collected during the present and the former studies after treating subjects suffering from sPTRCT with injection of either UA-ADRCs or corticosteroid.

| Time point | Subjects treated with injection of UA-ADRCs |  | Subjects treated with injection of corticosteroid |  | D (U/C) | p |
| --- | --- | --- | --- | --- | --- | --- |
| | Mean ± SEM | Median | Mean ± SEM | Median | | |
| BL | 557 ± 40.4 | 603.7 | 523 ± 40.4 | 560 | +6.5% | 0.320 |
| W24 | 696 ± 15.7 | 705.5 | 500 ± 127 | 606.5 | +39.1% | 0.013 |
| W52 | 654 ± 43.9 | 701.5 | 381 ± 166 | 422.7 | +71.5% | 0.307 |
| FSV | 602 ± 75.6 | 732 | 381 ± 166 | 422.7 | +58.0% | 0.220 |
| SSV | 582 ± 73.8 | 675.5 | 352 ± 152 | 422.7 | +65.4% | 0.179 |

Abbreviations: sPTRCT, symptomatic, partial-thickness rotator cuff tear; UA-ADRCs, fresh, uncultured, unmodified, autologous, adipose-derived regenerative cells; D (U/C), ratio of mean values of the subjects in the UA-ADRCs group and the subjects in the corticosteroid group; p, result (p value) of Mann-Whitney test; SEM, standard error of the mean; BL, baseline; W24 / W52, study visits scheduled in the former study at 24 and 52 weeks post treatment; FSV, first study visit of the present study; SSV, second study visit of the present study.

**TABLE S18.** Mean, standard error of the mean and median of the VAS Pain score (collected together with the ASES Total score) collected during the present and the former studies after treating subjects suffering from sPTRCT with injection of either UA-ADRCs or corticosteroid.

| Time point | Subjects treated with injection of UA-ADRCs |  | Subjects treated with injection of corticosteroid |  | D (U/C) | p |
| --- | --- | --- | --- | --- | --- | --- |
| | Mean ± SEM | Median | Mean ± SEM | Median | | |
| BL | 4.7 ± 0.8 | 5 | 5.0 ± 1.1 | 6 | -5.5% | 0.930 |
| W24 | 1.9 ± 0.8 | 1 | 5.6 ± 1.4 | 6 | -65.9% | 0.031 |
| W52 | 1.1 ± 0.5 | 0 | 5.8 ± 1.8 | 5 | -81.2% | 0.009 |
| FSV | 2.3 ± 1.1 | 0 | 5.6 ± 1.9 | 5 | -59.4% | 0.059 |
| SSV | 2.4 ± 1.0 | 1 | 5.8 ± 1.8 | 5 | -59.2% | 0.080 |

Abbreviations: sPTRCT, symptomatic, partial-thickness rotator cuff tear; UA-ADRCs, fresh, uncultured, unmodified, autologous, adipose-derived regenerative cells; D (U/C), ratio of mean values between the subjects in the UA-ADRCs group and the subjects in the corticosteroid group; p, result (p value) of Mann-Whitney test; SEM, standard error of the mean; BL, baseline; W24 / W52, study visits scheduled in the former study at 24 and 52 weeks post treatment; FSV, first study visit of the present study; SSV, second study visit of the present study.

### Part 5 Analysis of MRI scans

**FIGURE S19.**
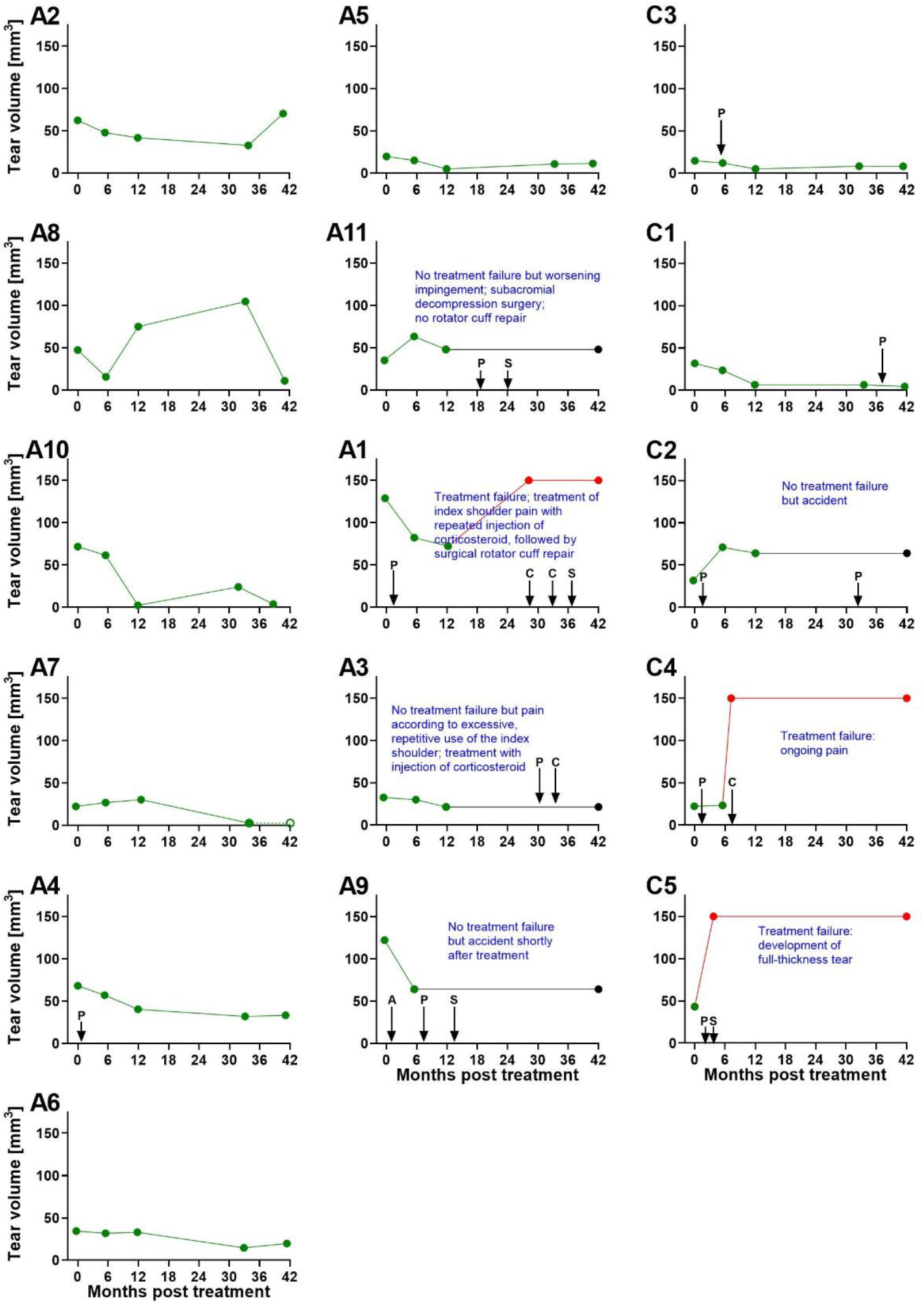
Individual tear size (calculated as ellipsoid volume) as a function of time post-treatment of subjects treated with injection of either UA-ADRCs (Subjects A1-A11) or corticosteroid (Subjects C1-C5). The data are arranged in descending order of individual treatment success (i.e., the data of the subjects with the hightest <u>ASES Total scores</u> are shown in the top row of the left column (Subject A2 treated with injection of corticosteroid) and the right column (Subject C3 treated with injection of UA-ADRCs) (c.f. Figure S16). In the graphs… • green dots and green lines indicate data of the present and the former studies that were available and could be used to assess treatment outcome, • black dots and black lines (Subjects A11, A13 and C2) indicate data of the present study that were imputed uisng the Last Observation Carried Forward approach and could be used to assess treatment outcome (reasons are indicated in the corresponding panels and outlined in detail in Table S15), and • red dots and red lines (Subjects A1, C4 and C5) indicate data of the present and the former studies that were imputed as “failures” (reasons are indicated in the corresponding panels and outlined in detail in Table S15). Abbreviations: A, accident affecting the index shoulder; C, injection of corticosteroid into the index shoulder; P, pain in the index shoulder; S, surgery of the index shoulder.

**TABLE S19.** Mean, standard error of the mean and median of the tear size (calculated as ellipsoid volume) collected during the present and the former studies after treating subjects suffering from sPTRCT with injection of either UA-ADRCs or corticosteroid.

| Time point | Subjects treated with injection of UA-ADRCs |  | Subjects treated with injection of corticosteroid |  | D (U/C) | p |
| --- | --- | --- | --- | --- | --- | --- |
| | Mean ± SEM<br>[mm<sup>3</sup>] | Median<br>[mm<sup>3</sup>] | Mean ± SEM<br>[mm<sup>3</sup>] | Median<br>[mm<sup>3</sup>] | | |
| BL | 58.6 ± 11.3 | 47.3 | 28.7 ± 4.8 | 31.7 | +104% | 0.055 |
| W24 | 45.0 ± 6.8 | 47.7 | 55.9 ± 25.6 | 23.5 | -19.6% | 0.827 |
| W52 | 44.5 ± 10.3 | 40.5 | 75.1 ± 32.4 | 63.9 | -40.7% | 0.564 |
| FSV | 55.7 ± 14.9 | 32.6 | 75.7 ± 32.1 | 63.9 | -26.5% | 0.827 |
| SSV | 49.3 ± 14.9 | 33.3 | 75.3 ± 32.3 | 63.9 | -34.5% | 0.660 |

Abbreviations: sPTRCT, symptomatic, partial-thickness rotator cuff tear; UA-ADRCs, fresh, uncultured, unmodified, autologous, adipose-derived regenerative cells; D (U/C), ratio of mean values of subjects in the UA-ADRCs group and subjects in the corticosteroid group; p, result (p value) of Mann-Whitney test; SEM, standard error of the mean; BL, baseline; W24 / W52, study visits scheduled in the former study at 24 and 52 weeks post treatment; FSV, first study visit of the present study; SSV, second study visit of the present study.

### Part 6 Relationship between treatment outcome and baseline data

**FIGURE S20.**
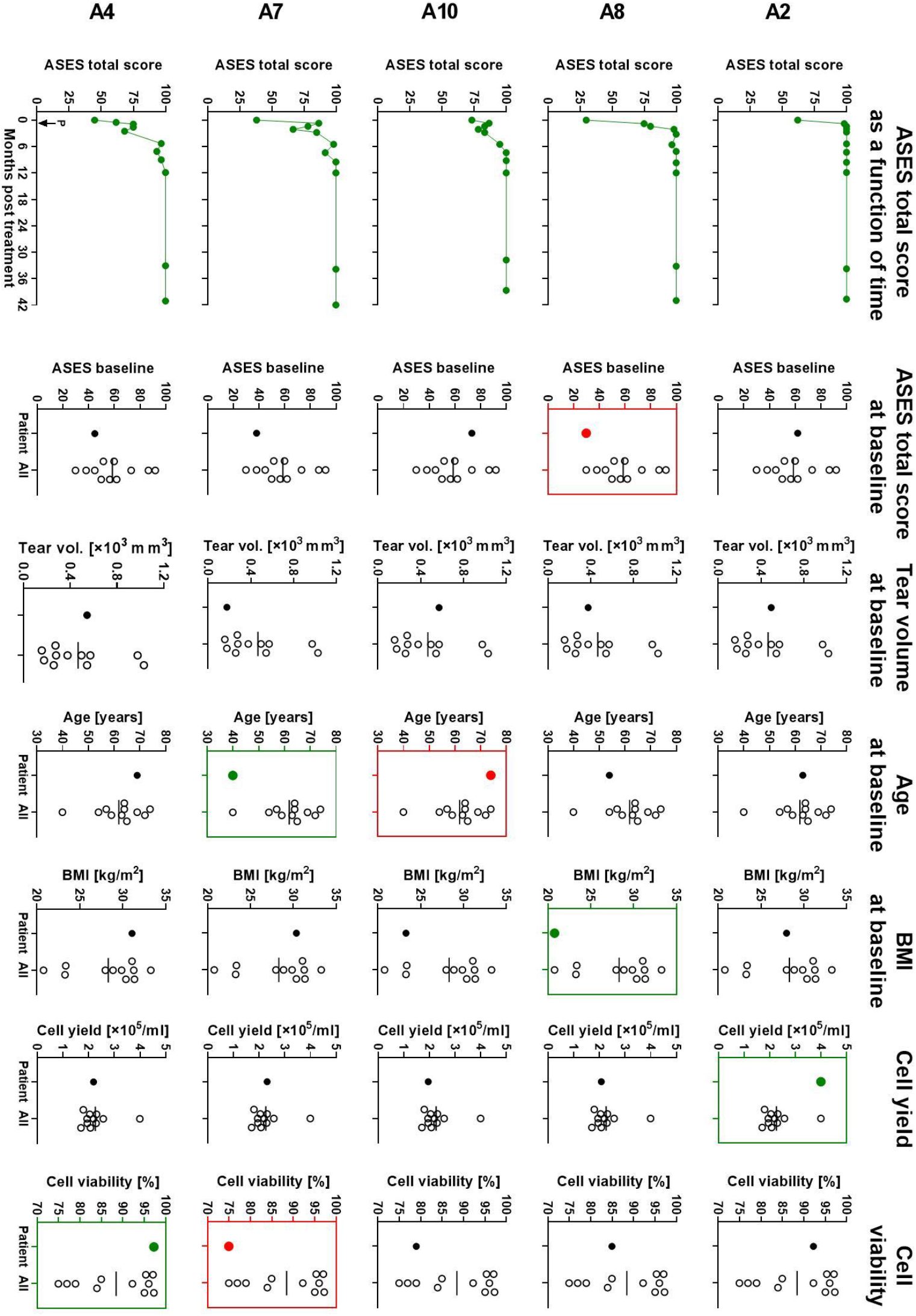
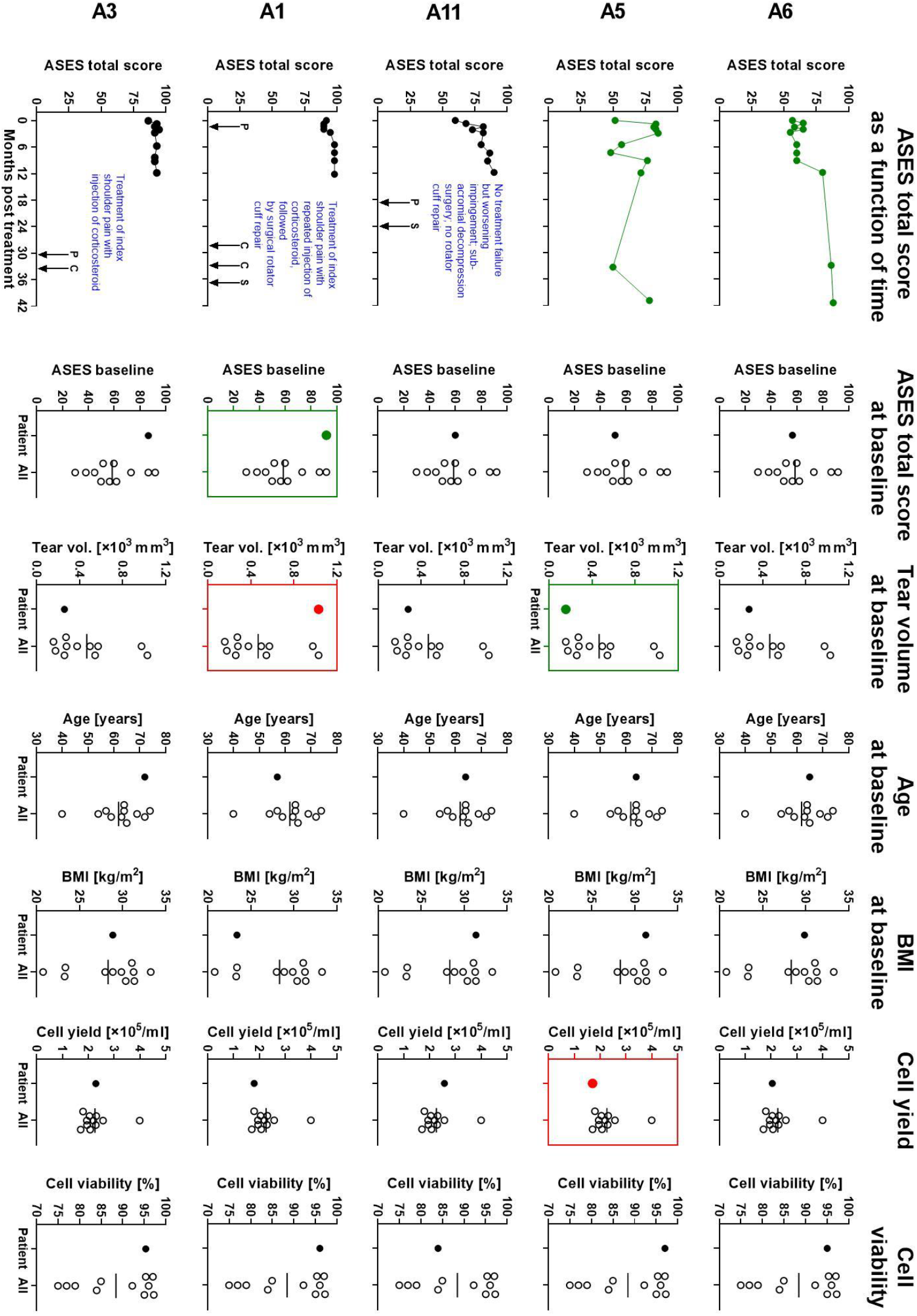
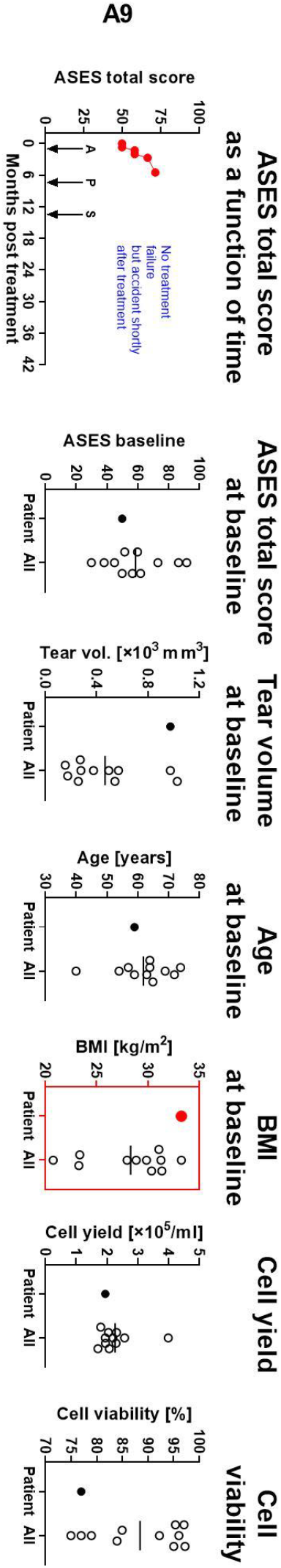
Individual ASES Total score as a function of time post-treatment, and individual data at baseline (ASES Total score, tear volume, age, body mass index, cell yield and cell viability) of the subjects who were treated with injection of UA-ADRCs (Subjects A1-A11). The data are arranged in descending order of individual treatment success (i.e., the data of the subject with the best treatment outcome (Subject A2) are shown in the top row). In the graphs showing individual ASES Total scores as a function of time post-treatment… • green dots and green lines (Subjects A2, A4, A5, A6, A7, A8 and A10) indicate that all data of the present and the former studies were available and could be used for assessing treatment outcome, • black dots and black lines (Subjects A1, A3 and A11) indicate that all data of the former study were available and could be used for assessing treatment outcome, but in the time period between the present and the former studies there was an incidence that rendered the data of the present study unsuitable for assessing treatment outcome (reasons are indicated in the corresponding panels and outlined in detail in Table S15), and • red dots and red lines (Subject A9) indicate that there was an incidence during the former study that rendered a part of the data of the former study (and, thus, all data of the present study in case they were collected) unsuitable for assessing treatment outcome (reasons are indicated in the corresponding panel and outlined in detail in Table S15). In the graphs showing individual data at baseline (ASES Total score, tear volume, age, body mass index, cell yield and cell viability)… • the individual data of the subject whose ASES Total score as a function of time post-treatment is shown in the same row are given on the left (“Subject”), and • the data of all subjects in the corticosteroid group are given on the right (“All”). The horizontal lines represent mean values. In addition, for each variable the subject treated with UA-ADRCs with… • presumably the best prognosis (at baseline highest ASES Total score, smallest tear volume, youngest age and lowest body mass index, as well as highest cell yield and highest cell viability) is indicated with a green dot and green frame surrounding the corresponding data, and • presumably the worst prognonis (at baseline lowest ASES Total score, largest tear volume, oldest age and highest body mass index, as well as lowest cell yield and lowest cell viability) is indicated with a red dot and red frame surrounding the corresponding data. Abbreviations: A, accident affecting the index shoulder; C, injection of corticosteroid into the index shoulder; P, pain in the index shoulder; S, surgery of the index shoulder.

**FIGURE S21.**
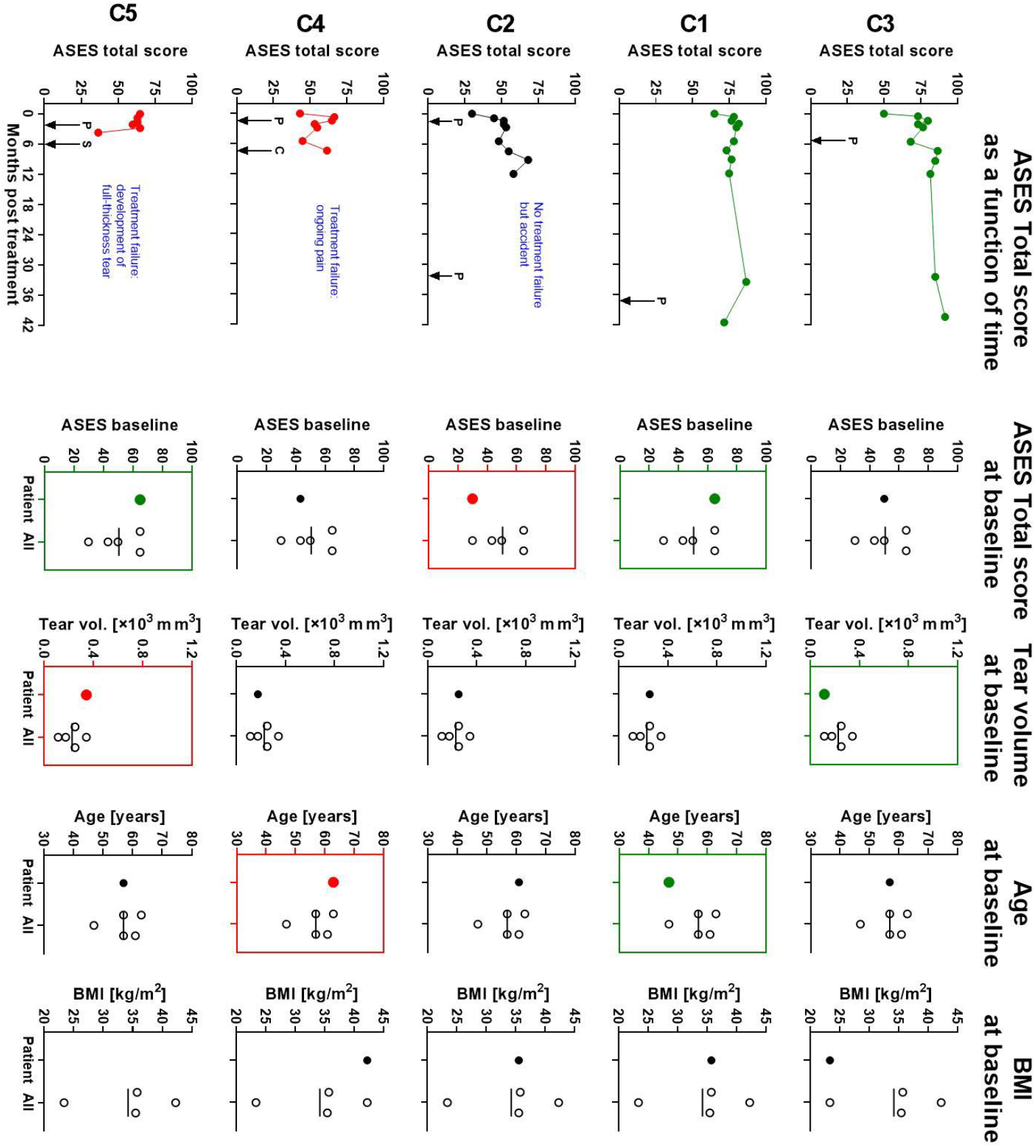
Individual ASES Total score as a function of time post-treatment, and individual data at baseline (ASES Total score, tear volume, age and body mass index) of the subjects who were treated with corticosteroid. The data are arranged in descending order of individual treatment success (i.e., the data of the subject with the best treatment outcome (C3) are shown in the top row). In the graphs showing individual ASES Total scores as a function of time post-treatment… • green dots and green lines (Subjects A2, A4, A5, A6, A7, A8 and A10) indicate that all data of the present and the former studies were available and could be used for assessing treatment outcome, • black dots and black lines (Subjects A1, A3 and A11) indicate that all data of the former study were available and could be used for assessing treatment outcome, but in the time period between the present and the former studies there was an incidence that rendered the data of the present study unsuitable for assessing treatment outcome (reasons are indicated in the corresponding panels and outlined in detail in Table S15), and • red dots and red lines (Subject A9) indicate that there was an incidence during the former study that rendered a part of the data of the former study (and, thus, all data of the present study in case they were collected) unsuitable for assessing treatment outcome (reasons are indicated in the corresponding panel and outlined in detail in Table S15). In the graphs showing individual data at baseline (ASES Total score, tear volume, age and body mass index)… • the individual data of the subject whose ASES Total score as a function of time post-treatment is shown in the same row are given on the left (“Subject”), and • the data of all subjects in the corticosteroid group are given on the right (“All”). The horizontal lines represent mean values. In addition, for each variable the subject treated with corticosteroid with… • presumably the best prognosis (at baseline highest ASES Total score, smallest tear volume, youngest age and lowest body mass index) is indicated with a green dot and green frame surrounding the corresponding data, and • presumably the worst prognonis (at baseline lowest ASES Total score, largest tear volume, oldest age and highest body mass index) is indicated with a red dot and red frame surrounding the corresponding data (note that this was not possible in case of the BMI at baseline because for Subject C5 no height was collected in the former study and, thus, no BMI could be calculated). Abbreviations: P, pain in the index shoulder; C, injection of corticosteroid into the index shoulder; S, surgery of the index shoulder.

### Part 7 Comparison of the results obtained in the present and the former studies after treatment of sPTRCT with injection of corticosteroid with corresponding results of other studies in the literature

**TABLE S20.** Details of published studies that investigated the efficacy of treating partial-thickness rotator cuff tears with injections of corticosteroids. References are in the main text.

| Study | Treatment | HE | HED | Additional injection of local anesthetic |
| --- | --- | --- | --- | --- |
| [12] | 2 mL 40 mg/ml methylprednisolone | 5 | 1 x 400 | 3 mL of 0.25% bupivacaine |
| [39] | 1 ml of 6 mg betamethasone | 25 | 1 x 150 | 4 mL of 2% xylocaine |
| [40] | 5 ml of 40 mg/ml triamcinolone acetonide* | 5 | 1 x 1000 (?)<sup>a</sup> | -- |
| [41] | 1mL of 40 mg/mL methylprednisolone acetate | 5 | 1 x 200 | 1mL of 1% lignocaine hydrochloride |
| [42] | 7 mg betamethasone (3x, 1x/week) | 25 | 3 x 175 | -- |
| [43] | 2 mL of 40 mg/ml triamcinolone acetonide | 5 | 1 x 400 | 2 mL of 1% lidocaine |

*Abbreviations: HE, hydrocortisone equivalent (https://clincalc.com/Corticosteroids/); HED, hydrocortisone equivalent dose [mg].*
*<sup>a</sup>, incomplete description of dosage ("patients received a cortisone injection [Dermapharm Kenacort® 40 mg (triamcinolone acetonide, crystal suspension)] by means of a 5-ml syringe under aseptic conditions.").*

**TABLE S21.** Results of the studies summarized in Table S20. References are in the main text.

| Study | VAS pain score |  |  |  |  |  | ASES total score |  |  |  |  |  |
| --- | --- | --- | --- | --- | --- | --- | --- | --- | --- | --- | --- | --- |
|  | n | BL Mean | SD | M6 Mean | SD | Sig | n | BL Mean | SD | M6 Mean | SD | Sig |
| [12] | 5 | 41.2 | 24.7 | 39.0 | 29.4 | >0.05 | 5 | 50.6 | 15.0 | 64.8 | 12.5 | <0.05 |
| [39] | 30 | 54.8<sup>a</sup> | 24 | 45.2 | 27.7 | -- | 30 | 46.9 | 18.3 | 62.3 | 22.9 | -- |
| [40] | 25 | -- | -- | -- | -- | -- | 25 | 50.6 | 14 | 77.1 | -- | <0.01 |
| [41] | 19 | 1.8<sup>b</sup> | 0.2 | 2.4<sup>b</sup> | 0.3 | 0.24 | -- | -- | -- | -- | -- | -- |
| [42] | 16 | 65<sup>c</sup> | -- | 40<sup>c</sup> | -- | -- | -- | -- | -- | -- | -- | -- |
| [43] | 33 | 56.3 | 9.3 | 37.7 | 14.1 | <0.001 | 33 | 40.13 | 8.18 | 55.63 | 11 | <0.001 |

*Abbreviations: BL, baseline; M6, six months post baseline; n, number of subjects investigated; SD, standard deviation; Sig, level of significance (P values); --, not reported.*
*<sup>a</sup>, pain with Neer impingement sign.*
*<sup>b</sup>, scored on a five-point Likert scale (very bad, bad, poor, fair, good) and converted to a numerical score from 0 to 4;*
*<sup>c</sup>, median data.*

## Appendix 2

Note: throughout this document the term “former study” refers to the following study:

### Estimand of the present study according to *The International Council for Harmonisation of Technical Requirements for Pharmaceuticals for Human Use (ICH) E9 (R1) Addendum*

#### 1. Population

The population of the present study comprised subjects who fulfilled the following inclusion criteria and did not fulfill any of the following exclusion criteria:

##### 1.1 Inclusion criteria of the present study

1. Subject completed participation in RC-001 (NCT02918136) study.
2. Must have the ability to understand and sign a written informed consent form (ICF), which must be obtained prior to initiation of study procedures associated with this trial.

##### 1.2 Exclusion criteria of the present study

1. None.

##### 1.3 Inclusion criteria of the former study

1. Males and females 30-75 years of age.
2. Clinical symptoms consistent with a rotator cuff lesion including but not limited to pain, muscle weakness, or active-limited range of motion (AROM).
3. Subjects who have not responded to physical therapy treatments for at least six weeks.
4. Subjects with >70% passive range of motion (PROM).
5. Diagnosed with >50% tear to supraspinatus muscle or < 5mm separation assessed by MRI.
6. Diagnosed with a partial-thickness rotator cuff tear.
7. The ability of subjects to give appropriate consent.

##### 1.2 Exclusion criteria of the former study

2. Age <30 or >75.
3. Diagnosed with a full-thickness rotator cuff tear.
4. Insufficient amount of subcutaneous tissue to allow recovery of 50 mL of lipoaspirate.
5. History of systemic malignant neoplasms within last 5 years.
6. History of local neoplasm within the last 6 months and any history of local neoplasm at site of administration.
7. Subject is receiving immunosuppressant therapy or has known immunosuppressive or severe autoimmune disease that requires chronic immunosuppressive therapy (e.g., human immunodeficiency virus, systemic lupus erythematosus, etc.).
8. Subjects who are known to be HIV positive.
9. Patients who have received a corticosteroid injection in rotator cuff site within last 3 months.
10. Severe arthrosis of the glenohumeral or acromioclavicular joint.
11. Irreparable rotator cuff tear (including rotator cuff tear arthropathy).
12. Fatty atrophy above Grade 2 in affected shoulder.
13. Previous shoulder surgeries in affected shoulder.
14. Any contraindication to MRI scan according to MRI guidelines, or unwillingness to undergo MRI procedures.
15. History of tobacco use within the last 3 months.
16. Patient is on an active regimen of chemotherapy.
17. Patients with a documented history of liver disease or an ALT value >400.
18. Allergy to sodium citrate of any “caine” type of local anesthetic.
19. Patient is pregnant or breast feeding.
20. Subject is, in the opinion of the investigator or designee, unable to comply with the requirements of the study protocol or is unsuitable for the study for any reason. This includes completion of Patient Reported Outcome instruments.
21. Subject is currently participating in another clinical trial that has not yet completed its primary endpoint.
22. Subject is part of a vulnerable population who, in the judgment of the investigator, is unable to give Informed Consent for reasons of incapacity, immaturity, adverse personal circumstances or lack of autonomy. This may include: individuals with mental disability, persons in nursing homes, children, impoverished persons, persons in emergency situations, homeless persons, nomads, refugees, and those incapable of giving informed consent. Vulnerable populations also may include members of a group with a hierarchical structure such as university students, subordinate hospital and laboratory personnel, employees of the sponsor, members of the armed forces, and persons kept in detention.
23. Uncooperative patients or those with neurological/psychiatric disorders who are incapable of following directions or who are predictably unwilling to return for follow-up examinations.

Note: the criteria 20-23 were related to the former study (and, thus, also to the present study) and may not apply in management of sPTRCT using UA-ADRCs generated by the Transpose RT System (InGeneron Inc., Houston, TX, USA).

#### 2. Variables

The variables of the present study were the following:

##### 2.1 Variables evaluating long-term effectiveness

- ASES Total score (used in the primary endpoints)
- SF-36 Total score (used in the primary endpoints)
- Magnetic resonance imaging (used as secondary endpoint)
- VAS – Pain score (collected together with the ASES Total score)

##### 2.2 Variables evaluating long-term safety

- Adverse event rate between the UA-ADRCs group and the corticosteroid group

#### 3. Intercurrent Events

The possible intercurrent events of the present and the former studies comprised

- adverse events,
- use of concomitant medication,
- single missed study visits,
- loss to follow-up,
- additional assessments outside the visit window,
- discontinuation by investigator, and
- discontinuation by subject.

These possible intercurrent events, strategies to handle them according to ICH E9(R1) and imputation of possible subjects’ missing data related to each type of intercurrent event are described in detail in the following.

##### 3.1 Adverse events

Since the protocol of the present study specified group-specific comparisons of adverse events for the time periods (i) from BL to W24 post-treatment (considering only data of the former study, (ii) from BL to FSV in the present study, and (iii) from BL to SSV in the present study (each considering data of the present and the former studies), the following description of the different types of adverse events applies to the present and the former studies.

###### 3.1.1 Anticipated adverse events

This type of intercurrent event did not comprise serious adverse events, because no serious adverse events were anticipated in the present and the former studies.

This type of intercurrent event comprised fever, bleeding, bruising, persistent swelling at injection site, tenderness at injection site, pain at injection site, infection at injection site, redness or swelling at injection site, lightening of the skin around the injection site, joint infection, inflammatory flare, thinning of the skin and soft tissue around the injection site, tendon weakening, shoulder pain, worsening shoulder pain, nerve damage, death of nearby bone, calcium deposits on the tendon site, death of cartilage, potential allergic reactions (including anaphylaxis), prolonged numbness, tingling, a feeling of “pins and needles”, temporary skin, discoloration, itching or swelling where the medication was injected, the feeling of anxiousness, shakiness, dizziness, restlessness, or depression, drowsiness, vomiting and nausea.

This type of intercurrent event could occur in the UA-ADRCs group and the corticosteroid group, and the intercurrent events could or could not lead to subjects’ missing data.

###### 3.1.1.1 Anticipated adverse events that were possibly, probably or definitely related to the study treatments or to the liposuction procedure, were temporary, and were not serious adverse events

Anticipated adverse events could have possibly, probably or definitely been related to the study treatments or to the liposuction procedure. It was expected that most of these anticipated adverse events are temporary intercurrent events, and are not serious adverse events. These intercurrent events were expected to also occur in clinical use of UA-ADRCs generated by the Transpose RT System (InGeneron) or in clinical use of corticosteroid, and as such were handled using the Treatment Policy, i.e., whether this type of intercurrent event had occurred or not was irrelevant, and the data were collected and analyzed regardless. Possible subjects’ missing data related to this type of intercurrent event were imputed using the Last Observation Carried Forward approach.

This type of intercurrent event did occur in the present and the former studies (Tables S1 and S2).

###### 3.1.1.2 Anticipated adverse events at the index shoulder that were possibly, probably or definitely related to the study treatments and required additional injections into the index shoulder

If any of the anticipated adverse events *tendon weakening* (of a tendon in the index shoulder), *shoulder pain* (of the index shoulder), *worsening shoulder pain* (of the index shoulder), *calcium deposits on the tendon site* (of a tendon in the index shoulder), *death of nearby bone* (at the index shoulder) and *death of cartilage* (at the index shoulder) were possibly, probably or definitely related to the study treatments and required additional injections into the index shoulder after study treatment (e.g., injection of corticosteroid as additional treatment during the follow-up period, regardless of the study treatment), they were considered representing treatment failure. In this case these intercurrent events were handled using a combination of the While-on-Treatment Strategy and the Composite Strategy. Specifically, response to study treatment before the occurrence of the intercurrent event was handled using the While-on-Treatment Strategy, whereas response to study treatment after the occurrence of the intercurrent event was imputed according to the Composite Variable Strategy as minimum ASES Total score (0), minimum SF-36 Total score (0), maximum VAS pain score (10) and maximum tear volume measured on MRIs (150 mm^3^, which was greater than all data measured during the present and the former studies). Accordingly, after occurrence of these intercurrent events subjects’ missing data related to this type of intercurrent event were imputed as “failures” (i.e. non-responders).

This type of intercurrent event did occur in the present and the former studies (Tables S2 and S15).

###### 3.1.2 Unanticipated adverse events

This type of intercurrent event comprised non-serious adverse events and serious adverse events (note that no serious adverse events were anticipated in the present and the former studies).

This type of intercurrent event comprised the death of a subject, a life-threatening illness or injury, a permanent impairment of a body structure or a body function, hospitalization or prolonged existing hospitalization, or an important medical event defined as an event requiring medical or surgical intervention to prevent one of the outcomes listed above in this definition.

Although unanticipated, this type of intercurrent event could occur in the UA-ADRCs group and the corticosteroid group, and the intercurrent events could or could not lead to subjects’ missing data. Possible subjects’ missing data related to this type of intercurrent event were imputed using the Last Observation Carried Forward approach.

###### 3.1.2.1 Death of a subject in the UA-ADRCs group that would possibly, probably or definitely have been related to the study treatment (i.e., to the injection of UA-ADRCs)

In the extremely unlikely event of the death of a subject in the UA-ADRCs group that would possibly, probably or definitely have been related to the study treatment (i.e., to the injection of UA-ADRCs), it was planned to handle response to study treatment before the occurrence of the intercurrent event according to the While-on-Treatment Strategy, and to prematurely discontinue the study because evidence would have emerged that would have made the study unethical. In this case subjects’ missing data related to this type of intercurrent event would have been imputed as “failures” (i.e. non-responders).

This type of intercurrent event **did not occur** in the present and the former studies.

###### 3.1.2.2 Death of a subject in the UA-ADRCs group or the corticosteroid group that would possibly, probably or definitely have been related to the liposuction procedure

In the extremely unlikely event of the death of a subject in the UA-ADRCs group that would have been possibly, probably or definitely related to the liposuction procedure, it was planned to handle response to study treatment before the occurrence of the intercurrent event according to the While-on-Treatment Strategy, and to prematurely discontinue the study because evidence would have emerged that would have made the study unethical. In this case subjects’ missing data related to this type of intercurrent event would have been imputed as “failures” (i.e. non-responders).

This type of intercurrent event **did not occur** in the present and the former studies.

###### 3.1.2.3 Death of a subject in the corticosteroid group that would possibly, probably or definitely have been related to the study treatment (i.e., to the injection of corticosteroid)

In the extremely unlikely event of the death of a subject in the corticosteroid group that would possibly, probably or definitely have been related to the study treatment (i.e., to the injection of corticosteroid), it was planned to handle response to study treatment before the occurrence of the intercurrent event according to the While-on-Treatment Strategy. Subjects’ missing data related to this type of intercurrent event would have been imputed as “failures” (i.e. non-responders).

This type of intercurrent event **did not occur** in the present and the former studies.

###### 3.1.2.4 Death of a subject in the UA-ADRCs group or the corticosteroid group that would have been unlikely related or unrelated to the study treatments or the liposuction procedure (e.g., infection with SARS-CoV2 resulting in subject’s death), or whose causality could not have been determined

In the event of the death of a subject in the UA-ADRCs group or the corticosteroid group that would have been unlikely related or unrelated to the study treatments or the liposuction procedure (e.g., infection with SARS-CoV2 resulting in subject’s death), or whose causality could not have been determined, it was planned to handle response to study treatment before the occurrence of the intercurrent event according to the While-on-Treatment Strategy. Subjects’ missing data related to this type of intercurrent event would have been imputed using their last available observation.

This type of intercurrent event **did not occur** in the present and the former studies.

###### 3.1.2.5 Unanticipated, serious adverse events that were possibly, probably or definitely related to the study treatments, required surgical intervention, and were not joint infection

If any of the anticipated adverse events *tendon weakening* (of a tendon in the index shoulder), *shoulder pain* (of the index shoulder), *worsening shoulder pain* (of the index shoulder), *calcium deposits on the tendon site* (of a tendon in the index shoulder), *death of nearby bone* (at the index shoulder) and *death of cartilage* (at the index shoulder) were possibly, probably or definitely related to the study treatments and required surgery of the index shoulder after study treatment (i.e., during the follow-up period of the study), they were considered representing treatment failure.

In this case these intercurrent events were handled using a combination of the While-on-Treatment Strategy and the Composite Strategy. Specifically, response to study treatment before the occurrence of the intercurrent event was handled using the While-on-Treatment Strategy, whereas response to study treatment after the occurrence of the intercurrent event was imputed according to the Composite Variable Strategy as minimum ASES Total score (0), minimum SF-36 Total score (0), maximum VAS pain score (10) and maximum tear volume measured on MRIs (150 mm^3^, which was greater than all data measured during the present and the former studies). Accordingly, after occurrence of these intercurrent events subjects’ missing data related to this type of intercurrent event were imputed as “failures” (i.e. non-responders).

This type of intercurrent event did occur in the present and the former studies (Tables S1, S2 and S15).

###### 3.1.2.6 Joint infections of the index shoulder that were possibly, probably or definitely related to the study treatments and require surgical intervention

If the anticipated adverse event *joint infection* (of the index shoulder) would have required surgery of the index shoulder after study treatment, it would have been necessary to rate this type of intercurrent event as serious adverse events. However, it was not anticipated that such intercurrent events would occur in the present and the former studies.

In case such an intercurrent event would have occured in the present and the former studies, response to study treatment before the occurrence of the intercurrent event would have been handled according to the While-on-Treatment Strategy, whereas response to study treatment after the occurrence of the intercurrent event would have been imputed according to a Hypothetical Strategy in which the intercurrent event would not have occured (because the intercurrent event would have been related to the injection procedure but not to the study treatment itself). Possible subjects’ missing data related to this type of intercurrent event would have been imputed using the Last Observation Carried Forward approach.

This type of intercurrent event **did not occur** in the present and the former studies.

###### 3.1.2.7 Unanticipated serious adverse events that were possibly, probably or definitely related to the study treatments or the liposuction procedure, but were not related to the index shoulder

All available (published and unpublished) data indicated that this type of intercurrent event would not occur in the present and the former studies.

In the unlikely event that this type of intercurrent event would have nevertheless occurred in the UA-ADRCs group or the corticosteroid group (with the possibility of subjects’ missing data) during the present and the former studies, response to study treatment before and after the occurrence of the intercurrent event would have been handled according to Treatment Policy, i.e., whether this type of intercurrent event would have occurred or not is irrelevant, and the data would have been collected and analyzed regardless. Possible subjects’ missing data related to this type of intercurrent event would have been imputed using the Last Observation Carried Forward approach.

This type of intercurrent event **did not occur** in the present and the former studies.

###### 3.1.2.8 Adverse events and serious adverse events that were unlikely related or unrelated to the study treatments or the liposuction procedure, or whose causality could not be determined, were related to the index shoulder (e.g., accidents involving the index shoulder) but did not result in the subjects’ death

This type of intercurrent event could occur in the UA-ADRCs group and the corticosteroid group, and the intercurrent events could or could not lead to subjects’ missing data.

In case this type of intercurrent event occured in the present and the former studies, it was handled using a combination of the While-on-Treatment Strategy and a Hyopthetical Strategy. Specifically, response to the study treatment before the occurrence of the intercurrent event was handled according to the While-on-Treatment Strategy, whereas response to the study treatment after the occurrence of the intercurrent event was imputed according to a Hypothetical Strategy in which the intercurrent event would not occur. Possible subjects’ missing data related to this type of intercurrent event was imputed using the Last Observation Carried Forward approach.

This type of intercurrent event did occur in the present and the former studies (Tables S1, S2 and S15).

###### 3.1.2.9 Non-serious adverse events that were unlikely related or unrelated to the study treatments or the liposuction procedure, or whose causality could not be determined, and were not related to the index shoulder (e.g., infection with SARS-CoV-2 that did not require hospitalization)

This type of intercurrent event could occur in the UA-ADRCs group and the corticosteroid group, and the intercurrent events could or could not lead to subjects’ missing data.

In case this type of intercurrent event occured in the present and the former studies, it was handled according to Treatment Policy, i.e., whether this type of intercurrent event had occurred or not was irrelevant, and the data were collected and analyzed regardless. Possible subjects’ missing data related to this type of intercurrent event were imputed using the Last Observation Carried Forward approach.

This type of intercurrent event did occur in the present and the former studies (Tables S1 and S2).

###### 3.1.2.10 Serious adverse events that would have been unlikely related or unrelated to the study treatments or the liposuction procedure, or whose causality could not have been determined, would not have been related to the index shoulder, and would have made collection of study data temporarily impossible (e.g., infection with SARS-CoV-2 that would have required hospitalization or car accidents)

This type of intercurrent event could have occured in the UA-ADRCs group and the corticosteroid group, and the intercurrent events could or could not have led to subjects’ missing data.

In case this type of intercurrent event would have occured in the study, it would have been handled using a combination of the While-on-Treatment Strategy and a Hyopthetical Strategy. Specifically, response to the study treatment before the occurrence of the intercurrent event would have been handled according to the While-on-Treatment Strategy, whereas response to the study treatment after the occurrence of the intercurrent event would have been imputed according to a Hypothetical Strategy in which the intercurrent event would not have occured. Possible subjects’ missing data related to this type of intercurrent event would have been imputed using the Last Observation Carried Forward approach.

This type of intercurrent event **did not occur** in the present and the former studies.

##### 3.2 Use of concomitant medication (except for injections into the index shoulder after randomization that did not represent study treatments)

This type of intercurrent event could occur in the UA-ADRCs group and the corticosteroid group.

In case this type of intercurrent event occured during the present and the former studies, response to study treatment before and after the occurrence of the intercurrent event was handled according to Treatment Policy, i.e., the data was collected and analyzed regardless.

This type of intercurrent event did occur in the present and the former studies (data not shown).

##### 3.3 Single missed study visits

This type of intercurrent event could have occured in the UA-ADRCs group and the corticosteroid group, and the intercurrent events could have led to subjects’ missing data.

In case this type of intercurrent event would have occurred during the present and the former studies, response to study treatment before and after the occurrence of the intercurrent event would have been handled according to a Hypothetical Strategy in which the intercurrent event would not have occured. Subjects’ missing data related to this type of intercurrent event would have been imputed using the Last Observation Carried Forward approach.

This type of intercurrent event **did not occur** in the present and the former studies.

##### 3.4 Loss to Follow-up

Loss to follow-up would have occured when a subject would have missed two consecutive, scheduled follow-up time points (study visits). If attempts to contact the subject or subject’s healthcare provider would have been unsuccessful, then the subject would have been considered lost-to-follow-up.

This type of intercurrent event could have occured in the UA-ADRCs group and the corticosteroid group, and the intercurrent events could have led to subjects’ missing data.

In case this type of intercurrent event would have occurred during the present and the former studies, we would have determined whether loss to follow-up is related to any of the types of intercurrent events outlined in this section, and would have applied the corresponding strategy for addressing the intercurrent event and adequate imputation of subjects’ missing data. Otherwise this type of intercurrent event would have been handled using a combination of the While-on-Treatment Strategy and a Hyopthetical Strategy. Specifically, response to the study treatment before the occurrence of the intercurrent event would have been handled according to the While-on-Treatment Strategy, whereas response to the study treatment after the occurrence of the intercurrent event would have been imputed according to a Hypothetical Strategy in which the intercurrent event would not have occured. Subjects’ missing data related to this type of intercurrent event would have been imputed using the Last Observation Carried Forward approach.

This type of intercurrent event **did not occur** in the present and the former studies.

##### 3.5 Assessments outside the study visit windows

This type of intercurrent event could occur in the UA-ADRCs group and the corticosteroid group.

In case this type of intercurrent event occured during the present and the former studies, response to study treatment before and after the occurrence of the intercurrent event was handled according to a Hypothetical Strategy in which the intercurrent event would not occur.

This type of intercurrent event did occur in the present and the former studies (Table S15).

##### 3.6 Discontinuation by investigator because the subject moved out of the country

This type of intercurrent event could have occured in the UA-ADRCs group and the corticosteroid group, and the intercurrent events could have led to subjects’ missing data.

In case this type of intercurrent event would have occured during the present and the former studies, it would have been handled using a combination of the While-on-Treatment Strategy and a Hyopthetical Strategy. Specifically, response to the study treatment before the occurrence of the intercurrent event would have been handled according to the While-on-Treatment Strategy, whereas response to the study treatment after the occurrence of the intercurrent event would have been imputed according to a Hypothetical Strategy in which the intercurrent event would not have occured. Possible subjects’ missing data related to this type of intercurrent event would have been imputed using the Last Observation Carried Forward approach.

This type of intercurrent event **did not occur** in the present and the former studies.

##### 3.6 Discontinuation by investigator for other reasons than the subject having moved out of the country

This type of intercurrent event could occur in the UA-ADRCs group and the corticosteroid group, and the intercurrent events could lead to subjects’ missing data.

In case this type of intercurrent event occured during the present and the former studies, the corresponding strategy for addressing the intercurrent event and adequate imputation of missing data was performed according to the reason that motivated the Investigator to discontinue the subject’s participation in the study (these reasons are addressed in this section).

This type of intercurrent event did occur in the present and the former studies (Table S15).

##### 3.7 Discontinuation by subject

This type of intercurrent event could occur in the UA-ADRCs group and the corticosteroid group, and the intercurrent events could lead to missing data.

Subjects’ participation in the present and the former studies was voluntary, and the subjects could discontinue participation (refuse all subsequent testing/follow-up) at any time without loss of benefits or penalty.

In case this type of intercurrent event occurred during the present and the former studies, we determined whether withdrawal of consent by the subject was related to any of the types of intercurrent events outlined in this section, and applied the corresponding strategy for addressing the intercurrent event and adequate imputation of missing data. Otherwise this type of intercurrent event was handled using a combination of the While-on-Treatment Strategy and a Hypothetical Strategy. Specifically, response to the study treatment before the occurrence of the intercurrent event was handled according to the While-on-Treatment Strategy, whereas response to the study treatment after the occurrence of the intercurrent event was imputed according to a Hypothetical Strategy in which the intercurrent event did not occur. Subjects’ missing data related to this type of intercurrent event were imputed using the Last Observation Carried Forward approach.

This type of intercurrent event did occur in the present and the former studies.

#### 4. Population-Level Summary

The Population-Level Summary of the present study (i.e., the variables on which the comparison between treatments were based) is described in detail in the Sections “*Outcome measurements and assessments*”, “*Analysis of MRI scans*” and “*Statistical Analysis*” in the *Methods* section of this paper.

#### 5. Missing data

Missing data were handled according to the strategies outlined in Section 3 (“*Intercurrent Events*”) of this appendix.

